# Healthome Polygon Framework: Comprehensive and Multi-dimensional Health Quantification Framework Using Artificial Intelligence and Multiomics Data

**DOI:** 10.1101/2025.03.10.25323468

**Authors:** Youngmin Bhak, Eun-Seok Shin, Sungwon Jeon, Jong Bhak

## Abstract

**Background:** Quantifying human health and disease necessitates a transformative framework capable of integrating diverse biomedical data in a standardized manner. Such a system could enable precise preemptive health and disease prevention through the application of artificial intelligence.

**Objectives:** We propose a novel health quantification framework using Pseudo Super Healthy (PSH) subjects—virtual optimal health conditions, and the Multi-Omics Health Index (MOHI) which consolidates health scores across eight “omes”: Physiome, Metabolome, Vasculome, Inflammatome, Immunome, Psycholome, Transcriptome, and Epigenome.

**Methods:** PSH subjects were identified using algorithmic or clinical domain knowledge-based methods in the first five omes, utilizing data from the Korea National Health and Nutrition Examination Survey and the Korean Genome Project (KGP). Individual health status in these omes was quantified using Euclidean distances (ED) from PSH subjects. For the last three omes, machine learning (ML) models trained on KGP data predicted individual health probabilities.

**Results:** PSH subjects were stratified by age and sex. In two available case studies—a male in his mid-50s and a female in her mid-30s—their MOHI scores were 30 and 49, respectively. ED-based health indicators correlated significantly with the Number of Abnormal Traits (NATs) in Physiome (Pearson’s r = 0.62 for males, 0.70 for females, *P* < 0.001). Three ML models used to quantify health for the Psycholome, Transcriptome, and Epigenome achieved area under the curve values of 0.79, 0.85, and 0.96, respectively.

**Conclusions:** Individual health can be represented by a radar chart (Healthome Polygon) derived from PSH subjects and ML models, resulting in MOHI scores that serve as a personalized health quantification metric.

## Introduction

The traditional approach to health monitoring and assessing has predominantly relied on isolated measures such as clinical diagnoses, laboratory tests, or imaging. However, as our understanding of human biology deepens, it has become evident that health is a multi-dimensional construct influenced by a complex interplay of molecular, cellular, and physiological factors.[1] This complexity underscores the limitations of conventional methods, which often fail to capture the holistic nature of health. In the era of personalized and precision medicine, there is an increasing demand for comprehensive and integrated health assessment tools that can provide accurate insight into an individual’s unique health status and risk factors.[2] One promising approach to addressing this challenge lies in the integration of multiomics data, which encompasses multiple biological layers such as genomics, transcriptomics, proteomics, metabolomics, and epigenomics.[3] When combined with advanced machine learning (ML) technologies, multiomic data has the potential to revolutionize healthcare by offering a standardized and comprehensive representation of an individual’s health status. This integration enables preventive healthcare, early disease detection, and tailored therapeutic interventions.[4,5]

Recent efforts to develop health indices have demonstrated both progress and limitations. For instance, Kaiser et al. proposed a multidimensional health index using 21 health scores visualized in a radar chart,[6] while, Pano et al. constructed a lifestyle and well-being index based on multivariate linear models using twelve lifestyle items.[7] Although these studies provide valuable insights, they face challenges in interpretation due to the complexity of managing multiple health scores and dimensions, which often exceed the optimal number of easy comprehension.[6] Furthermore, these indices lack quantitative health traits beyond self-reported questionnaires and do not incorporate genomic data. Crucially, neither study identifies individuals with optimal health status, which could serve as a reference for health assessment.[6,7]

To address these limitations, we propose the Multi-Omics Health Index (MOHI), a novel framework designed to quantify and integrate diverse omics layers into a unified health index. Accompanied by an intuitive visualization tool, Healthome Polygon (HP), this framework provides a comprehensive solution for health assessment.

MOHI integrates data from eight critical omics domains—Physiome, Metabolome, Vasculome, Inflammatome, Immunome, Psycholome, Transcriptome, and Epigenome—reflecting the interconnectedness of biological systems that influence health, for example, alterations in the genome can lead to changes in the transcriptome and metabolome which in turn may impact immune function and disease susceptibility.[8]

Specifically about each ome, Physiome is a quantitative description of the totality of an organism’s physiological state and functional behavior. It’s a term that combines the words “physio” (life) and “ome” (as a whole).[9] Metabolome is all low molecular weight metabolites within a cell, both qualitatively and quantitatively, that are involved in metabolic reactions essential for its maintenance, growth, and function.[10] Vasculome provides a quantitative representation of an organism’s vascular system, capturing its structural state, functional dynamics, blood vessel architecture, hemodynamics, and endothelial interactions. Inflammatome quantitatively characterizes an organism’s inflammatory state and functional activity, encompassing mediators such as cytokines, cellular interactions, and the balance between pro- and anti-inflammatory processes. Immunome is the set of factors that constitute and reflect the immune system. Psycholome provides a quantitative assessment of an organism’s psychological state and functioning, including cognitive processes, emotional states, behavioral patterns, and neural activity. Transcriptome includes all coding and non-coding RNA transcripts present in a cell or group of cells.[11] Epigenome of an organism includes chemical alterations to its DNA and histone proteins that control when, where, and how genes are expressed. These changes can modify chromatin structure and impact genome function.[12]

By leveraging a multiomics approach, MOHI offers a more accurate and dynamic representation of an individual’s health status than traditional single-dimensional metrics.

The foundation of MOHI lies in the conceptualization and construction of Pseudo Super Healthy (PSH) subjects, which serve as a reference for optimal health. These PSH subjects are generated using sophisticated ML algorithms that analyze large-scale omics datasets, such as those from the Korean Genome Project (KGP), to identify patterns indicative of optimal health. By comparing an individual’s multiomics profile to that of the PSH subjects, health status is quantified using the Euclidean distance metric, providing a numerical health score that reflects the alignment with optimal health. These scores are visualized through HP, a radar chart that intuitively displays the multidimensional aspects of health, enabling healthcare providers and researchers to easily interpret and act upon the results.

## Methods

### The Korea National Health and Nutrition Examination Survey data to establish Pseudo Super Healthy subjects

The Korea National Health and Nutrition Examination Survey (KNHANES),[13] conducted annually by the Korea Centers for Disease Control and Prevention (KCDC), is a comprehensive assessment of nutritional and health status in South Korea. From the 2009-2021 survey period, 83,935 subjects were retained from 105,843 subjects with 24 health check-up covariates. Seven key covariates—Age, Systolic Blood Pressure, Diastolic Blood Pressure, Fasting Blood Sugar, Triglycerides, Cholesterol, and Hemoglobin—were identified as impactful compared to others such as Sex, Height, Weight, Waist circumference, BMI, Hematocrit, Platelets, RBC, WBC, HbA1c, HDL, Blood Urea Nitrogen, Creatinine, ALT, AST, Urine pH, and Urine specific gravity. Three models were developed using these covariates and additional model-specific traits: the Physiome model (2009-2021), the Metabolome Thyroid model (2013-2015), and the Metabolome Uric Acid model (2016-2021). The utilization of KNHANES within the HP framework’s overall process is described in both Figure 1 and Supplementary Figure 1.

**Figure 1.**
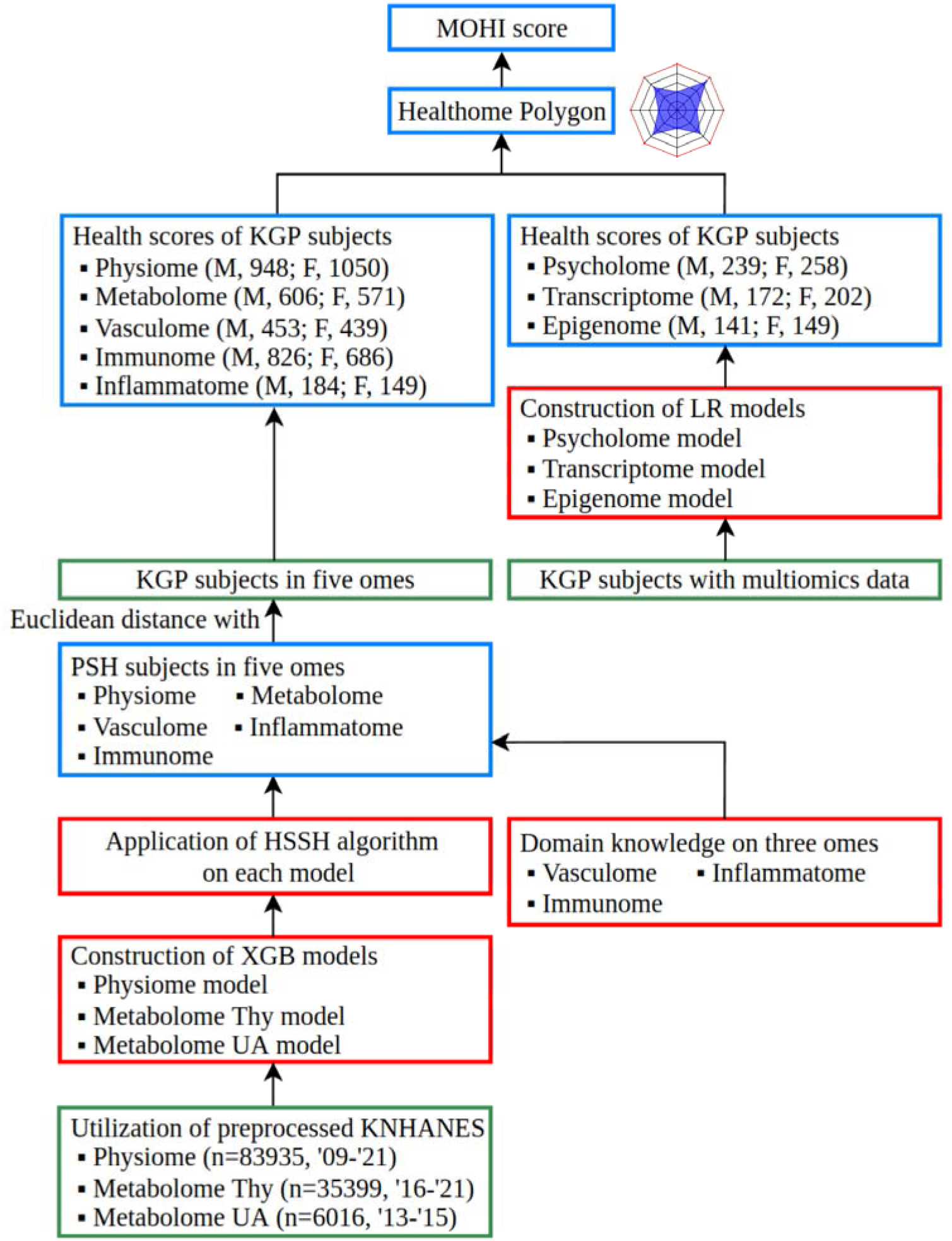
The flowchart of implementation for Healthome Polygon framework. This figure illustrates the overall process of constructing HP framework. Pseudo Super Healthy subjects for five omes (Physiome, Metabolome, Vasculome, Inflammatome, Immunome) were identified using a combination of clinical domain knowledge and the HSSH algorithm, which is derived from an XGBoost model trained on the KNHANES database. Subsequently, the Euclidean distance between the PSH subjects and each KGP subject was calculated to determine health scores for the five omes. In addition, the multiomics dataset from KGP was used to generate health scores for Psycholome, Transcriptome, and Epigenome. Health scores across all eight ome-components were visualized using HP, the radar charts, and the MOHI was calculated. For additional information, boxes related to databases, methods, and results were colorized by green, red, and blue, respectively. HP, Healthome Polygon; PSH, Pseudo Super Healthy; HSSH, Health check-up data and a SHAP-value-based pseudo Super Healthy; KNHANES, The Korea National Health and Nutrition Examination Survey; MOHI, Multi-Omics Health Index; Thy, Thyroid; UA, Uric Acid; KGP, Korean Genome Project; XGB, eXtreme Gradient Boosting; LR, Logistic Regression; M, Male; F, Female

### Defining Pseudo Super Healthy subjects in five ome-components through data-driven artificial intelligence method and utilizing empirical domain knowledge

Defining point-estimated Super Healthy subjects is challenging because clinical normality is range-based. Therefore, we adopted a data-driven AI model-based approach to define point-estimated Super Healthy subjects, which we term Pseudo Super Healthy (PSH) subjects. To define PSH subjects, two primary methods were used: a supervised learning-based model and the HSSH algorithm (Figure 2).

**Figure 2.**
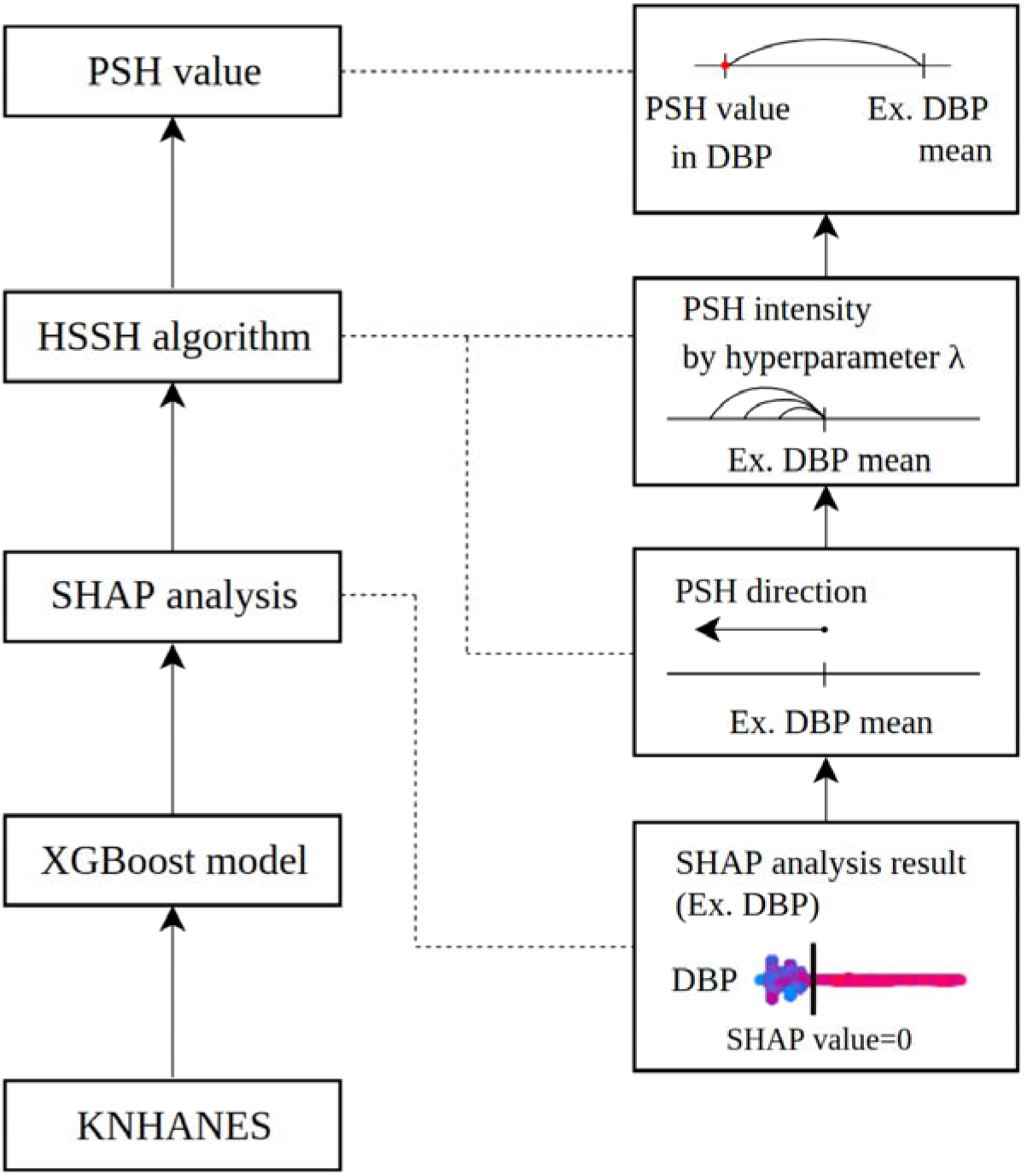
The flowchart of the identification process for PSH subjects. The KNHANES database was used to train a binary classification model using the XGBoost algorithm and unhealthy label information. The trained model was interpreted using the SHAP algorithm. By applying the HSSH algorithm to the analyzed SHAP values, the PSH direction—either left or right from the trait’s mean value—was determined. Next, the PSH intensity, representing the distance from the trait’s mean, was defined by the hyperparameter **λ**, which is set by the user. In the figure, three PSH intensity values are shown as examples, corresponding to three different hyperparameter ***λ*** values. Finally, the PSH value which is marked as a red point for each trait was calculated. The boxes for SHAP analysis, the HSSH algorithm, and the PSH value were connected to their corresponding boxes with dashed lines for a detailed illustration. In the figure, DBP is provided as an example. PSH, Pseudo Super Healthy; HSSH, Health-check-up data and SHAP-value-based pseudo Super Healthy subject; XGBoost, eXtreme Gradient Boosting; KNHANES, The Korea National Health and Nutrition Examination Survey; DBP, Diastolic Blood Pressure

#### Supervised Learning on XGBoost model with preprocessed KNHANES data

To define Physiome, using 83,935 subjects of the KNHANES dataset, we developed an XGBoost-based classification model for identifying unhealthy individuals.[14] For the classification of unhealthiness, we collected 26 traits of diagnosis information provided by medical professionals, 26 traits of self-reported current disease information, and six traits generated from health examinations for each subject. Each trait encompasses three categories: True for presence of disease, False for absence of disease, and unknown or no-response. We treated instances of unknown or no-response as False to keep as high as possible data sample size, under the assumption that the majority of instances falling under unknown or no-response category correspond to absence of disease. Subsequently, if any of the 58 traits were labeled as True, the subject was categorized as unhealthy; otherwise, they were deemed healthy. Supplementary Table 1 and Supplementary Table 2 provide a breakdown of the 58 traits associated with disease names and their statistics regarding True and False labels. Then, subjects taking medication for hypertension, dyslipidemia, or diabetes were excluded. Missing values were imputed using a multivariate imputation method employing IterativeImputer() from scikit-learn library.[15] The dataset was divided into train, validation, and test sets comprising 50,361, 16,787, and 16,787 samples respectively, with labels stratified across the datasets. The train set was utilized for training the model, while the validation set was employed to assess model performance during training. Hyperparameter optimization was conducted using Optuna to maximize the Area Under the Curve (AUC) calculated from the validation dataset.[16] The test set was used to evaluate the final performance after the completion of the model training. For Metabolome Thyroid and Metabolome Uric acid models, same procedure to the above Physiome model was applied.

#### HSSH algorithm to find pseudo super healthy subject with optimal value of Health-check-up data traits

After training and testing the XGBoost model, the HSSH (Health-check-up data and SHAP-value-based pseudo Super Healthy subject) algorithm was applied to the trained model. The HSSH algorithm automatically determines the PSH direction and PSH intensity needed to identify PSH value and PSH subject (Figure 2).

In the Physiome SHAP summary plot (Supplementary Figure 2A), the Diastolic Blood Pressure (DBP) trait exhibits a concentration of dominant red dots in the area where the SHAP value is greater than zero.[17,18] This indicates that a high DBP value contributes to unhealthiness. Consequently, it can be inferred that lowering DBP below the mean DBP value promotes healthiness, aligning with the identified optimal direction, which is defined as the PSH direction. Additionally, the extent to which the DBP value should deviate from the mean (PSH intensity) to achieve the optimal value (PSH value) is determined after establishing the direction and is fine-tuned using the hyperparameter λ (Figure 2).

To determine the direction, the dominant color was quantitatively identified by counting the red and blue dots in the region where the SHAP value exceeded zero. SHAP values were computed for 24 traits across 50,361 subjects in the training set. For each subject and trait, the SHAP value was evaluated to check if it exceeded zero. If it did, the corresponding trait value for that subject was compared to the mean value of the trait column. If the trait value was higher than the mean, the red dot count was incremented; if it was lower, the blue dot count was incremented.

This process allowed the dominant color, either red or blue, to be determined by comparing the number of red and blue dots under the condition that the SHAP value was greater than zero. Subsequently, the log ratio of the red and blue dot counts was calculated by dividing the two counts and applying a logarithmic function to stabilize the variability in the resulting values. The mean value of the trait column (*µ_trait_*), the log ratio (*log R*), and the hyperparameter value (λ) were then substituted into the following equations. Eq1 was used when *x_optimal_*was smaller than *µ_trait,_* while Eq2 was otherwise.

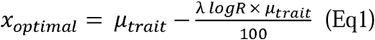

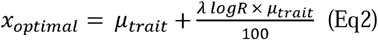

where, λ ∈ {x ∈ ℝ | x > 0} is a user-defined hyperparameter that generates extreme pseudo super healthy (PSH) subjects by allowing values to deviate further from the mean as λ increases. For this study, λ=1.0 was used after several experiments empirically. The procedure for establishing PSH subjects within the HP framework’s overall process is thoroughly described in Supplementary Figure 1.

#### Utilization of Domain knowledge to find pseudo super healthy subjects in Vasculome, Inflammatome, and Immunome

Due to the lack of associated traits for the Vasculome, Inflammatome, and Immunome components in the KNHANES database, PSH subjects in these omes were identified using appropriate PSH values. For the Vasculome, the PSH value for the homocysteine trait was set to the minimum homocysteine value within each sex and age group. Since the Physiome KNHANES model incorporates age as a covariate, PSH age values were determined as being in the 30s for both males and females. Accordingly, the minimum homocysteine values for males and females in their 30s were used as the homocysteine PSH values.

For Immunome traits, such as WBC, monocyte, neutrophil, eosinophil, basophil, and lymphocyte counts, PSH values were set to the median value, based on reports indicating that both abnormally low and high levels of these immune traits are signs of unhealthiness.[19] For Inflammatome traits, such as Rheumatoid Factor and C-Reactive Protein, PSH values were set to zero, as values outside the normal range indicate inflammation.

Additionally, for unavailable traits in the KNHANES database, such as LDL and GGT within the Physiome component, PSH values were derived as follows: the PSH value for LDL was set to the mean LDL value for females, while the PSH value for GGT was set to a value lower than the male mean value.

### Developing the five ome-components of Healthome Polygon using KGP health check-up data

Each ome-component dataset, which is used for this study, from the Korean Genome Project (KGP) dataset was curated with ome-relevant traits (Table 1). The KGP dataset was previously published as part of the Korea1K and Korea4K studies, which provide large and precise multiomics data for the Korean population.[20,21] However, it lacks direct label information, such as unhealthiness. To address this limitation, the KNHANES dataset was incorporated to complement the unhealthiness labels, along with the expanded Korea10K version of the KGP dataset, which includes more subjects than Korea1K and Korea4K, to quantify individual health using AI. As part of this process, Pseudo Super Healthy subjects were defined, and the final quantified health scores were visualized.

**Table 1.** Overview of KGP cohort datasets used to develop Healthome Polygon framework, the individual’s health status quantification framework.

|  | # of features | # of subjects |
| --- | --- | --- |
| Clinome (Health-Check-up) | 330 | 6719 |
| Physiome <sup>a</sup> | 15 | 2196 |
| Metabolome <sup>a</sup> | 5 | 974 |
| Vasculome <sup>a</sup> | 6 | 905 |
| Immunome <sup>a</sup> | 6 | 1513 |
| Inflammatome <sup>a</sup> | 2 | 333 |
| Lifestylome (Lifestyle Questionnaires) | 657 | 4925 |
| Transcriptome | 69,222 | 926 |
| Transcriptome after preprocessing <sup>b</sup> | 23,227 | 926 |
| Epigenome | 663,904 | 1211 |

<sup>a</sup> From the final state after the preprocessing
<sup>b</sup> Redundantly duplicated and unexpressed (median=0) transcript columns were excluded #, The number
<sup>b</sup> Redundantly duplicated and unexpressed (median=0) transcript columns were excluded

Subsequent preprocessing steps, such as selection of samples, imputation for missing values, and exclusion of outliers, were executed on KGP dataset (Supplementary Figure 3 and Supplementary Method 1).

Euclidean distances between PSH and KGP subjects were calculated for the five ome components: Physiome, Metabolome, Vasculome, Immunome, and Inflammatome. For the male group, the Euclidean distances were computed for 948, 606, 453, 826, and 184 KGP subjects, respectively, while for the female group, they were computed for 1,050, 571, 439, 686, and 149 KGP subjects, respectively, after standardizing for each sex and for each PSH subject pertaining to an ome component using Eq3 in the respective ome component.

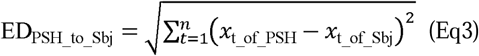

where, *t* represents the trait, *n* is the number of traits within each ome-component, and *x* denotes the trait value.

To assess whether the Euclidean distance between PSH and KGP subjects serves as a valid health indicator, we introduced the NATs (Number of Abnormal Trait values) for each subject. This metric was designed to exhibit a positive Pearson’s correlation coefficient between the Euclidean distance and the number of abnormal trait values if the Euclidean distance accurately reflects health status. The procedure for calculating Euclidean distance within the HP framework’s overall process is described in Supplementary Figure 1.

### Utilizing KGP multiomics data to construct the Psycholome, Transcriptome, and Epigenome components of the Healthome Polygon

In contrast to the five ome-components (Physiome, Metabolome, Vasculome, Immunome, and Inflammatome), the Psycholome, Transcriptome, and Epigenome components utilized multiomics data from the KGP dataset, and health status on those omes was quantified based on predicted health probabilities from a supervised logistic regression model.

Psychological unhealthiness traits, e.g., suicide attempt, anxiety disorder, and depressive disorder, included in KGP dataset were used to label Psycholome dataset which is multiomic data composed of both transcriptome and epigenome. However, since explicit unhealthiness labels did not exist in the KGP database, pseudo-unhealthiness labels were used for the Transcriptome and Epigenome datasets. These labels were predicted by a logistic regression model trained on the KNHANES dataset (83935 subjects, unhealthy 44541, healthy 39394, male 38861, female 45074, 14 covariates: Sex, Age, Waist circumference, BMI, SBP, DBP, Hemoglobin, Fasting Blood Sugar, HDL, Triglyceride, Cholesterol, Creatinine, ALT, AST, unhealthiness label was generated in the same way of Physiome). The validation for the pseudo unhealthiness label was performed by using chi-square test between pseudo unhealthiness label and the questionnaires of KGP dataset on 171 number of diseases, clinical states, or health-checkup traits (Supplementary Table 5).

For preprocessing these KGP multiomics datasets, redundantly duplicated and unexpressed gene columns were excluded from the transcriptomic gene expression value data. Batch effects from different sequencing platforms and years were mitigated using the pyComBat library,[22] applied to both gene expression and epigenomic beta value data. Batch-effect correction was assessed using scatter plots of data subjected to dimensionality reduction via Principal Component Analysis (PCA) and deep learning based Autoencoder.[23,24]

Subsequently, these datasets were performed by feature selection using a log likelihood test.[25] The compositions for null and alternative models were chosen which resulted in the best proper number of selected features and the best predictive performance. For Psycholome with 682348 multiomic features, gene_expression (y) ∼ age + sex (x), gene_expression (y) ∼ age + sex + psychological_unhealthy_label (x), for Transcriptome with 18444 transcript features, pseudo_unhealthy_label (y) ∼ sex (x), pseudo_unhealthy_label (y) ∼ sex + gene_expression (x), for Epigenome with 663,904 epigenomic features, pseudo_unhealthy_label (y) ∼ sex (x), pseudo_unhealthy_label (y) ∼ sex + CpG_beta_value (x) were used as null and alternative models, respectively. All transcript and epigenomic features were iteratively applied to the models, resulting in *p*-values which stand for the statistically significant difference between null and alternative models. The *p*-values for the log-likelihood test on Psycholome, Transcriptome, and Epigenome were adjusted by Benjamini-Hochberg method.[26] The adjusted *p*-value cutoff was set at <0.05.

With the selected traits, the final three logistic regression models with 10-fold cross-validation using CalibratedClassifierCV were developed for Psycholome with 497 subjects (unhealthy 162, healthy 335 of male 239 and female 258), Transcriptome with 374 subjects (unhealthy 199, healthy 175 of male 172 and female 202), and Epigenome with 290 subjects (unhealthy 166, healthy 124 of male 141 and female 149).[15] The 95% confidence intervals (CI) were calculated by using bootstrap resampling.[27] The procedure for predicting psychological health probabilities within the HP framework’s overall process is described in Supplementary Figure 1.

### Healthome Polygon: comprising eight health-related components to quantify an individual’s health status

#### Construction of Healthome Polygon by gathering scores from eight ome-components

The Euclidean distance value of each KGP subject for each ome-component (Physiome, Metabolome, Vasculome, Immunome, and Inflammatome) was transformed into a score ranging from 0 (representing unhealthy) to 100 (representing healthy). Similarly, the predicted health probability on Psycholome, Transcriptome, and Epigenome was adjusted to fall within a range of 0 (unhealthy) to 100 (healthy). The above-mentioned eight “omes” represent physiology, metabolism, cardiovascular system, immunity, inflammation, psychology, the transcriptome, and the epigenome. These eight health scores were then depicted on a radar chart using Pygal library.[28]

The arrangement of the eight ome components on the radar chart was determined by the correlation between their paired scores (Supplementary Table 9) rather than by random positioning, as the arrangement affects the polygon’s area. Therefore, negatively correlated ome components were positioned in opposite directions, whereas positively correlated components were placed adjacent to each other as much as possible to ensure a reasonable arrangement of omes from both meaningful and analytical perspectives.

Subsequently, the area of the radar chart corresponding to each subject was computed and normalized to a scale of 0 (unhealthy) to 100 (healthy), thereby constituting the MOHI, which serves to quantify an individual’s health across the eight dimensions or omes.

#### Detailed method for calculating and normalizing radar area

To compute the area of each triangle within the radar chart, we initially divided 360 degrees by eight to obtain the contained angle θ*_i_* for two lines in a triangle using Eq4, resulting in 45 degrees equivalent to 0.79 radians.

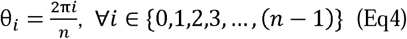

Then the Cartesian coordinates of the i-th point are calculated by Eq5.

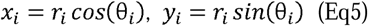

where, *r_i_* is i-th ome-component health score. Then, each triangle’s area *t_i_* could be calculated by Eq6.

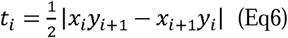

To close the HP, start and end points should be identical like (*x_n,_ y_n_*) = (*x_0,_ y_0_*).

Finally, total area *T* can be calculated by summing up the area of all triangles like Eq7.

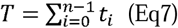

The calculated total area T should be normalized into 0 to 100 scale. For that purpose, to obtain a normalizing scale factor γ which divides total area T, maximum area 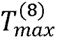 of HP whose health scores from all eight ome-components are 100 was calculated as Eq8.

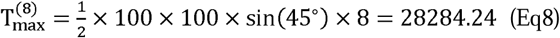

Then, normalizing scale factor γ which makes 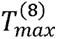 100 was calculated by Eq9.

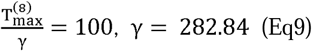

Finally, the MOHI of one subject was defined as 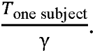

### Explainability of Healthome Polygon and MOHI

Sex-stratified datasets for five omes (Physiome, Metabolome, Vasculome, Immunome, and Inflammatome) from the KGP cohort, as well as sex-stratified PSH subjects’ data for the same five omes, were min-max normalized. The differences between each subject and the PSH data were then calculated. Thresholds were set arbitrarily at the 50th and 75th percentiles of these differences, considering these thresholds as the ideal criteria for distinguishing between normal and abnormal values within the bell-shaped distribution of the differences. If a target subject’s difference from the PSH data for each trait falls below the 50th percentile, it is classified as low abnormality. Similarly, differences falling within 50th to <75th percentiles are classified as moderate abnormality, while differences ≥75th percentile are classified as high abnormality. For Psycholome, Transcriptome, and Epigenome, each model was explained by SHAP waterfall plots. The SHAP waterfall plot explains the contribution pattern of each feature to the predicted probability.[17]

## Results

### Healthome Polygon of two selected KGP subjects

The health quantification framework, which reflects the concepts outlined in the Introduction, was successfully developed. Only two KGP subjects, who had data for all eight omes, were included as examples of the framework’s results.

The health statuses of these two subjects (KGP-04332 and KGP-10305, a male in his mid-50s and a female in her mid-30s) from the KGP cohort were quantified using MOHI scores of 30 and 49, respectively (Figure 3). These two subjects initially had scores for seven ome-components, excluding the Vasculome. Therefore, homocysteine values were imputed based on their corresponding age group and sex, resulting in health scores for all eight ome-components.

**Figure 3.**
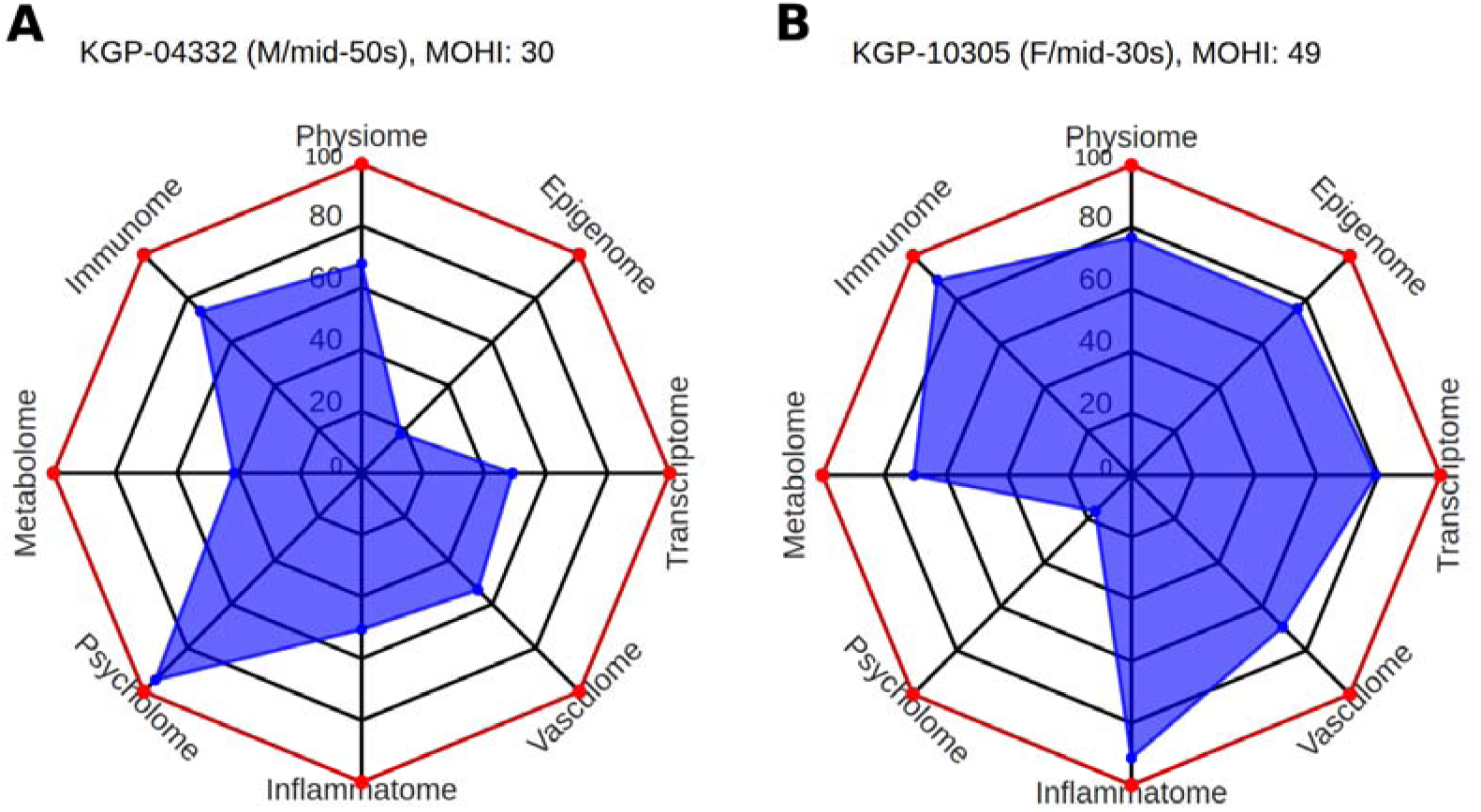
Healthome Polygons depicting the health status of two selected KGP subjects (KGP-04332 and KGP-10305) who owned available datasets in most omes. These two subjects initially had health scores from seven ome-components, excluding Vasculome. Imputation of homocysteine levels based on the corresponding age group and sex resulted in the inclusion of all eight ome-components. Their individual health status was quantified using MOHI scores. The red edges and vertices stand for HPs of PSH subjects. (A) HP of KGP04332: a male in his mid-50s with a MOHI score of 30. (B) HP of KGP-10305: a female in her mid-30s with a MOHI score of 49. KGP, Korean Genome Project; M, Male; F, Female; MOHI, Multi-Omics Health Index

Subsequently, the results of the HP and MOHI for KGP-04332 and KGP-10305 were consistently and reasonably explained using the explainability methods (Figure 4 and Figure 5).

**Figure 4.**
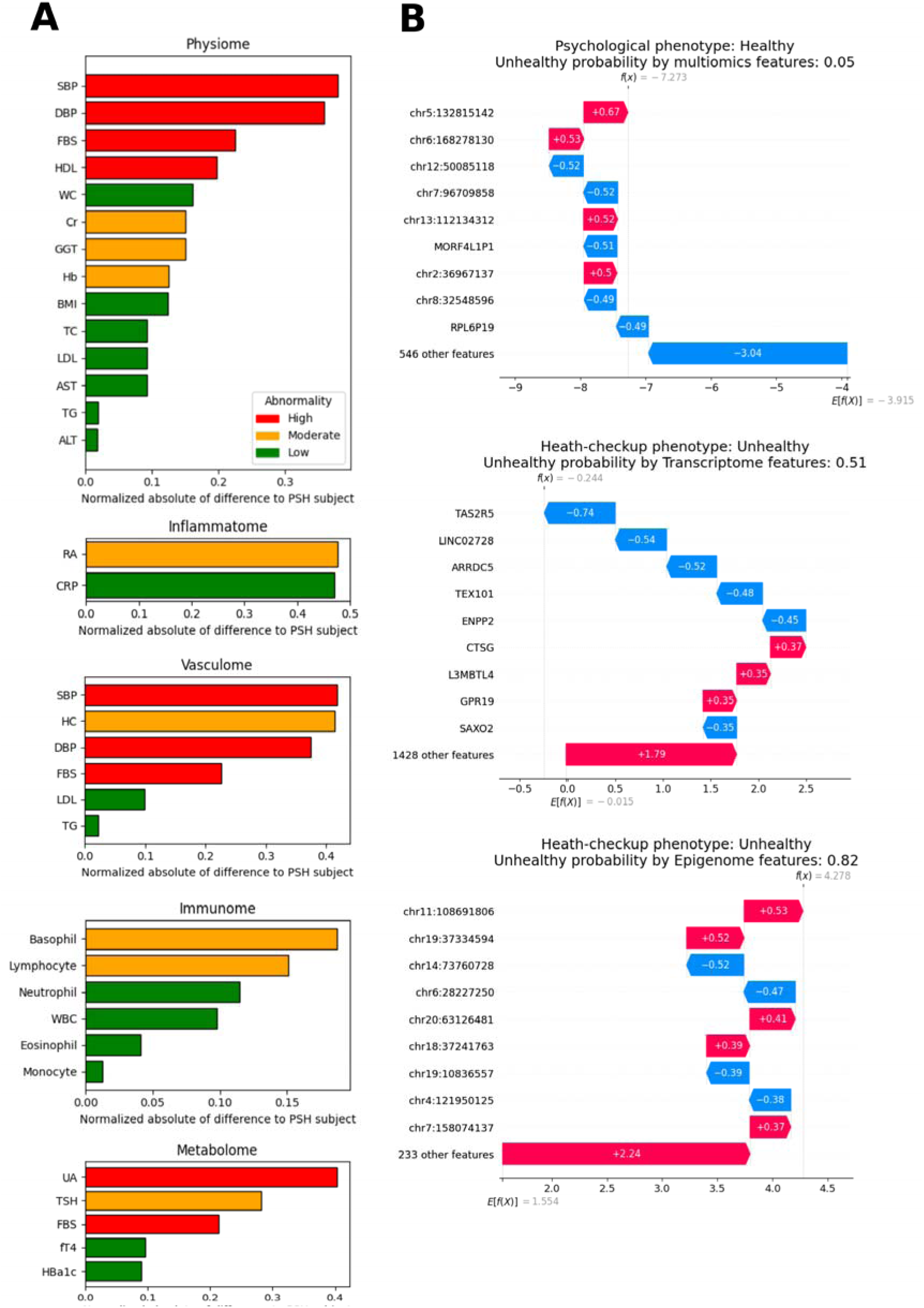
Explainability of the HP chart for KGP-04332 (a male in his mid-50s). (A) Min-max normalized data for each ome-component was subtracted from the corresponding min-max normalized data of the PSH subjects. Threshold values at the 50th and 75th percentiles were then determined. If the normalized data for a subject is less than the 50th percentile, it is classified as low abnormality; data between the 50th and 75th percentiles is classified as moderate abnormality; and data greater than the 75th percentile is classified as high abnormality. Red, yellow, and green bars stand for high, moderate, and low abnormality, respectively. (B) For Psycholome, Transcriptome, and Epigenome, a linear regression model was analyzed by the SHAP algorithm to explain the results using SHAP waterfall plot. The SHAP waterfall plot explains the contribution of each feature to the predicted probability. As a single factor, the most intensive positive and negative factors for unhealthiness were chr5:132815142 and chr12:50085118 CpG sites, CTSG (Cathepsin G) and TAS2R5 (Taste 2 Receptor Member 5) genes, chr11:108691806 and chr14:73760728 CpG sites for Psycholome, Transcriptome, and Epigenome, respectively. HP, Healthome Polygon; PSH, Pseudo Super Healthy; SHAP, SHapley Additive exPlanations; SBP, Systolic Blood Pressure; DBP, Diastolic Blood Pressure; FBS, Fasting Blood Sugar; HDL, High Density Lipoprotein; WC, Waist Circumference; Cr, Creatinine; GGT, Gamma-Glutamyl Transferase; Hb, Hemoglobin; BMI, Body Mass Index; TC, Total Cholesterol; LDL, Low Density Lipoprotein; AST, Aspartate Transaminase; TG, Triglyceride; ALT, Alanine Transaminase; RA, Rheumatoid Factor; CRP, C-Reactive Protein; HC, Homocysteine; WBC, White Blood Cell; UA, Uric Acid; TSH, Thyroid Stimulating Hormone; fT4, Free thyroxine; HBa1c, Glycated Hemoglobin A1c

**Figure 5.**
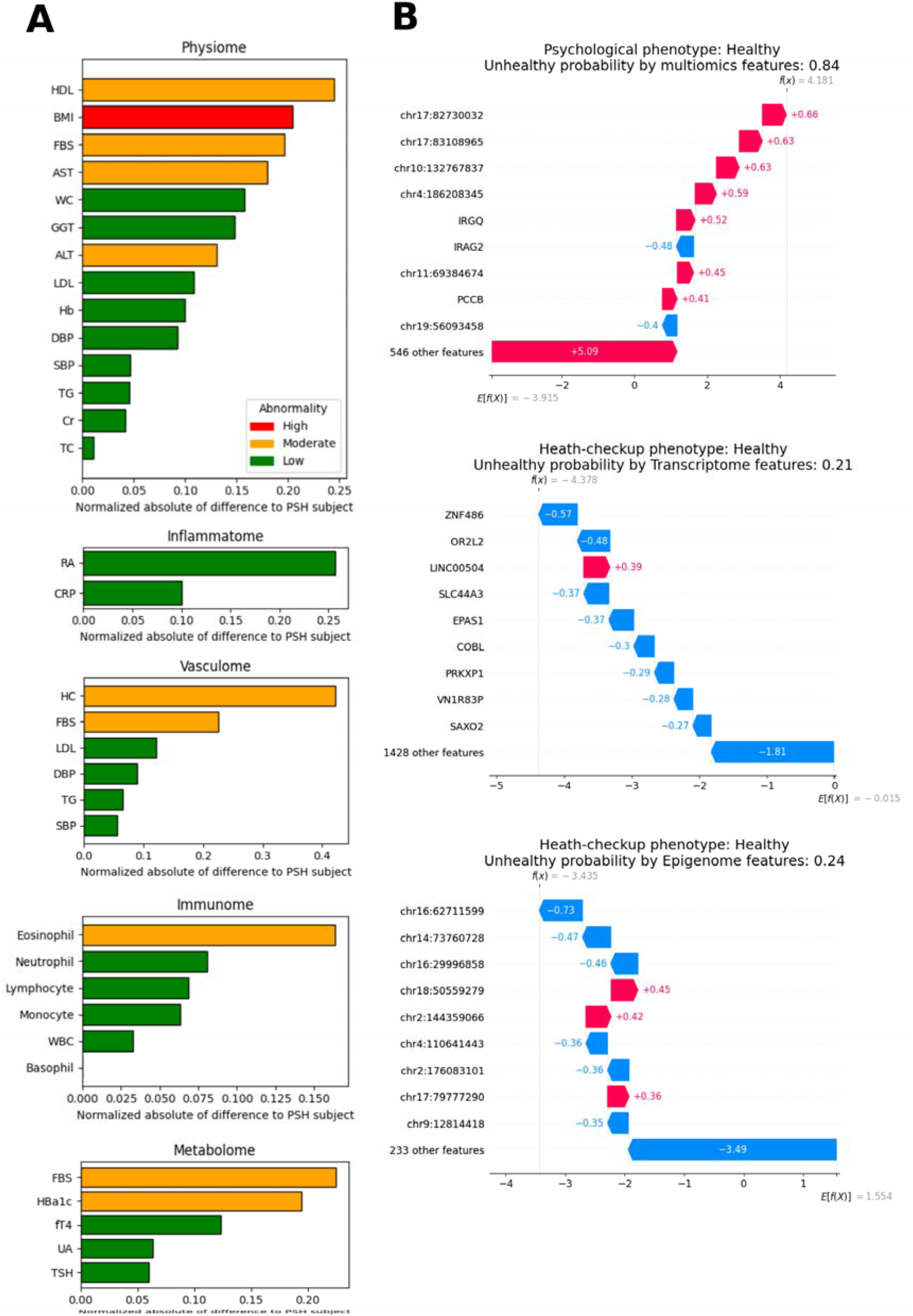
Explainability of the Healthome Polygon chart for KGP-10305 (a female in her mid-30s). (A) Explainability for health scores in five omes. (B) As a single factor, the most intensive positive and negative factors for unhealthiness were chr17:82730032 CpG site and IRAG2 (Inositol 1,4,5-Triphosphate Receptor Associated 2) gene, LINC00504 and ZNF486 (Zinc Finger Protein 486) genes, chr18:50559279 and chr16:62711599 CpG sites for Psycholome, Transcriptome, and Epigenome, respectively.

### Pseudo Super Healthy subjects of five ome-components

After preprocessing the KNHANES dataset to develop the Physiome model, which was used to identify Physiome PSH subjects, 44,541 unhealthy and 39,394 healthy subjects were obtained. The Physiome model trained on this dataset achieved an AUC of 0.92. The Metabolome Thyroid and Metabolome Uric Acid models, which were used to identify Metabolome PSH subjects, demonstrated AUCs of 0.85 and 0.94, respectively, with their corresponding preprocessed datasets comprising 19,936 unhealthy and 15,463 healthy subjects, and 3,375 unhealthy and 2,641 healthy subjects, respectively (Figure 7A).

The HSSH algorithm was then applied to the trained XGBoost models, successfully identifying pseudo super healthy subjects across all sex and age groups. Specific pseudo super healthy subjects were further identified within each sex group (male and female) and each age group (10≤age<20, 20≤age<30, 30≤age<40, 40≤age<50, 50≤age<60, 60≤age<70, and age≥70) (Supplementary Table 3).

Finally, using both the HSSH algorithm and domain knowledge, PSH subjects stratified by sex across all age groups were established for five ome components: Physiome, Metabolome, Vasculome, Immunome, and Inflammatome (Table 2).

**Table 2.**
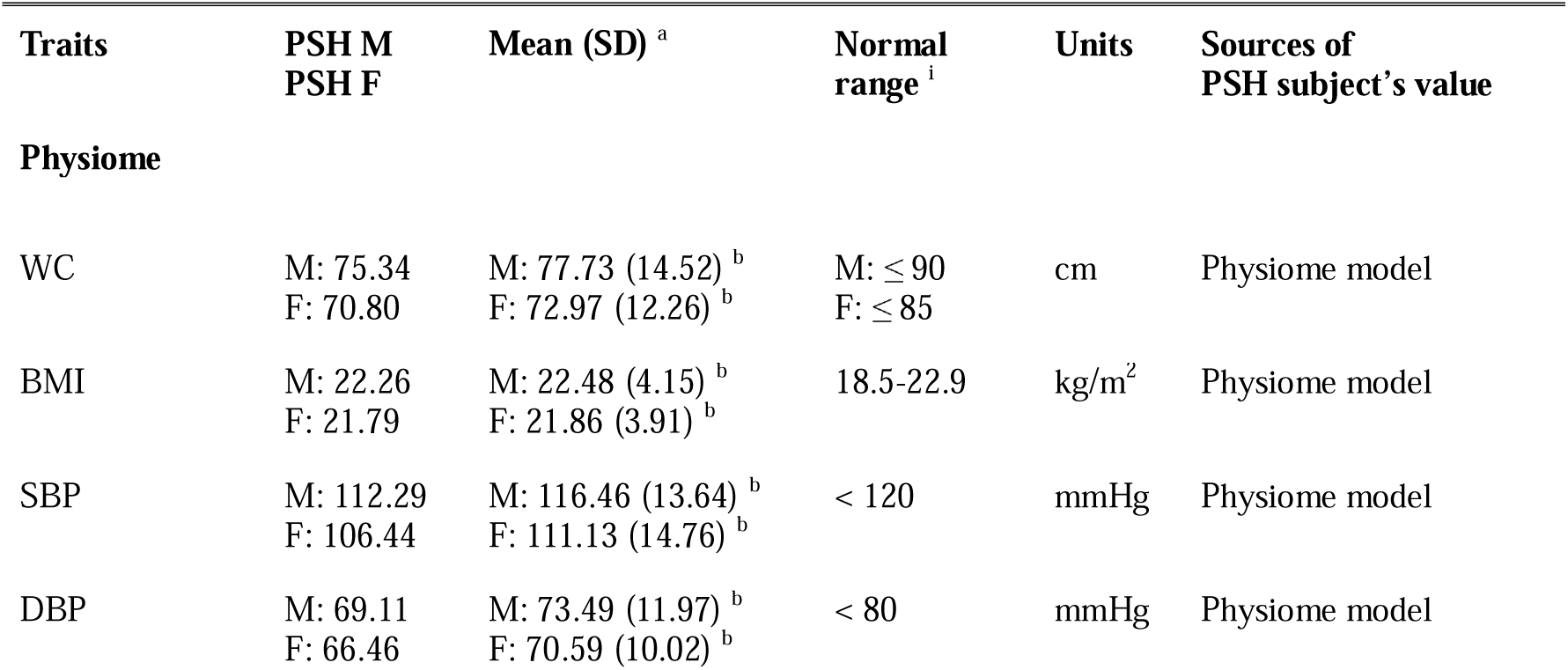
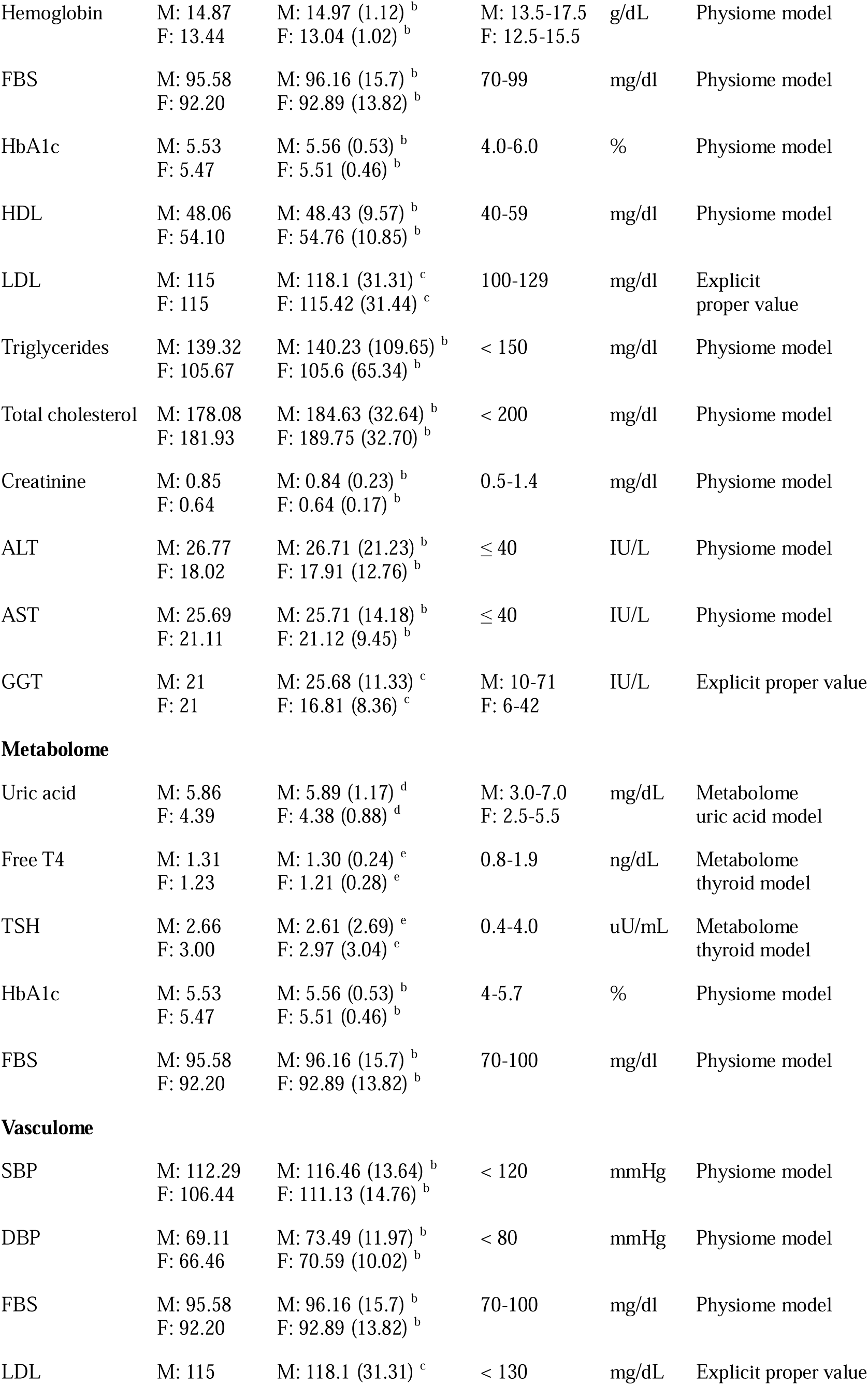
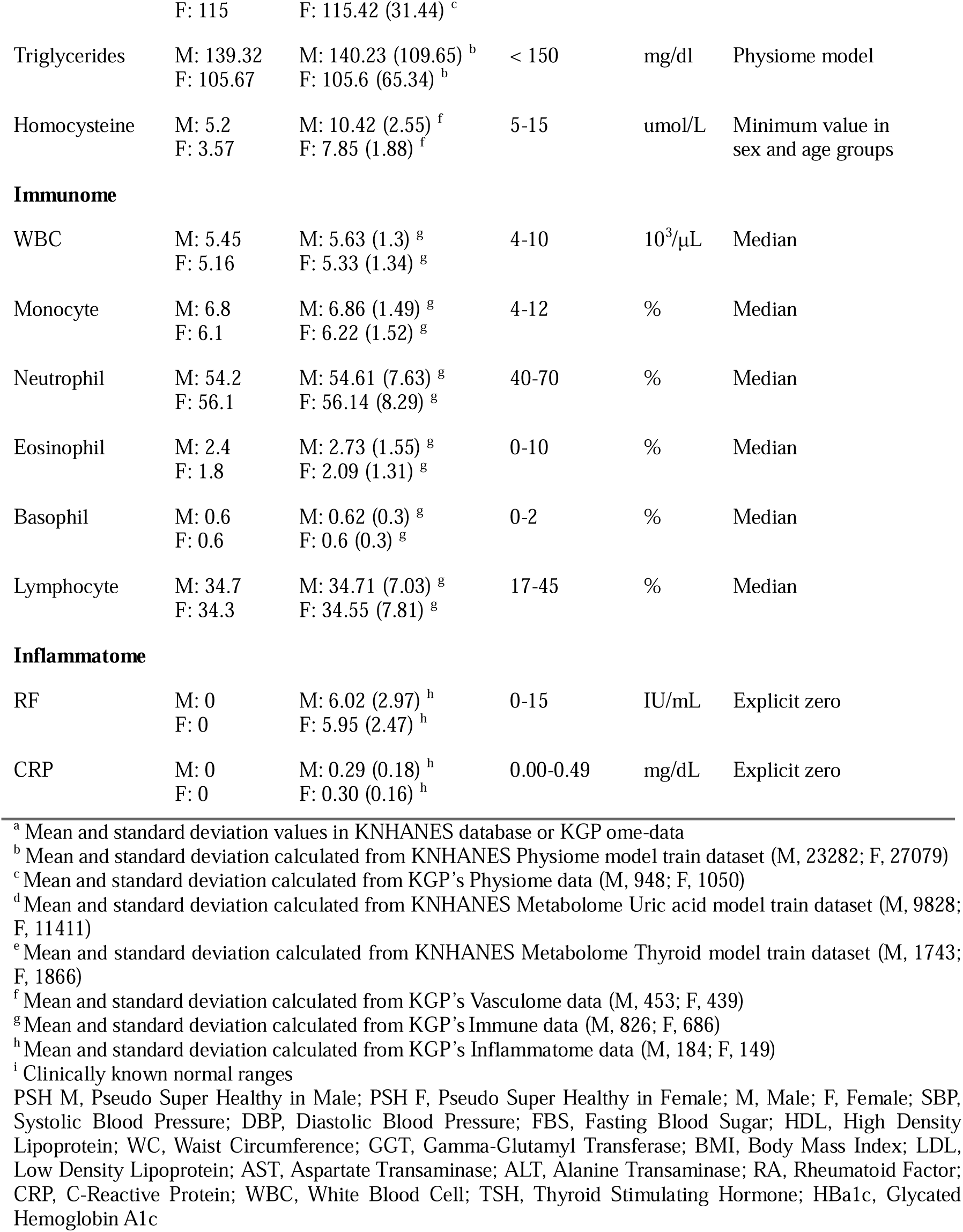
PSH (Pseudo Super Healthy) subjects in male and female groups found by HSSH (Health-check-up data and SHAP-value based pseudo Super Healthy) algorithm.

### Euclidean distances of five ome-components and the validation on them as health indicators

Euclidean distances between PSH and KGP subjects in Physiome, Metabolome, Vasculome, and Immunome showed a bell-shaped distribution, which is ideal in a health quantification system because it indicates frequent values around the mean and rarer values at both ends. On the other hand, Euclidean distances in the Inflammatome showed a large peak, suggesting that trait values of subjects are clustered around the mean, resulting in similar Euclidean distance values from the Inflammatome PSH (Figure 7, Supplementary Figure 4, and Supplementary Table 4).

The histogram of NATs in the Physiome showed a bell-shaped distribution with a mean and standard deviation of 3.48 and 1.74 for males, and 2.79 and 1.69 for females. The Euclidean distances and NATs in the Physiome exhibited moderate positive correlations (male: Pearson’s r = 0.62, *P* < 0.001; female: r = 0.70, *P* < 0.001). This suggests that Euclidean distance can serve as an indicator of health status (Figure 7B and Figure 7C).

In further analysis of the mean ED values of the Physiome by age groups, the values were stable between G0 and G7; however, the mean ED values began increasing at G7 (Figure 6).

**Figure 6.**
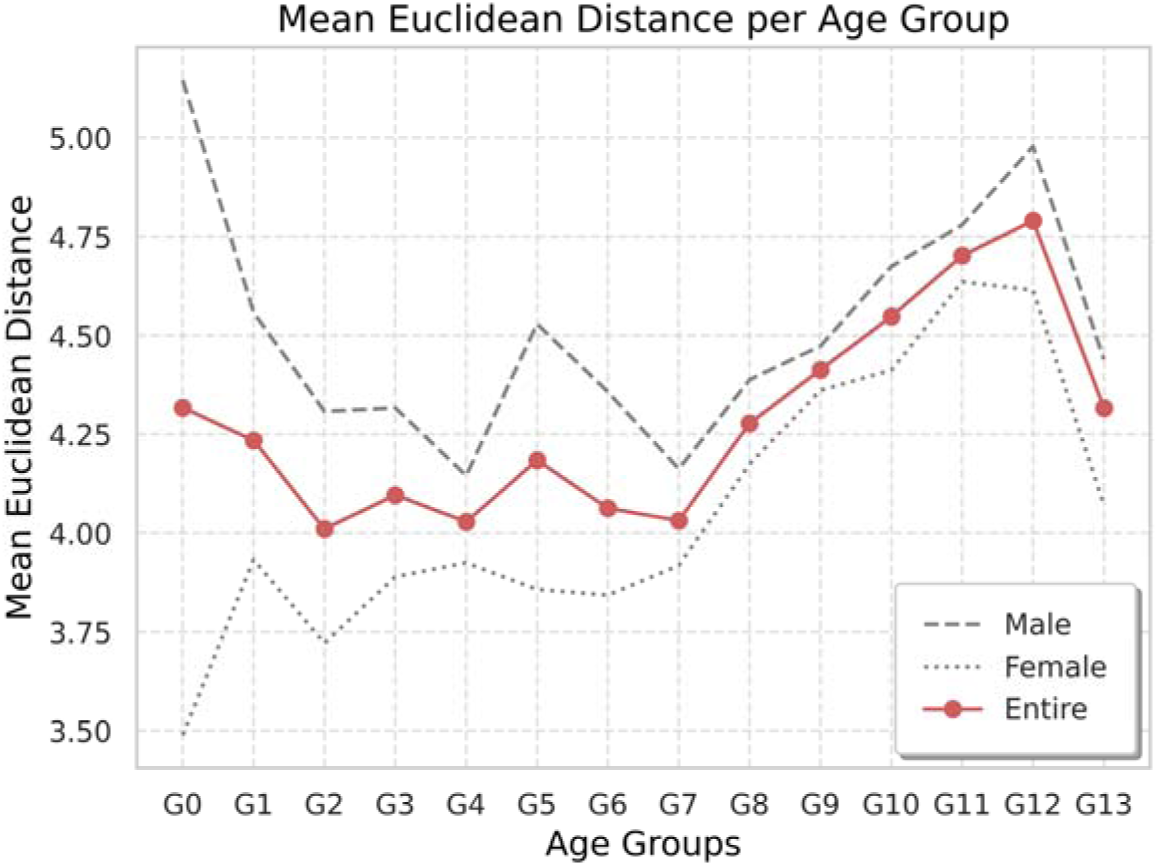
Line plot of the mean Euclidean distance per age group and sex in Physiome. Age groups were defined in 5-year intervals. Age groups are 15 ≤ G0 < 20, 20 ≤ G1 < 25, 25 ≤ G2 < 30, 30 ≤ G3 < 35, 35 ≤ G4 < 40, 40 ≤ G5 < 45, 45 ≤ G6 < 50, 50 ≤ G7 < 55, 55 ≤ G8 < 60, 60 ≤ G9 < 65, 65 ≤ G10 < 70, 70 ≤ G11 < 75, 75 ≤ G12 < 80, 80 ≤ G13 years

**Figure 7.**
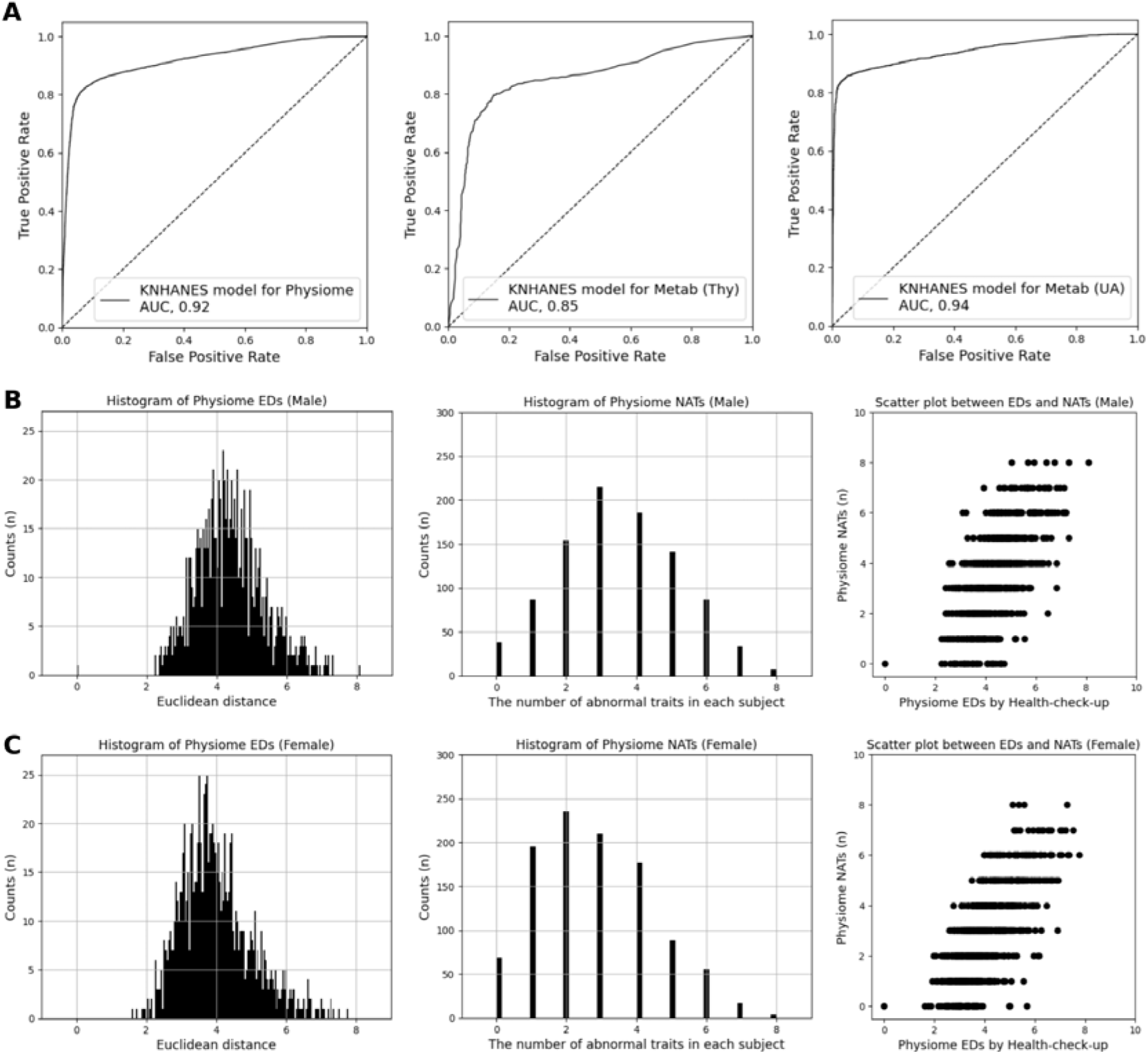
The performances of KNHANES models for Physiome, Metabolome Thyroid, and Metabolome Uric acid as well as the histogram of ED and the validation of ED based on NATs. The performance of KNHANES models which were used to find PSH subjects in Physiome and Metabolome was described as AUC and ROC curve in A. The distribution of Physiome’s ED values by sex is shown and PSH subjects are located at zero location in Euclidean distance (Left of B and C). Additionally, the distribution of Physiome’s NATs by sex is illustrated (Middle of B and C). The scatter plot between ED and NATs demonstrates a positive correlation in visual qualitative evaluation (Right of B and C). This positive correlation was confirmed by Pearson’s correlation analysis, with coefficients of 0.62 and 0.70 and corresponding *p*-values of 0.001 for the male and female groups, respectively. In the scatter plot between EDs and NATs, PSH subjects were shown at (0,0) position. ED, Euclidean distance; NATs, The Number of Abnormal Traits; KNHANES, The Korea National Health and Nutrition Examination Survey

### Psycholome, Transcriptome, and Epigenome health scores using logistic regression models and their evaluation as health indicators

In the transcriptome dataset, a clear batch-effect pattern was observed, associated with the sequencing platform and sequencing year. In contrast, the epigenome dataset, constructed from a single sequencing platform, exhibited only a minor batch effect related to the sequencing year (Supplementary Figure 5).

After a log-likelihood test, which was used to select important features that can maintain high model accuracy, 557 multiomics features were selected, consisting of 327 epigenomic features and 230 transcriptomic features (Supplementary Table 6). The Psycholome model, which was trained with these features to predict the Psycholome health score, showed an AUC of 0.79 [95% CI: 0.72-0.87] (Figure 8B).

**Figure 8.**
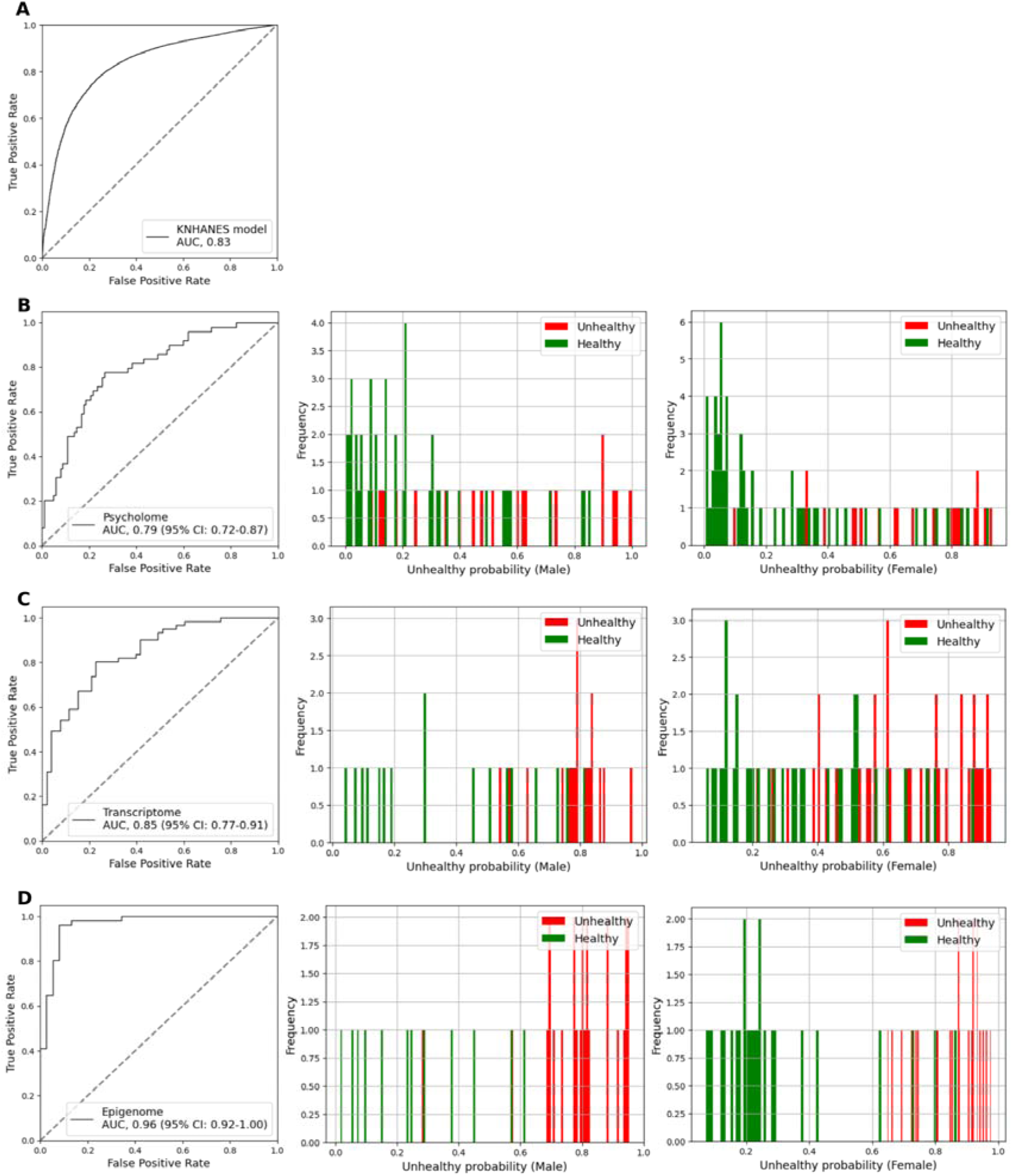
Results of the KNHANES, Psycholome, Transcriptome, and Epigenome models. (A) The KNHANES model was developed to generate pseudo-unhealthy labels for the Transcriptome and Epigenome datasets. The AUC of the model was 0.83 (B–D) The ROC curve and AUC of the models, as well as histograms of predicted unhealthy probability values for males and females, were illustrated for the Psycholome, Transcriptome, and Epigenome models.

The trained KNHANES model, which was used to construct the pseudo-unhealthy label for the Transcriptome and Epigenome of KGP subjects, showed an AUC of 0.83 (Figure 8A). The reasonability of the pseudo-unhealthy labels was validated by the statistical significance of 27 traits from the KGP questionnaire dataset out of a total of 171 traits. Since the pseudo-unhealthy label was generated using many common disease variables in KNHANES, these 27 significant traits were also related to common diseases such as, Hypertension, No circulatory disease, Presbyopia, Hyperlipidemia, No cancer, Hypercholesterolemia, Diabetes mellitus type2, among others (Table 3).

**Table 3.** Chi-square test between pseudo unhealthy labels generated by KNHANES model for Transcriptome and Epigenome datasets and each trait of KGP questionnaire dataset out of 171 health related traits, ascending sorted by adjusted *p*-value. The number of samples in this test is male 1048 and female 1146. The following 27 traits of KGP questionnaire dataset, which are highly associated with common diseases, e.g., hypertension, hyperlipidemia, and diabetes mellitus type2, were statistically significant to pseudo unhealthy labels.

| Traits | $\chi^2$ statistic | *p*-value | adjusted *p*-value |
| --- | --- | --- | --- |
| Hypertension | 177.13 | 2.05E-40 | 2.95E-38 |
| No circulatory disease | 152.32 | 5.39E-35 | 3.88E-33 |
| Presbyopia | 130.20 | 3.70E-30 | 1.78E-28 |
| Hyperlipidemia | 100.79 | 1.02E-23 | 3.68E-22 |
| Colon polyps | 81.54 | 1.72E-19 | 4.94E-18 |
| No cancer | 76.84 | 1.85E-18 | 4.44E-17 |
| Hypercholesterolemia | 53.75 | 2.28E-13 | 4.69E-12 |
| Age-related cataract | 48.42 | 3.45E-12 | 6.21E-11 |
| Myopia | 45.41 | 1.60E-11 | 2.56E-10 |
| Diabetes mellitus type2 | 37.39 | 9.66E-10 | 1.39E-08 |
| No metabolic disease | 26.79 | 2.27E-07 | 2.97E-06 |
| Dry eye syndrome | 20.33 | 6.52E-06 | 7.67E-05 |
| Allergic rhinitis | 20.22 | 6.92E-06 | 7.67E-05 |
| Astigmatism | 19.97 | 7.88E-06 | 8.10E-05 |
| PolyCystic Ovary Syndrome | 18.72 | 1.51E-05 | 1.45E-04 |
| Age-related hearing loss | 15.13 | 1.00E-04 | 9.02E-04 |
| Angina | 14.43 | 1.45E-04 | 1.23E-03 |
| Gout | 12.90 | 3.28E-04 | 2.62E-03 |
| Hemorrhoids | 11.24 | 8.02E-04 | 6.08E-03 |
| Hyperopia | 9.34 | 2.24E-03 | 1.61E-02 |
| Obesity | 9.06 | 2.61E-03 | 1.79E-02 |
| Sleep disorder | 8.61 | 3.35E-03 | 2.19E-02 |
| No blood disease | 8.50 | 3.56E-03 | 2.23E-02 |
| Iron deficiency anemia | 8.00 | 4.67E-03 | 2.80E-02 |
| Chronic sinusitis | 7.73 | 5.43E-03 | 3.13E-02 |
| Tinnitus | 7.06 | 7.88E-03 | 4.20E-02 |
| Bell's palsy | 7.07 | 7.85E-03 | 4.20E-02 |

For the Transcriptome model, which was used for predicting Transcriptome health scores, the log-likelihood test selected 1,437 important transcript features (Supplementary Table 7). The Transcriptome model showed an AUC of 0.85 [95% CI: 0.77-0.91] and a balanced probability distribution, which is ideal for a health quantification framework (Figure 8C).

For the Epigenome model, which was used for predicting Epigenome health scores, the log-likelihood test selected 242 important epigenomic features (Supplementary Table 8). This Epigenome model showed high accuracy with an AUC of 0.96 [95% CI: 0.92-1.00] and a balanced health probability as well (Figure 8D).

## Discussions

### Limitations and challenges with the Healthome Polygon framework

While the Healthome Polygon framework represents a significant advancement in multi-dimensional health quantification, several limitations and challenges must be acknowledged.

First, the identification of PSH subjects for the Physiome and Metabolome was achieved using data-driven AI algorithms, which leverage large-scale datasets to define optimal health. However, for other “omes”—such as the Vasculome, Immunome, and Inflammatome—PSH subjects were determined based on domain knowledge rather than data-driven approaches. This reliance on expert judgment was necessitated by the lack of relevant feature data in the KNHANES, which prevented the development of robust AI models for these domains. This limitation underscores the importance of data-driven approaches for defining PSH and highlights the need for more comprehensive datasets to enable AI-based modeling across all “omes.”

Second, the unhealthiness labels used for developing the Transcriptome and Epigenome models were not directly derived from clinical or biological measurements. Instead, these labels were pseudo-unhealthiness labels predicted by a trained model using the KNHANES database, similar to the approach used for generating Physiome labels. While this method allowed for the integration of Transcriptome and Epigenome data into the framework, it introduces potential biases and uncertainties, as the labels may not fully capture the true biological state of these “omes.” Future studies should aim to incorporate direct measurements or more robust biomarkers to improve the accuracy of these models.

Third, the supervised models trained on the KNHANES database are skewed toward common diseases, such as hypertension, dyslipidemia, and diabetes, rather than evenly reflecting the full spectrum of disease occurrences. This bias stems from the overrepresentation of these prevalent conditions among the subjects included in the dataset. Furthermore, the construction of unhealthy labels in the KNHANES database, which relies on aggregating “yes” responses across three sources of disease information—diagnoses by medical professionals, self-reported current disease status, and health examination results—has resulted in a disproportionately high ratio of unhealthy labels. This simplistic approach may not adequately capture the nuanced health status of individuals, highlighting the need for more detailed and stringent criteria for unhealthy labeling.

Fourth, the KNHANES dataset is exclusively composed of Korean subjects, limiting its generalizability to other ethnic groups due to the absence of diverse biological patterns. As a result, the performance of models trained on this dataset cannot be guaranteed for populations with different genetic and environmental backgrounds.[29]

Despite these limitations, the Healthome Polygon framework offers a scalable and adaptable approach to health quantification. The challenges identified here can be addressed through the collection of more comprehensive datasets, the development of advanced AI models, and the integration of direct biological measurements. These improvements will further enhance the accuracy, reliability, and generalizability of the framework, paving the way for its broader adoption in personalized medicine and healthcare management.

To address these limitations, we propose the development of Diseasome Polygon, which incorporates specific disease labels to move beyond the traditional disease-centric model. Notably, this framework will be expanded to include associations with cancers, providing a more comprehensive understanding of disease dynamics. Furthermore, both the Healthome and Diseasome Polygons will be developed using multiomics data diverse populations across multiple countries, establishing a multiethnic health quantification framework. By leveraging the well-defined concept of PSH subjects, these limitations can be effectively mitigated, enabling a more accurate and globally applicable health assessment system.

### Motivation and necessity of the Healthome Polygon framework

The development of the MOHI and HP is motivated by the growing recognition that health is a dynamic and multifactorial condition influenced by genetics, lifestyle, environment, and medical history. Traditional health monitoring systems often focus on isolated risk factors or single disease outcomes, failing to capture the complex interactions that define an individual’s overall health. By integrating multiomics data, MOHI provides a powerful tool for quantifying these complex interactions, offering insights into both current health and future risks. This approach represents a paradigm shift from reactive disease management to proactive and preventative health monitoring, aligning with the principles of personalized medicine.

### Justification and key benefits of the multiomics-based Healthome Polygon framework

The adoption of a multiomics-based health assessment model is further justified by the increasing availability of large-scale health data repositories, such as the KGP and the KNHANES. These datasets enable the generation of the health indices based on diverse populations, ensuring the applicability of the HP framework across different demographic groups. Moreover, the integration of machine learning techniques allows for the identification of patterns and correlations across multiple biological layers, advancing our understanding of how different omics domains interact to influence health outcomes.

One of the key benefits of the HP framework is its potential to personalize health assessments and interventions. Unlike traditional health assessments, which often provide generalized recommendations, the MOHI enables a more individualized approach. And individuals can make an effort to enhance health by changing unhealthy omes. Therefore, the composition of the eight “omes” was defined as modifiable components rather than almost non-modifiable ones like the Genome, because the Transcriptome,[30,31] Epigenome,[30,31] Psycholome,[32–34] and general health check-up traits composing the Physiome,[35–37] Metabolome,[30,31,36] Inflammatome,[38] Immunome,[39] and Vasculome[40] are known to be changeable.

By evaluating an individual’s health status in relation to the PSH reference group, healthcare providers can identify specific areas of risk or opportunities for improvement. This capability holds significant promise for early disease detection, as subtle deviations in omics data—such as changes in gene expression or metabolite levels—can be detected before clinical symptoms manifest. Additionally, the integration of omics data with clinical measurements facilitates the prediction of disease progression, the tailoring of preventative strategies, and the real-time monitoring of treatment effectiveness.

### Addressing age, a potential bias in Euclidean distance indicator

Initially, age was considered as a trait for calculating Euclidean distance due to its established role as a risk factor for aging-related diseases such as neurodegenerative, metabolic, and cardiovascular diseases.[41,42] However, Pearson’s correlation analysis related that the Euclidean distance calculated without including age was not strongly correlated with age. For example, in a dataset of 949 male subjects (mean age: 45.4 ± 14.5 years), the PCC (Pearson’s correlation coefficient) was 0.075 (*p*-value = 0.022) between age and Euclidean distance (mean: 4.4 ± 1.0). Similarly, for 1,051 female subjects (mean age: 45.4 ± 14.1 years), the correlation coefficient was 0.197 (*p*-value < 0.01) between age and Euclidean distance (mean: 4.0 ± 1.0) (Supplementary Figure 6B). These findings suggest that age can introduce bias into the ED calculation. For instance, a subject with an age similar to that age of PSH subjects but unhealthy values in other traits may incorrectly appear healthier due to this age bias. Consequently, age was excluded from Euclidean distance calculation. However, age remains a relevant factor in health assessment and can be considered from two perspectives.

First, as datasets grow larger, age may become a more useful feature for describing health with great precision. As evidence of this, our analysis showed that correlation coefficients exhibited an increasing pattern in older age groups (PCC_male_young_ = -0.128, PCC_female_young_ = 0.028, PCC_male_middle_ = -0.068, PCC_female_middle_ = 0.109, PCC_male_old_ = 0.146, PCC_female_old_ = 0.117, and their corresponding *p*-values are 0.015, 0.588, 0.168, 0.017, 0.055, 0.111, respectively) (Supplementary Figure 6C to Supplementary Figure 6E). Additionally, in the plot of the mean Physiome ED per age group and sex, an increase in ED began at age group 7 (50–55 years; mean age of subjects in this group: 52.05 ± 1.41 years; M: 110, F: 127) (Figure 6), based on the largest slope value of 0.246 between age groups 7 and 8, except for the slope value between age group 12 and age group 13 (all slope values are in Supplementary Table 10). Similar to this result, a previous study reported two inflection points in aging, at which mortality risk and aging-associated diseases accelerate—at 44 years and 60 years.[43] On average, the mean Physiome ED was higher in males than in females across all age groups. The high mean ED in male age group 0 is likely due to sample bias (n = 2; detailed clinical values are in Supplementary Table 11) or fluctuations in aging characteristics at this young age. For example, the previous study reported all omics exhibited high variability in the child cohort, with gene expression being the least stable.[44] Consequently, the reliability of this young age group can be reduced, and aging omics patterns become more stable after this developmental stage, enhancing the reliability of the health quantification framework.

Interestingly, age group 13 (80–85 years) exhibited a lower ED. This may be attributed to the fact that individuals in this group who participated in the Korea10K project—which involves blood sampling, health check-ups, and health questionnaires—represent a selectively healthier population that has survived age-related mortality, introducing a potential sample bias (n = 4 for males and n = 2 for females; detailed clinical values are in Supplementary Table 12). Therefore, in further studies, elderly subjects in age group 13 should have their telomere length examined, as several studies have reported a link among telomere length, health, and longevity. Additionally, to better reflect aging patterns, incorporating traits highly associated with aging, such as muscle mass or sarcopenia status, into the Euclidean distance calculation will be helpful.[45,46]

Second, age can be interpreted as an outcome of health quantification analysis—such as in the Healthome Polygon framework—rather than as an input feature. This allows the quantified health score to reflect biological age rather than chronological age. Aging is a dynamic process. The conventional perspective assumed that aging follows a fixed pattern, meaning chronological age directly determines the level of aging. However, the aging mechanism is a physiological and mechanical process that results in different aging levels than those expected from chronological age.[47–49] Viewing aging in this way could transform the principle of health management.

To measure aging precisely, a health quantification framework like this is necessary. The above ED values are derived from the Physiome model. However, incorporating additional omes would create a more robust tool for assessing health or biological aging, making it a more absolute measure than relative characteristic chronological age. As a result, interpreting age as a quantifiable health outcome aligns with the ultimate goal of capturing the dynamic and multifactorial nature of health by integrating multidimensional input features within the health quantification framework.

### Dataset collection from the medical institutes for future health application

The KGP dataset, constructed by collecting data from multiple hospitals and institutes, exhibits variability in data traits and a high ratio of missing values. These challenges hinder the development of health monitoring applications, such as the HP. For example, when constructing the HP with eight omes, the number of applicable subjects was drastically reduced due to missing data (Supplementary Figure 8). To address this issue, future studies should focus on standardizing health traits and conducting health examinations using these standardized protocols across medical institutes. Such efforts will enhance data quality and facilitate the development of robust health monitoring applications.

## Conclusion

The HP framework represents a transformative approach to health quantification, addressing the limitations of traditional models by integrating multiomics data and leveraging artificial intelligence. By developing the Healthome Polygon and expanding the framework to include multiethnic data, we aim to create a globally applicable system for health assessment and disease monitoring. The exclusion of age as a bias in Euclidean distance calculations and the emphasis on standardized data collection further enhance the framework’s accuracy and reliability. Ultimately, the HP framework holds significant promise for advancing personalized medicine, enabling early disease detection, and improving healthcare management on a global scale.

## Funding

This work was supported by the U-K BRAND Research Fund (1.200108.01) of UNIST (Ulsan National Institute of Science & Technology). This work was also supported by the Research Project Funded by Ulsan City Research Fund (1.200047.01) of UNIST (Ulsan National Institute of Science & Technology). This work was also supported by the Promotion of Innovative Businesses for Regulation-Free Special Zones funded by the Ministry of SMEs and Startups (MSS, Korea) (Grant number [P0016195, P0016193] (1425156792, 1425157301) (2.220035.01, 2.220036.01)). This work was also supported by the Establishment of Demonstration Infrastructure for Regulation-Free Special Zones fund (MSS, Korea) (Grant number [P0016191] (2.220037.01) (1425157253)) by the Ministry of SMEs and Startups. Ulsan university hospital biobank provided DNA sample and clinical data from 696 participants (60SA2017002-006-4, 60SA2016001-010-5).

## Acknowledgements

We extend our gratitude to all participants and the citizens of Ulsan for their support of the Korean Genome Project, which provided the foundation for this study. This research was made possible through the Biodatafarm computing infrastructure, funded by the Ulsan Metropolitan City Government. We appreciate the colleagues at KOGIC (Korean Genomics Center) and GenomeLab of UNIST (Ulsan National Institute of Science and Technology) for their valuable discussions on this study. We thank all honest and purpose-oriented scientists who made scientific bases and devoted their lives in enhancing syntropy of the world.

## Ethics declarations

Sample collection and sequencing were approved by the Institutional Review Board (IRB) of the Ulsan National Institute of Science and Technology (UNISTIRB-15-19-A and UNISTIRB-16-13-C). The data employed in our study originates from voluntary blood or saliva donations, and we have diligently secured explicit, comprehensive consent forms from all participants prior to sample collection. These consent forms explicitly outline the intended use of their data for research purposes and underscore the voluntary nature of their participation. Furthermore, our study adheres to the ethical guidelines and regulations stipulated by the IRB. As a result, we can make the data of 3,839 individuals publicly available while respecting the privacy and consent of the participants. KGP-04332 and KGP-10305 are anonymized IDs, and they were not known to anyone outside the research group.

KNHANES IV (2007-2009), KNHANES V (2010–2012), KNHANES VI (2013–2015), KNHANES VII (2016-2018), and KNHANES VIII (2019-2021) were approved by the KCDC research ethics committee (approval numbers; 2009-01CON-03-2C, 2010-02CON-21-C, 2011-02CON06-C, 2012-01EXP-01-2C, 2013-07CON-03-4C, 2013-12EXP-03-5C, 2018-01-03-P-A, 2018-01-03-C-A, 2018-01-03-2C-A, 2018-01-03-5C-A), and all participants provided written informed consent for participation in KNHANES.

## Conflict of interest

J.B. is the founder of AgingLab. S.J. is the CEO of Geromics, Inc. and CEO of AgingLab, and an employee at Clinomics, Inc.

## Author contributions

J.B. generated, supervised, and edited the manuscript on healthomics, health polygon, and multiomics domains integration to quantify human health for computers. Y.B. acquired the KGP and KNHANES datasets for the development of the Healthome Polygon framework and conducted data analysis and interpretation. Y.B. performed statistical analyses and model development for this study. Y.B. drafted and completed the manuscript. J.B., S.J., and E-S.S. made reviews of the manuscript. J.B. and S.J. supervised this study. S.J. and J.B. contributed equally as co-corresponding authors.

## Supplementary material

supplement.docx

## Data availability

The Korean Genome Project (Korea1K, Korea4K and Korea10K) data are available for sharing. Please contact the corresponding authors or visit the study’s official page at https://koreangenome.org/Healthome_polygon for more information. The KNHANES database can be accessed through the KNHANES official page at https://knhanes.kdca.go.kr/knhanes/eng/index.do, managed by the Korea Disease Control and Prevention Agency.

## Notes

### Author Declarations

Sample collection and sequencing were approved by the Institutional Review Board (IRB) of the Ulsan National Institute of Science and Technology (UNISTIRB-15-19-A and UNISTIRB-16-13-C). The data employed in our study originates from voluntary blood or saliva donations, and we have diligently secured explicit, comprehensive consent forms from all participants prior to sample collection. These consent forms explicitly outline the intended use of their data for research purposes and underscore the voluntary nature of their participation. Furthermore, our study adheres to the ethical guidelines and regulations stipulated by the IRB. As a result, we can make the data of 3,839 individuals publicly available while respecting the privacy and consent of the participants. KGP-04332 and KGP-10305 are anonymized IDs, and they were not known to anyone outside the research group. KNHANES IV (2007-2009), KNHANES V (2010-2012), KNHANES VI (2013-2015), KNHANES VII (2016-2018), and KNHANES VIII (2019-2021) were approved by the KCDC research ethics committee (approval numbers; 2009-01CON-03-2C, 2010-02CON-21-C, 2011-02CON06-C, 2012-01EXP-01-2C, 2013-07CON-03-4C, 2013-12EXP-03-5C, 2018-01-03-P-A, 2018-01-03-C-A, 2018-01-03-2C-A, 2018-01-03-5C-A), and all participants provided written informed consent for participation in KNHANES.

