## Supplementray Figures, Tables, Method for "Healthome Polygon Framework: Comprehensive and Multi-dimensional Health Quantification Framework Using Artificial Intelligence and Multiomics Data"

### **Supplementary Figures**

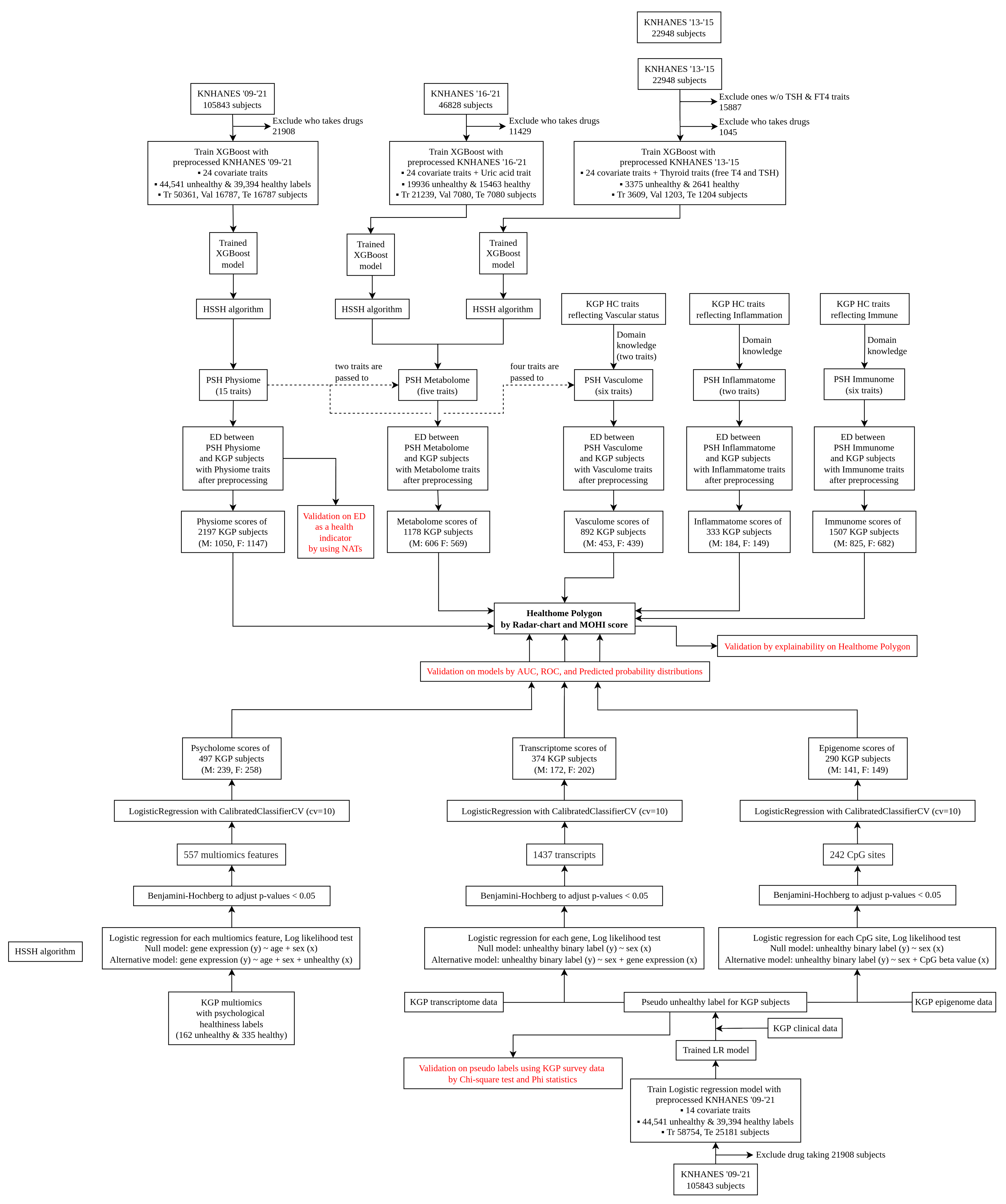

Supplementary Figure 1. Flow of implementation for Healthome Polygon

XGBoost, eXtreme Gradient Boosting; KNHANES, The Korea National Health and Nutrition Examination Survey; Tr, Train set; Val, Validation set; Te, Test set; HSSH, Health-check-up data and SHAP-value based pseudo Super Healthy; PSH, Pseudo Super Healthy; ED, Euclidean Distance; M, Male; F, Female; NATs, The Number of Abnormal Traits; HC, Health-Check-up; MOHI, Multi-Omics Health Index; cv, Cross Validation

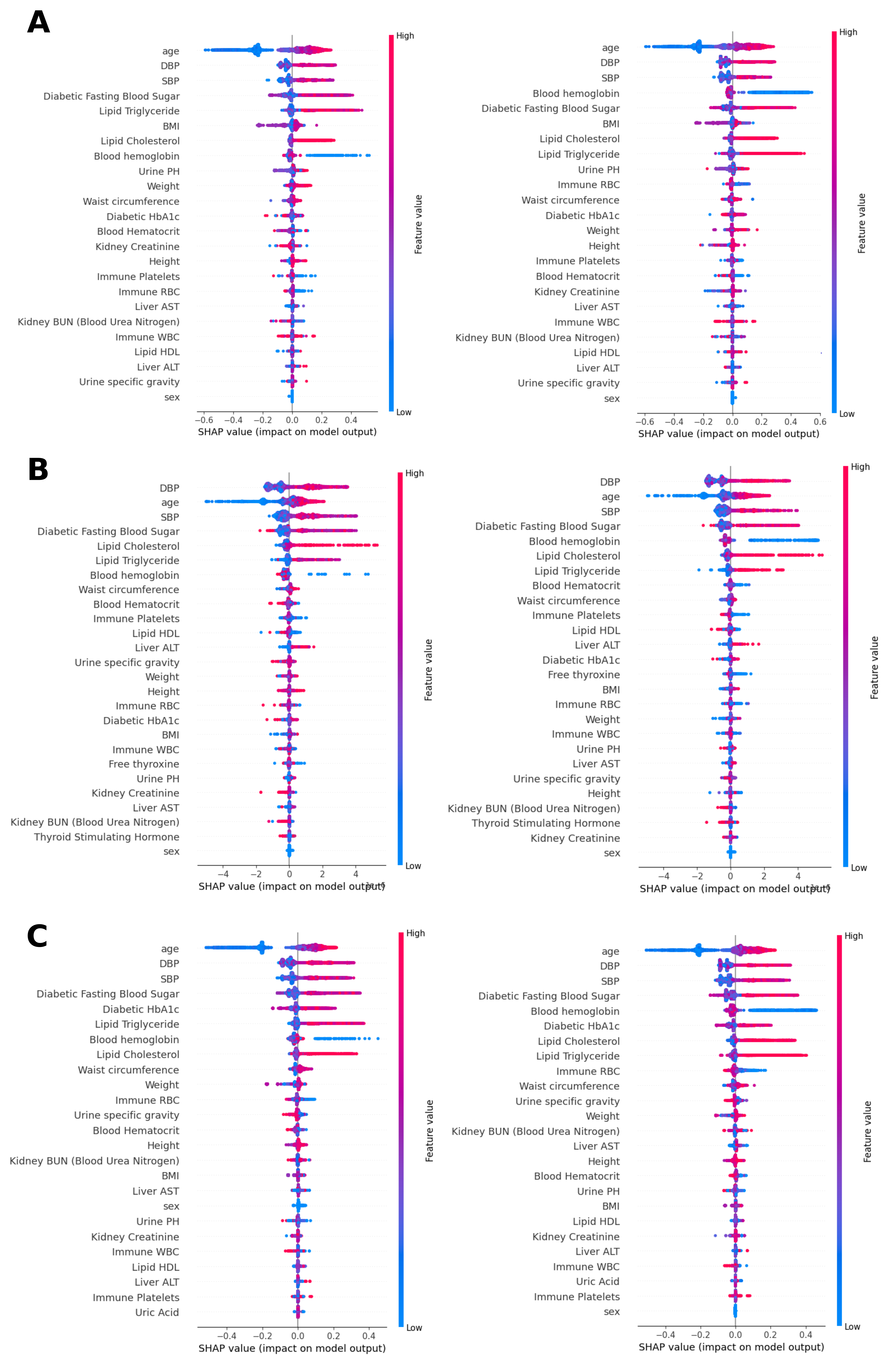

Supplementary Figure 2. Inspect contribution patterns of traits to unhealthy based on SHAP summary plots, which are information that is used for identifying PSH. (A) Male (Left) and female (right) in Physiome model, (B) Male (Left) and female (right) in Metabolome Thyroid model, (C) Male (Left) and female (right) in Metabolome Uric acid model.

SHAP, SHapley Additive exPlanations; PSH, Pseudo Super Healthy

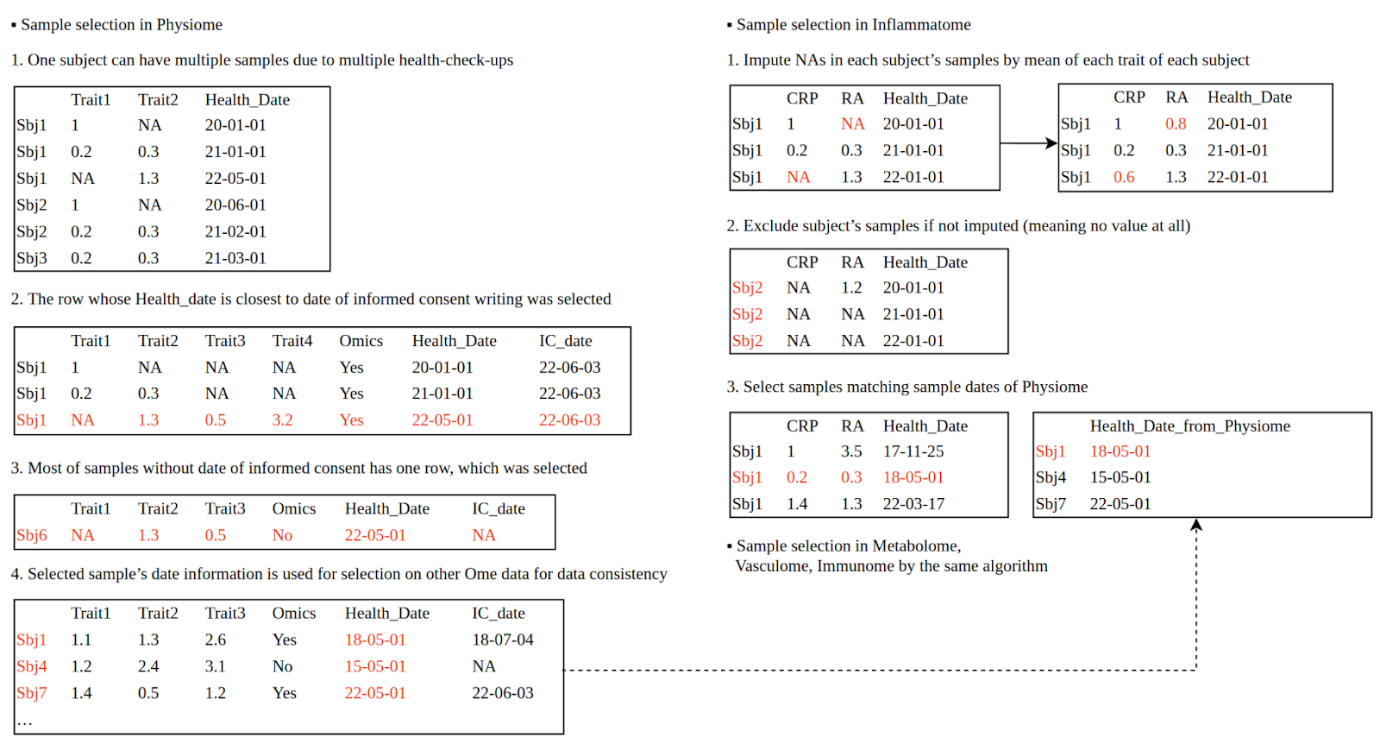

Supplementary Figure 3. Explanation of sample selection algorithm for five ome-components through the virtual example

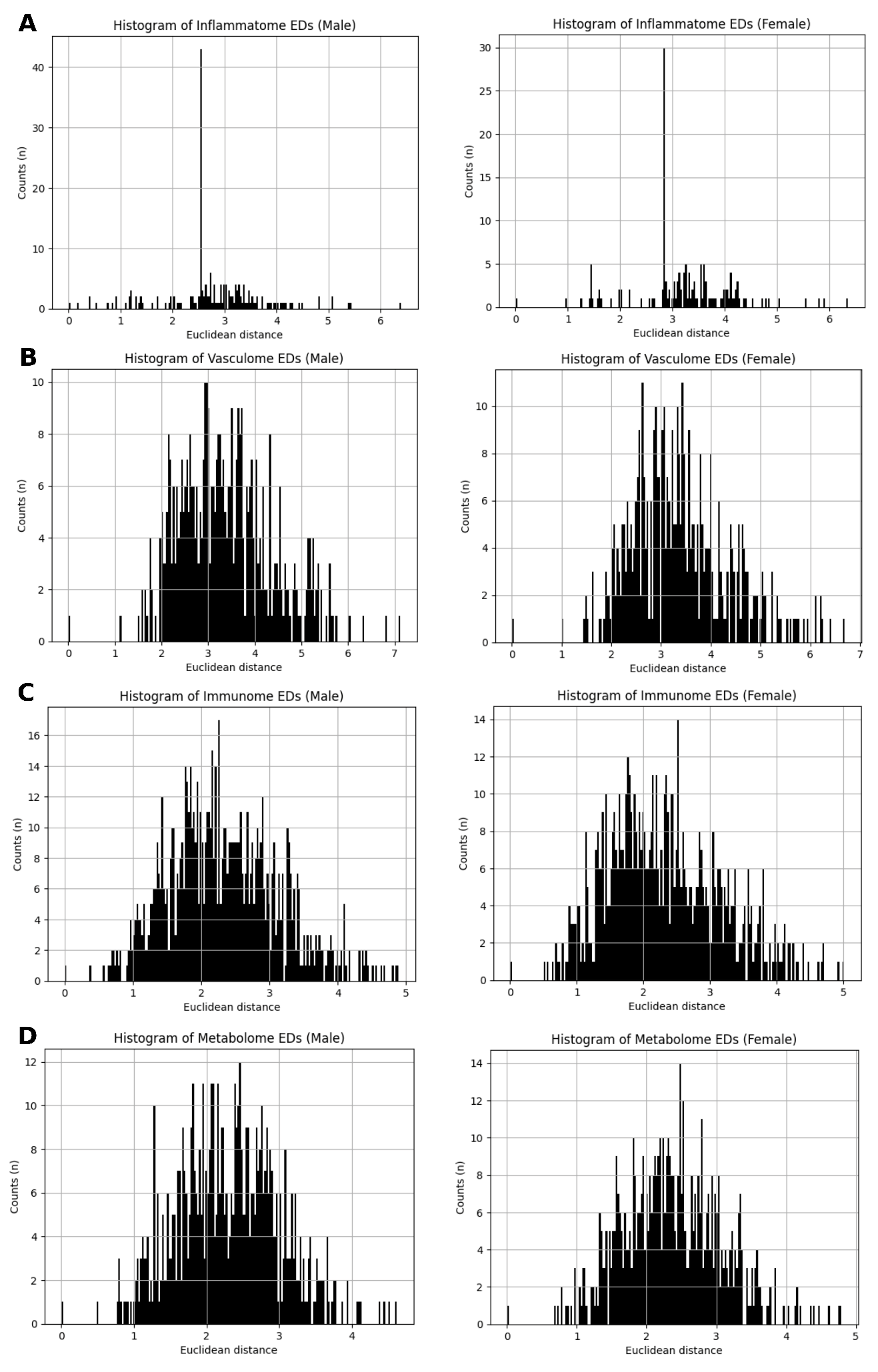

Supplementary Figure 4. The histogram of EDs in the four ome-components. (A) Histogram of Inflammatome EDs. Mean, std, min, and max were 2.77, 0.97, 0.17, 6.4 and 3.2, 0.95, 0.98, 6.35 for male and female, (B) Histogram of Vasculome EDs. Mean, std, min, and max were 3.38, 1, 1.12, 7.12 and 3.37, 0.99, 1.01, 6.7 for male and female, (C) Histogram of Immunome EDs. Mean, std, min, and max were 2.33, 0.81, 0.39, 4.89 and 2.31, 0.85, 0.52, 5.0 for male and female, (D) Histogram of Metabolome EDs. Mean, std, min, and max were 2.37, 0.71, 0.69, 4.8 and 2.37, 0.71, 0.69, 4.8 for male and female. PSH subjects of male and female were shown at ED=0.

ED, Euclidean distance; PSH, Pseudo Super Healthy

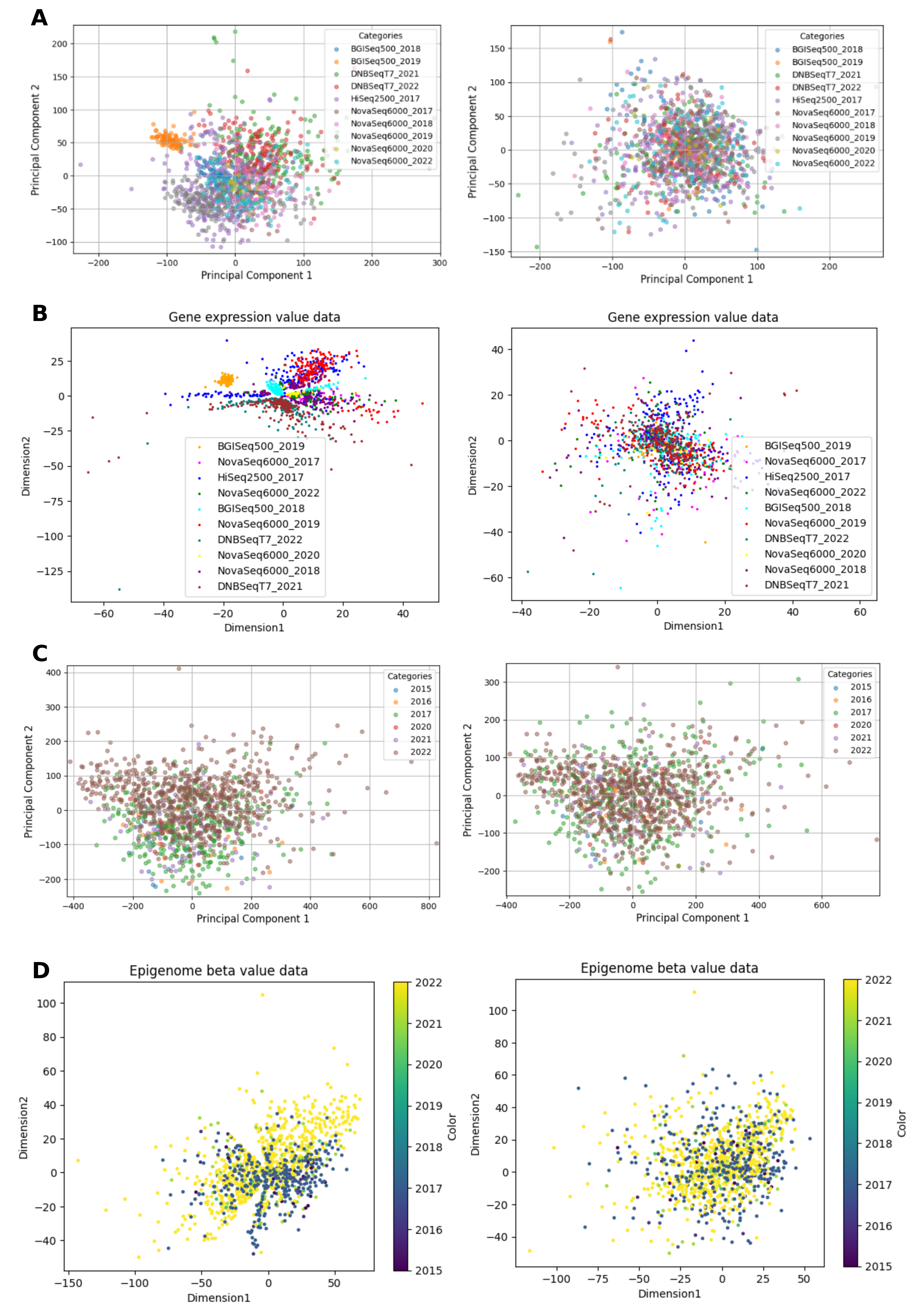

Supplementary Figure 5. Visually confirmation of batch effect correction. (A) Data visualization by PCA before (left) and after (right) batch-effect removal in Transcriptome data (B) Data visualization by deep learning-based Autoencoder before (left) and after (right) batch-effect removal in Transcriptome data (C) Data visualization by PCA before (left) and after (right) batch-effect removal in Epigenome data (D) Data visualization by deep learning-based Autoencoder before (left) and after (right) batch-effect removal in Epigenome data.

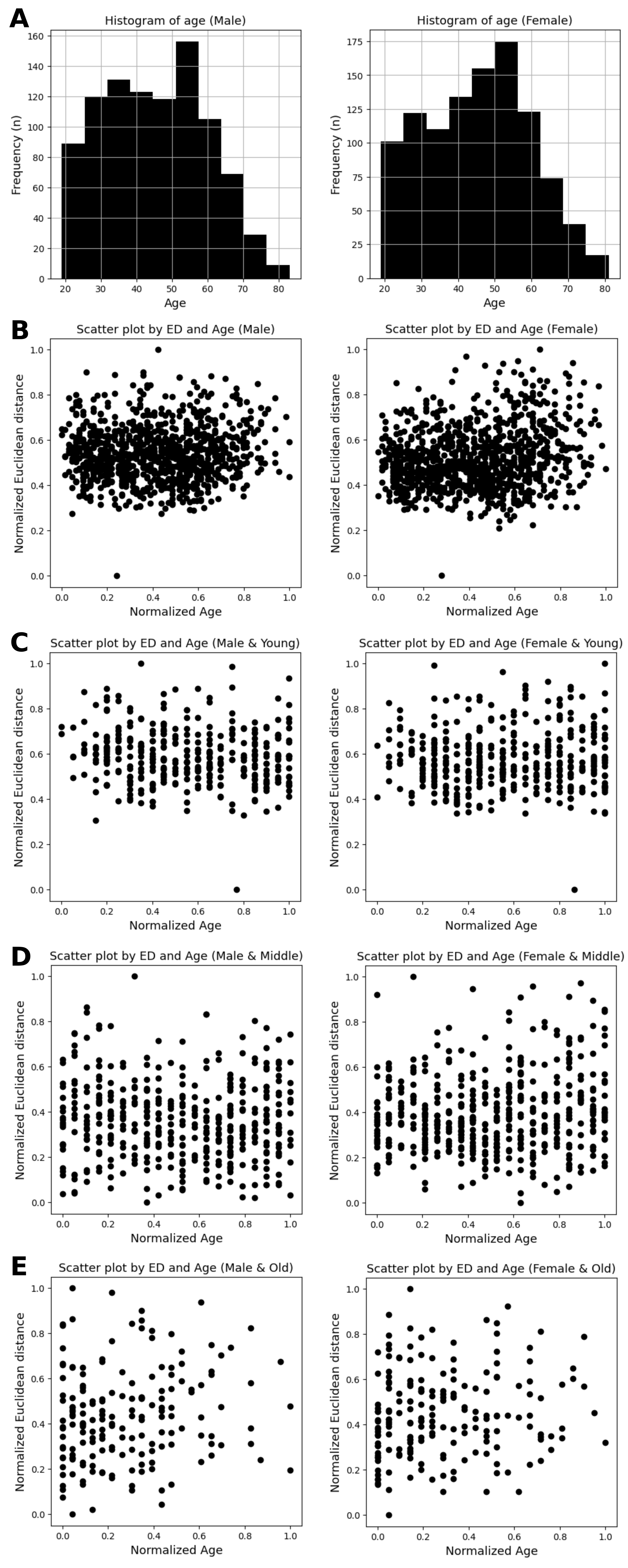

Supplementary Figure 6. Correlation between age and ED. (A) Histogram of age in male (left) and female subjects (right) (B) For 949 male subjects (mean age: 45.4 ± 14.5 years), the PCC was 0.075 (p-value = 0.022) between age and ED (mean: 4.4 ± 1.0). For 1051 female subjects (mean age: 45.4 ± 14.1 years), the PCC was 0.197 (p-value < 0.01) between age and ED (mean: 4.0 ± 1.0). (C) For 360 young male subjects (mean age: 30.1 ± 5.4 years), the PCC was -0.128 (p-value = 0.015) between age and ED (mean: 4.3 ± 0.9). For 386 young female subjects (mean age: 30.2 ± 5.6 years), the PCC was 0.028 (p-value = 0.588) between age and ED (mean: 3.9 ± 0.9). (D) For 415 middle male subjects (mean age: 49.7 ± 5.7 years), the PCC was -0.068 (p-value = 0.168) between age and ED (mean: 4.4 ± 1.0). For 477 middle female subjects (mean age: 49.6 ± 5.6 years), the PCC was 0.109 (p-value = 0.017) between age and ED (mean: 3.9 ± 1.0). (E) For 174 old male subjects (mean age: 66.5 ± 5.5 years), the PCC was 0.146 (p-value = 0.055) between age and ED (mean: 4.6 ± 0.9). For 188 old female subjects (mean age: 66.1 ± 5.3 years), the PCC was 0.117 (p-value = 0.111) between age and ED (mean: 4.4 ± 1.1).

PCC, Pearson’s Correlation Coefficient; ED, Euclidean Distance

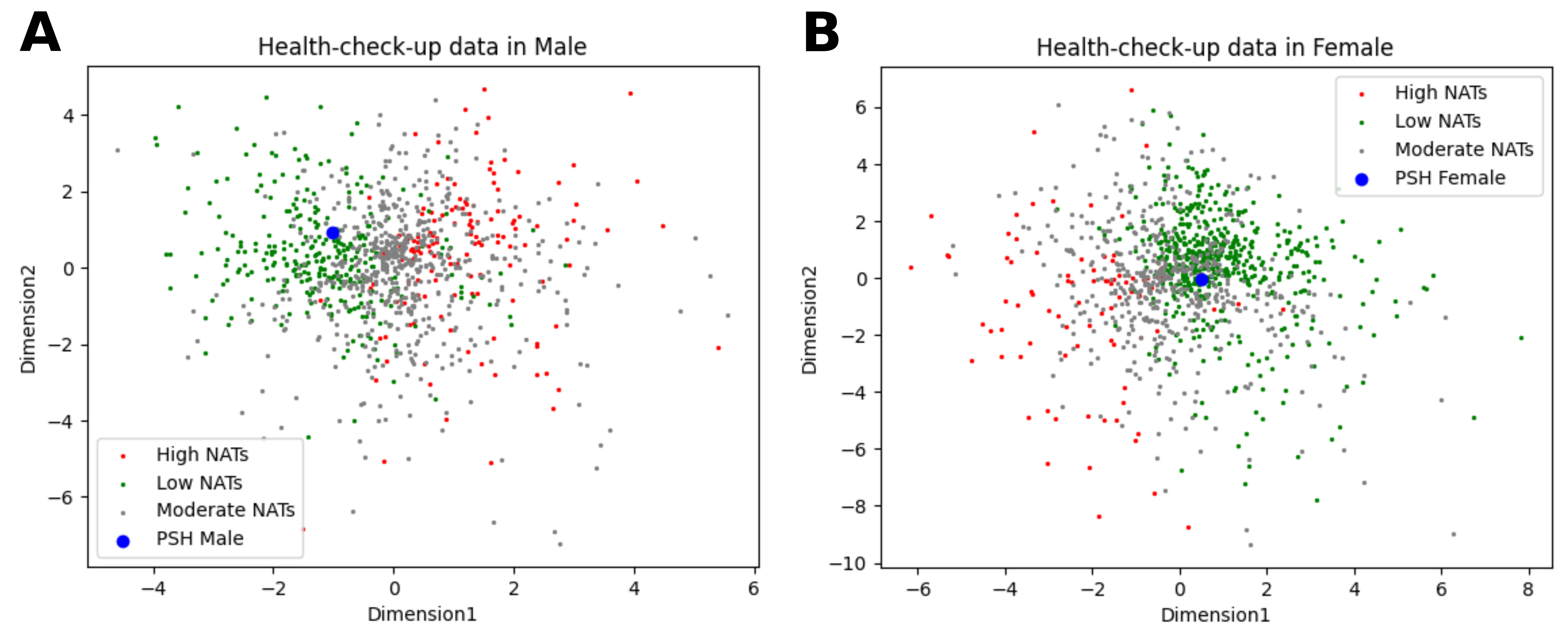

Supplementary Figure 7. Scatter plot of KGP subjects’ health-check-up data colorized by NATs class and PSH after dimensionality reduction using Autoencoder.

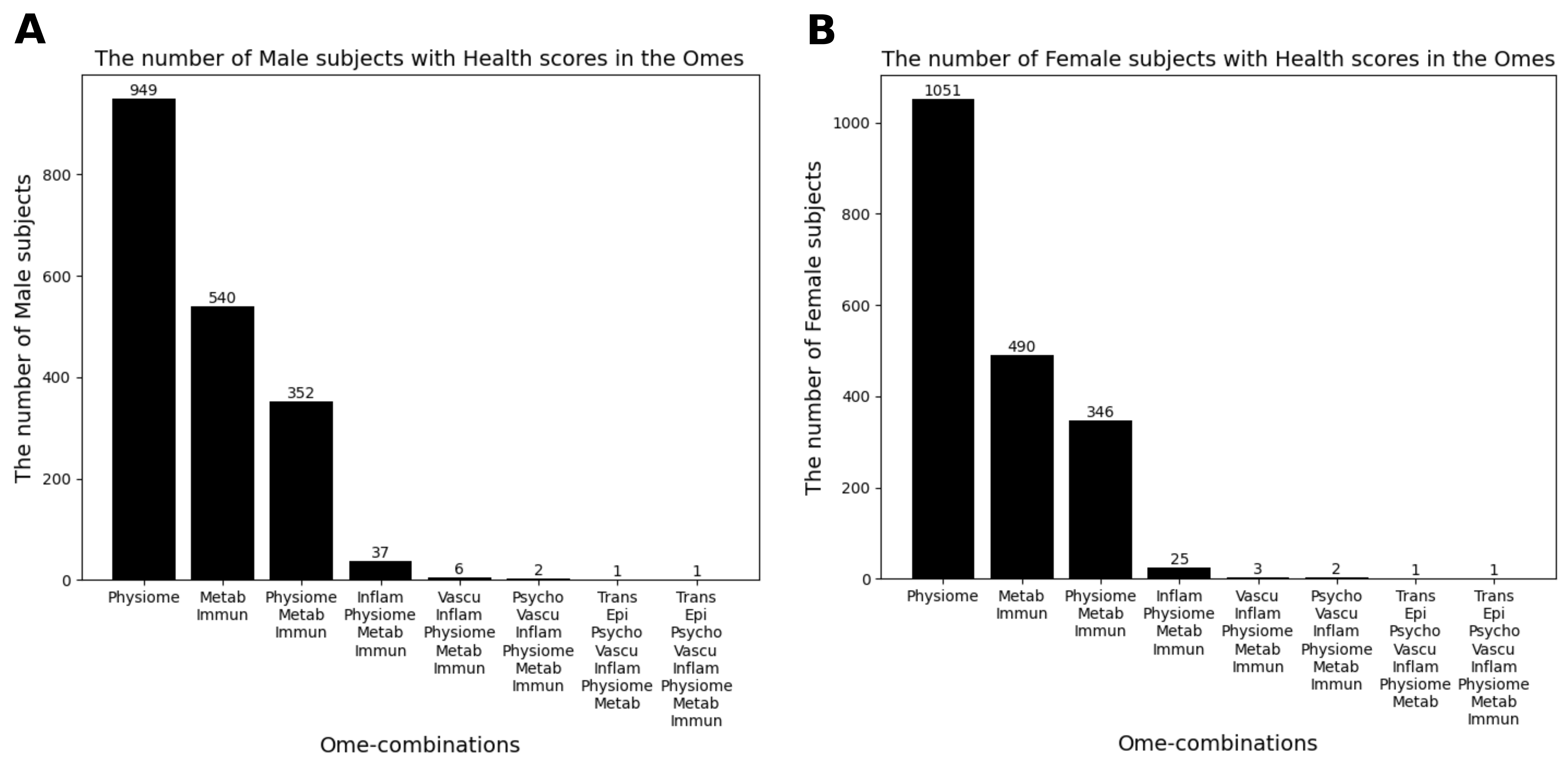

Supplementary Figure 8. Merging health scores (Euclidean distances and Unhealthy probabilities) of subjects drastically reduces the available number of subjects. (A) The number of Male subjects with Health scores in the ome-combination. (B) The number of Female subjects with Health scores in the ome-combination.

Metab, Metabolome; Immun, Immunome; Inflam, Inflammatome; Vascu, Vasculome; Psycho, Psycholome; Trans, Transcriptome; Epi, Epigenome.

### **Supplementary Tables**

Supplementary Table 1. Disease name and distribution by disease diagnosis or self-report

| **Disease name (Variable name)** | **True** | **False** |
| --- | --- | --- |
| Overall disease | 42006 | 63837 |
| Hypertension (di1) | 18682 | 87161 |
| Hyperlipidemia (di2) | 12102 | 93741 |
| Stroke (di3) | 1724 | 104119 |
| Myocardial infarction or angina (di4) | 2162 | 103681 |
| Myocardial infarction (di5) | 774 | 105069 |
| Angina (di6) | 1524 | 104319 |
| Arthritis (dm1) | 10451 | 95392 |
| Osteoarthritis (dm2) | 9161 | 96682 |
| Rheumatoid arthritis (dm3) | 1533 | 104310 |
| Tuberculosis (dj2) | 3201 | 102642 |
| Asthma (dj4) | 3294 | 102549 |
| Thyroid disease (de2) | 2921 | 102922 |
| Diabetes (de1) | 7319 | 98524 |
| Stomach cancer (dc1) | 615 | 105228 |
| Liver cancer (dc2) | 96 | 105747 |
| Colon cancer (dc3) | 401 | 105442 |
| Breast cancer (dc4) | 479 | 105364 |
| Cervical cancer (dc5) | 363 | 105480 |
| Lung cancer (dc6) | 132 | 105711 |
| Other cancers (dc11) | 898 | 104945 |
| Depression (df2) | 3738 | 102105 |
| Atopic dermatitis (dl1) | 4975 | 100868 |
| Kidney disease (dn1) | 451 | 105392 |
| Hepatitis B (dk8) | 1009 | 104834 |
| Hepatitis C (dk9) | 163 | 105680 |
| Liver cirrhosis (dk4) | 244 | 105599 |

Supplementary Table 2. Disease name and distribution by examination result

| **Disease from exam (Variable name)** | **True** | **False** |
| --- | --- | --- |
| Overall disease | 58116 | 47727 |
| Hypertension (he_hp) | 44007 | 33027 |
| Diabetes (he_dm) | 27884 | 43946 |
| Hypercholesterolemia (he_hchol) | 14061 | 57860 |
| hypertriglyceridemia (he_htg) | 8808 | 56864 |
| HBV surface antigen (he_hepab) | 2499 | 81775 |
| Anemia (he_anem) | 7403 | 76587 |

Supplementary Table 3. Pseudo Super Healthy along sex and age groups

| **Traits** | **PSH M**  **PSH F** | **Mean (SD) ^a^** | **Normal**  **range ^b^** | **Units** | **Optimal sources** |
| --- | --- | --- | --- | --- | --- |
| **10**≤**age<20** | | | | | |
| **Physiome** | | | | | |
| Waist | M: 70.46  F: 66.78 | M: 72.56 (10.96)  F: 67.72 (8.6) | M: ≤ 90  F: ≤ 85 | cm | Physiome model |
| BMI | M: 21.28  F: 20.47 | M: 21.31 (3.99)  F: 20.5 (3.56) | 18.5-22.9 | kg/m^2^ | Physiome model |
| SBP | M: 105.1  F: 101.24 | M: 109.98 (10.4)  F: 105.09 (8.84) | < 120 | mmHg | Physiome model |
| DBP | M: 64.54  F: 64.15 | M: 67.0 (9.4)  F: 66.41 (8.05) | < 80 | mmHg | Physiome model |
| Hemoglobin | M: 14.95  F: 13.98 | M: 14.7 (1.07)  F: 13.27 (0.86) | M: 13.5-17.5  F: 12.5-15.5 | g/dL | Physiome model |
| FBS | M: 91.08  F: 88.96 | M: 91.58 (7.83)  F: 89.7 (8.75) | 70-99 | mg/dl | Physiome model |
| HbA1c | M: 5.39  F: 5.35 | M: 5.4 (0.28)  F: 5.36 (0.28) | 4.0-6.0 | % | Physiome model |
| HDL | M: 49.2  F: 52.48 | M: 50.12 (9.31)  F: 53.49 (9.29) | 40-59 | mg/dl | Physiome model |
| Triglycerides | M: 90.02  F: 86.06 | M: 90.37 (51.37)  F: 87.09 (46.57) | < 150 | mg/dl | Physiome model |
| Total cholesterol | M: 157.44  F: 162.97 | M: 159.53 (25.76)  F: 167.75 (24.85) | < 200 | mg/dl | Physiome model |
| Creatinine | M: 0.77  F: 0.62 | M: 0.76 (0.16)  F: 0.62 (0.1) | 0.5-1.4 | mg/dl | Physiome model |
| ALT | M: 19.92  F: 13.53 | M: 20.08 (19.8)  F: 13.47 (10.17) | ≤ 40 | IU/L | Physiome model |
| AST | M: 22.15  F: 18.22 | M: 22.08 (12.07)  F: 18.16 (5.45) | ≤ 40 | IU/L | Physiome model |
| **Metabolome** | | | | | |
| Uric acid | M: 5.95  F: 4.59 | M: 5.98 (1.22)  F: 4.6 (0.89) | M: 3.0-7.0  F: 2.5-5.5 | mg/dL | Metabolome  uric acid model |
| Free T4 | M: 1.37  F: 1.28 | M: 1.36 (0.39)  F: 1.25 (0.21) | 0.8-1.9 | ng/dL | Metabolome  thyroid model |
| TSH | M: 2.88  F: 2.9 | M: 2.83 (1.55)  F: 2.81 (1.77) | 0.4-4.0 | uU/mL | Metabolome  thyroid model |
| **Vasculome** | | | | |  |
| Homocysteine | M: 12.59  F: 8.85 | M: 12.59 (NA)  F: 8.85 (NA) | 5-15 | umol/L | Minimum value in  sex and age groups |
| **20**≤**age<30** | | | | | |
| **Physiome** | | | | | |
| Waist | M: 81.66  F: 71.48 | M: 82.28 (10.53)  F: 72.13 (8.94) | M: ≤ 90  F: ≤ 85 | cm | Physiome model |
| BMI | M: 23.92  F: 21.81 | M: 23.99 (4.01)  F: 21.74 (3.67) | 18.5-22.9 | kg/m^2^ | Physiome model |
| SBP | M: 111.05  F: 100.52 | M: 115.46 (10.65)  F: 105.19 (9.12) | < 120 | mmHg | Physiome model |
| DBP | M: 70.02  F: 65.44 | M: 75.01 (9.24)  F: 69.4 (7.97) | < 80 | mmHg | Physiome model |
| Hemoglobin | M: 15.7  F: 13.86 | M: 15.59 (0.94)  F: 13.12 (0.98) | M: 13.5-17.5  F: 12.5-15.5 | g/dL | Physiome model |
| FBS | M: 90.82  F: 87.98 | M: 90.89 (11.85)  F: 88.57 (11.45) | 70-99 | mg/dl | Physiome model |
| HbA1c | M: 5.36  F: 5.32 | M: 5.35 (0.41)  F: 5.31 (0.41) | 4.0-6.0 | % | Physiome model |
| HDL | M: 49.02  F: 57.18 | M: 49.58 (10.38)  F: 57.57 (11.37) | 40-59 | mg/dl | Physiome model |
| Triglycerides | M: 123.01  F: 83.74 | M: 123.96 (102.7)  F: 84.3 (57.22) | < 150 | mg/dl | Physiome model |
| Total cholesterol | M: 169.46  F: 170.7 | M: 179.21 (30.41)  F: 176.8 (29.57) | < 200 | mg/dl | Physiome model |
| Creatinine | M: 0.98  F: 0.68 | M: 0.94 (0.12)  F: 0.68 (0.09) | 0.5-1.4 | mg/dl | Physiome model |
| ALT | M: 27.38  F: 14.32 | M: 27.79 (24.77)  F: 14.2 (10.19) | ≤ 40 | IU/L | Physiome model |
| AST | M: 23.3  F: 17.77 | M: 23.35 (12.56)  F: 17.76 (6.63) | ≤ 40 | IU/L | Physiome model |
| **Metabolome** | | | | | |
| Uric acid | M: 6.32  F: 4.55 | M: 6.32 (1.17)  F: 4.56 (0.97) | M: 3.0-7.0  F: 2.5-5.5 | mg/dL | Metabolome  uric acid model |
| Free T4 | M: 1.37  F: 1.24 | M: 1.37 (0.17)  F: 1.23 (0.17) | 0.8-1.9 | ng/dL | Metabolome  thyroid model |
| TSH | M: 2.48  F: 2.61 | M: 2.44 (1.44)  F: 2.55 (1.51) | 0.4-4.0 | uU/mL | Metabolome  thyroid model |
| **Vasculome** | | | | |  |
| Homocysteine | M: 5.9  F: 3.58 | M: 10.04 (2.41)  F: 8.04 (1.97) | 5-15 | umol/L | Minimum value in  sex and age groups |
| **30≤age<40** | | | | | |
| **Physiome** | | | | | |
| Waist | M: 84.54  F: 73.1 | M: 85.16 (9.57)  F: 75.74 (9.47) | M: ≤ 90  F: ≤ 85 | cm | Physiome model |
| BMI | M: 24.57  F: 22.46 | M: 24.66 (3.54)  F: 22.53 (3.63) | 18.5-22.9 | kg/m^2^ | Physiome model |
| SBP | M: 112.7  F: 101.79 | M: 116.78 (11.73)  F: 106.28 (10.76) | < 120 | mmHg | Physiome model |
| DBP | M: 73.15  F: 66.22 | M: 78.94 (9.96)  F: 70.79 (8.53) | < 80 | mmHg | Physiome model |
| Hemoglobin | M: 15.42  F: 13.31 | M: 15.49 (0.96)  F: 12.94 (1.13) | M: 13.5-17.5  F: 12.5-15.5 | g/dL | Physiome model |
| FBS | M: 94.75  F: 90.06 | M: 95.35 (15.56)  F: 91.31 (14.2) | 70-99 | mg/dl | Physiome model |
| HbA1c | M: 5.51  F: 5.4 | M: 5.51 (0.53)  F: 5.42 (0.45) | 4.0-6.0 | % | Physiome model |
| HDL | M: 46.76  F: 55.33 | M: 47.26 (10.24)  F: 55.83 (11.84) | 40-59 | mg/dl | Physiome model |
| Triglycerides | M: 159.41  F: 97.03 | M: 161.89 (129.41)  F: 97.39 (65.16) | < 150 | mg/dl | Physiome model |
| Total cholesterol | M: 183.72  F: 175.66 | M: 194.19 (33.15)  F: 183.67 (31.79) | < 200 | mg/dl | Physiome model |
| Creatinine | M: 0.98  F: 0.68 | M: 0.94 (0.12)  F: 0.68 (0.09) | 0.5-1.4 | mg/dl | Physiome model |
| ALT | M: 31.43  F: 16.4 | M: 31.41 (31.75)  F: 16.12 (15.91) | ≤ 40 | IU/L | Physiome model |
| AST | M: 24.89  F: 18.62 | M: 24.9 (15.65)  F: 18.7 (9.15) | ≤ 40 | IU/L | Physiome model |
| **Metabolome** | | | | | |
| Uric acid | M: 6.26  F: 4.42 | M: 6.28 (1.23)  F: 4.41 (0.92) | M: 3.0-7.0  F: 2.5-5.5 | mg/dL | Metabolome  uric acid model |
| Free T4 | M: 1.3  F: 1.24 | M: 1.3 (0.18)  F: 1.22 (0.24) | 0.8-1.9 | ng/dL | Metabolome  thyroid model |
| TSH | M: 2.39  F: 2.82 | M: 2.35 (1.64)  F: 2.75 (2.49) | 0.4-4.0 | uU/mL | Metabolome  thyroid model |
| **Vasculome** | | | | |  |
| Homocysteine | M: 5.23  F: 3.57 | M: 10.3 (2.75)  F: 7.32 (1.84) | 5-15 | umol/L | Minimum value in  sex and age groups |
| **40≤age<50** | | | | | |
| **Physiome** | | | | | |
| Waist | M: 84.32  F: 73.36 | M: 85.03 (8.83)  F: 77.02 (8.67) | M: ≤ 90  F: ≤ 85 | cm | Physiome model |
| BMI | M: 24.25  F: 22.97 | M: 24.37 (3.15)  F: 23.09 (3.36) | 18.5-22.9 | kg/m^2^ | Physiome model |
| SBP | M: 116.03  F: 106.71 | M: 118.37 (13.18)  F: 111.24 (13.51) | < 120 | mmHg | Physiome model |
| DBP | M: 78.84  F: 68.77 | M: 80.6 (10.27)  F: 74.01 (9.22) | < 80 | mmHg | Physiome model |
| Hemoglobin | M: 15.31  F: 13.3 | M: 15.39 (1.02)  F: 12.87 (1.3) | M: 13.5-17.5  F: 12.5-15.5 | g/dL | Physiome model |
| FBS | M: 98.77  F: 93.87 | M: 99.74 (21.04)  F: 94.55 (17.29) | 70-99 | mg/dl | Physiome model |
| HbA1c | M: 5.63  F: 5.49 | M: 5.64 (0.67)  F: 5.52 (0.54) | 4.0-6.0 | % | Physiome model |
| HDL | M: 46.67  F: 54.71 | M: 46.93 (10.25)  F: 55.19 (11.94) | 40-59 | mg/dl | Physiome model |
| Triglycerides | M: 175.23  F: 106.43 | M: 177.5 (147.58)  F: 106.05 (79.36) | < 150 | mg/dl | Physiome model |
| Total cholesterol | M: 189.04  F: 184.51 | M: 199.66 (34.18)  F: 191.88 (32.02) | < 200 | mg/dl | Physiome model |
| Creatinine | M: 0.98  F: 0.7 | M: 0.94 (0.17)  F: 0.7 (0.1) | 0.5-1.4 | mg/dl | Physiome model |
| ALT | M: 29.16  F: 17.02 | M: 28.98 (23.35)  F: 16.75 (12.81) | ≤ 40 | IU/L | Physiome model |
| AST | M: 25.64  F: 19.59 | M: 25.58 (20.03)  F: 19.69 (8.59) | ≤ 40 | IU/L | Physiome model |
| **Metabolome** | | | | | |
| Uric acid | M: 6.05  F: 4.32 | M: 6.07 (1.22)  F: 4.3 (0.9) | M: 3.0-7.0  F: 2.5-5.5 | mg/dL | Metabolome  uric acid model |
| Free T4 | M: 1.28  F: 1.18 | M: 1.27 (0.18)  F: 1.17 (0.21) | 0.8-1.9 | ng/dL | Metabolome  thyroid model |
| TSH | M: 2.5  F: 3.29 | M: 2.47 (1.83)  F: 3.21 (3.99) | 0.4-4.0 | uU/mL | Metabolome  thyroid model |
| **Vasculome** | | | | |  |
| Homocysteine | M: 5.79  F: 4.23 | M: 9.86 (2.33)  F: 7.11 (1.4) | 5-15 | umol/L | Minimum value in  sex and age groups |
| **50≤age<60** | | | | | |
| **Physiome** | | | | | |
| Waist | M: 83.9  F: 74.3 | M: 84.75 (7.84)  F: 78.91 (8.26) | M: ≤ 90  F: ≤ 85 | cm | Physiome model |
| BMI | M: 23.75  F: 23.42 | M: 23.85 (2.74)  F: 23.46 (3.03) | 18.5-22.9 | kg/m^2^ | Physiome model |
| SBP | M: 121.0  F: 114.06 | M: 122.01 (15.2)  F: 117.72 (16.35) | < 120 | mmHg | Physiome model |
| DBP | M: 79.01  F: 70.97 | M: 80.86 (9.82)  F: 76.47 (9.48) | < 80 | mmHg | Physiome model |
| Hemoglobin | M: 15.78  F: 13.75 | M: 15.21 (1.1)  F: 13.28 (0.99) | M: 13.5-17.5  F: 12.5-15.5 | g/dL | Physiome model |
| FBS | M: 101.32  F: 95.49 | M: 101.81 (20.22)  F: 95.95 (14.7) | 70-99 | mg/dl | Physiome model |
| HbA1c | M: 5.74  F: 5.62 | M: 5.73 (0.68)  F: 5.66 (0.49) | 4.0-6.0 | % | Physiome model |
| HDL | M: 47.3  F: 54.08 | M: 47.39 (10.64)  F: 54.51 (12.0) | 40-59 | mg/dl | Physiome model |
| Triglycerides | M: 167.43  F: 119.79 | M: 168.97 (139.69)  F: 119.46 (69.41) | < 150 | mg/dl | Physiome model |
| Total cholesterol | M: 188.95  F: 199.39 | M: 198.33 (33.35)  F: 209.4 (34.47) | < 200 | mg/dl | Physiome model |
| Creatinine | M: 0.96  F: 0.7 | M: 0.93 (0.28)  F: 0.7 (0.11) | 0.5-1.4 | mg/dl | Physiome model |
| ALT | M: 25.95  F: 19.83 | M: 25.5 (16.61)  F: 19.48 (12.86) | ≤ 40 | IU/L | Physiome model |
| AST | M: 25.86  F: 22.44 | M: 25.84 (13.81)  F: 22.4 (12.7) | ≤ 40 | IU/L | Physiome model |
| **Metabolome** | | | | | |
| Uric acid | M: 5.77  F: 4.44 | M: 5.8 (1.15)  F: 4.42 (0.9) | M: 3.0-7.0  F: 2.5-5.5 | mg/dL | Metabolome  uric acid model |
| Free T4 | M: 1.24  F: 1.22 | M: 1.23 (0.18)  F: 1.2 (0.52) | 0.8-1.9 | ng/dL | Metabolome  thyroid model |
| TSH | M: 2.54  F: 3.66 | M: 2.5 (2.48)  F: 3.58 (4.82) | 0.4-4.0 | uU/mL | Metabolome  thyroid model |
| **Vasculome** | | | | |  |
| Homocysteine | M: 5.2  F: 4.57 | M: 10.01 (2.39)  F: 7.69 (1.68) | 5-15 | umol/L | Minimum value in  sex and age groups |
| **60≤age<70** | | | | | |
| **Physiome** | | | | | |
| Waist | M: 83.57  F: 76.08 | M: 84.59 (8.2)  F: 81.19 (8.37) | M: ≤ 90  F: ≤ 85 | cm | Physiome model |
| BMI | M: 23.28  F: 23.71 | M: 23.4 (2.74)  F: 23.75 (3.0) | 18.5-22.9 | kg/m^2^ | Physiome model |
| SBP | M: 123.76  F: 122.27 | M: 124.77 (16.91)  F: 123.73 (17.14) | < 120 | mmHg | Physiome model |
| DBP | M: 73.2  F: 70.79 | M: 77.86 (9.69)  F: 75.79 (8.88) | < 80 | mmHg | Physiome model |
| Hemoglobin | M: 15.57  F: 13.7 | M: 14.94 (1.15)  F: 13.25 (0.91) | M: 13.5-17.5  F: 12.5-15.5 | g/dL | Physiome model |
| FBS | M: 101.8  F: 97.35 | M: 102.17 (19.42)  F: 97.67 (15.56) | 70-99 | mg/dl | Physiome model |
| HbA1c | M: 5.79  F: 5.75 | M: 5.78 (0.68)  F: 5.76 (0.51) | 4.0-6.0 | % | Physiome model |
| HDL | M: 47.53  F: 51.07 | M: 47.81 (11.08)  F: 51.78 (11.5) | 40-59 | mg/dl | Physiome model |
| Triglycerides | M: 148.6  F: 129.68 | M: 148.51 (111.68)  F: 129.27 (79.21) | < 150 | mg/dl | Physiome model |
| Total cholesterol | M: 185.4  F: 200.46 | M: 193.41 (33.53)  F: 209.82 (33.69) | < 200 | mg/dl | Physiome model |
| Creatinine | M: 0.95  F: 0.71 | M: 0.93 (0.14)  F: 0.71 (0.22) | 0.5-1.4 | mg/dl | Physiome model |
| ALT | M: 23.97  F: 19.64 | M: 23.29 (14.38)  F: 19.28 (11.94) | ≤ 40 | IU/L | Physiome model |
| AST | M: 26.19  F: 23.45 | M: 26.14 (14.13)  F: 23.44 (11.82) | ≤ 40 | IU/L | Physiome model |
| **Metabolome** | | | | | |
| Uric acid | M: 5.6  F: 4.39 | M: 5.63 (1.13)  F: 4.36 (0.89) | M: 3.0-7.0  F: 2.5-5.5 | mg/dL | Metabolome  uric acid model |
| Free T4 | M: 1.2  F: 1.18 | M: 1.19 (0.18)  F: 1.17 (0.18) | 0.8-1.9 | ng/dL | Metabolome  thyroid model |
| TSH | M: 3.71  F: 3.18 | M: 3.62 (6.83)  F: 3.13 (2.33) | 0.4-4.0 | uU/mL | Metabolome  thyroid model |
| **Vasculome** | | | | |  |
| Homocysteine | M: 7.09  F: 5.56 | M: 10.9 (2.27)  F: 8.78 (1.71) | 5-15 | umol/L | Minimum value in  sex and age groups |
| **70≤age** | | | | | |
| **Physiome** | | | | | |
| Waist | M: 81.74  F: 77.6 | M: 82.91 (8.88)  F: 80.92 (9.1) | M: ≤ 90  F: ≤ 85 | cm | Physiome model |
| BMI | M: 22.18  F: 23.09 | M: 22.3 (2.77)  F: 23.18 (3.1) | 18.5-22.9 | kg/m^2^ | Physiome model |
| SBP | M: 125.41  F: 129.01 | M: 126.64 (17.14)  F: 129.55 (17.49) | < 120 | mmHg | Physiome model |
| DBP | M: 69.08  F: 70.81 | M: 72.91 (10.17)  F: 73.88 (9.85) | < 80 | mmHg | Physiome model |
| Hemoglobin | M: 15.1  F: 13.48 | M: 14.36 (1.27)  F: 13.01 (1.07) | M: 13.5-17.5  F: 12.5-15.5 | g/dL | Physiome model |
| FBS | M: 99.39  F: 98.93 | M: 99.93 (15.68)  F: 99.01 (15.96) | 70-99 | mg/dl | Physiome model |
| HbA1c | M: 5.77  F: 5.84 | M: 5.79 (0.54)  F: 5.82 (0.53) | 4.0-6.0 | % | Physiome model |
| HDL | M: 47.38  F: 49.83 | M: 47.62 (10.59)  F: 50.14 (10.55) | 40-59 | mg/dl | Physiome model |
| Triglycerides | M: 124.68  F: 132.24 | M: 123.58 (64.88)  F: 131.28 (66.03) | < 150 | mg/dl | Physiome model |
| Total cholesterol | M: 177.73  F: 197.98 | M: 186.83 (32.86)  F: 204.4 (32.59) | < 200 | mg/dl | Physiome model |
| Creatinine | M: 0.97  F: 0.73 | M: 0.96 (0.19)  F: 0.73 (0.17) | 0.5-1.4 | mg/dl | Physiome model |
| ALT | M: 19.69  F: 16.88 | M: 19.31 (9.14)  F: 16.69 (7.06) | ≤ 40 | IU/L | Physiome model |
| AST | M: 24.93  F: 22.96 | M: 25.05 (11.6)  F: 23.04 (6.53) | ≤ 40 | IU/L | Physiome model |
| **Metabolome** | | | | | |
| Uric acid | M: 5.41  F: 4.38 | M: 5.46 (1.19)  F: 4.35 (0.98) | M: 3.0-7.0  F: 2.5-5.5 | mg/dL | Metabolome  uric acid model |
| Free T4 | M: 1.25  F: 1.21 | M: 1.23 (0.18)  F: 1.2 (0.29) | 0.8-1.9 | ng/dL | Metabolome  thyroid model |
| TSH | M: 2.18  F: 3.26 | M: 2.15 (1.47)  F: 3.21 (2.45) | 0.4-4.0 | uU/mL | Metabolome  thyroid model |
| **Vasculome** | | | | |  |
| Homocysteine | M: 8.35  F: 6.25 | M: 12.51 (2.5)  F: 9.38 (2.04) | 5-15 | umol/L | Minimum value in  sex and age groups |

LDL, GGT, Immunome, Inflammatome are not displayed

Hotocysteine (n): M: 1 (1, 10), 2 (60, 10), 3 (86, 10), 4 (91, 10), 5 (106, 10), 6 (63, 10), 7 (46, 10)

F: 1 (1, 10), 2 (77, 10), 3 (103, 10), 4 (74, 10), 5 (89, 10), 6 (63, 10), 7 (32, 10)

Supplementary Table 4. Statistics of Euclidean distance distribution of five omes.

|  | Mean | Std | Min | Max |
| --- | --- | --- | --- | --- |
| Physiome | M: 4.39  F: 4.0 | M: 0.97  F: 1.01 | M: 1.62  F: 1.62 | M: 8.09  F: 7.76 |
| Inflammatome | M: 2.77  F: 3.2 | M: 0.97  F: 0.95 | M: 0.17  F: 0.98 | M: 6.4  F: 6.35 |
| Vasculome | M: 3.38  F: 3.37 | M: 1.0  F: 0.99 | M: 1.12  F: 1.01 | M: 7.12  F: 6.7 |
| Immunome | M: 2.33  F: 2.31 | M: 0.81  F: 0.85 | M: 0.39  F: 0.52 | M: 4.89  F: 5.0 |
| Metabolome | M: 2.37  F: 2.37 | M: 0.71  F: 0.71 | M: 0.69  F: 0.69 | M: 4.8  F: 4.8 |

Supplementary Table 5. 171 diseases or health-checkup traits

**Cancer-Related Variables**

Stomach cancer, Colon cancer, Rectal cancer, Lung cancer, Liver cancer, Prostate cancer, Thyroid cancer, Parathyroid carcinoma, Bladder cancer, Kidney cancer, Pancreatic cancer, Non-Hodgkin lymphoma, Malignant lymphoma, Breast cancer, Breast fibroadenoma, Melanoma, Cervical cancer, Endometrial cancer, Gallbladder cancer, Leukemia, Ovarian cancer, Brain cancer, Esophageal cancer, Testicular cancer, Duodenal cancer, Larynx cancer, Tonsillar cancer, Malignant bone tumor, Glioma, No cancer

**Benign Tumors and Non-Malignant Conditions**

Colon polyps, Uterine fibroids, Neurofibromatosis, Lipoma, Pituitary adenoma, Thyroid nodule

**Metabolic Diseases**

Diabetes mellitus type 1, Diabetes mellitus type 2, Graves' disease, Hashimotos thyroiditis, Hypothyroidism, Primary hyperparathyroidism, Growth hormone deficiency, PolyCystic Ovary Syndrome, Lactose intolerance, Hypercholesterolemia, Hypertriglyceridemia, Gout, Hemochromatosis, Cystic fibrosis, Gilbert syndrome, Obesity, No metabolic disease

**Blood Diseases**

Iron deficiency anemia, Pernicious anemia, Folate deficiency anemia, AIHA, ITP, Other thrombophilia, No blood disease

**Infectious Diseases**

AIDS

**Vitamin Deficiencies**

Avitaminosis

**Allergic and Immunological Conditions**

Hives, Allergic rhinitis

**Neurological Diseases**

Alzheimer's disease, Palsy, Parkinson's disease, Cerebral palsy, Epilepsy, Narcolepsy, Schizophrenia, Bipolar disorder, Depressive disorder, Intellectual disability, Attention deficit disorder, Cluster headaches, Chronic tension headaches, Migraine with aura, Migraine without aura, Essential tremor, Restless legs syndrome, Spinal muscular atrophy, Multiple sclerosis, Trigeminal neuralgia, Bell's palsy, Carpal tunnel syndrome, HMSN, Other peripheral neuropathy, Muscular dystrophy, Myasthenia Gravis, Recurrent sleep paralysis, Sleep disorder, No neuro disease

**Visual and Auditory Diseases**

Retinal detachment, Diabetic retinopathy, Hypertensive retinopathy, Central serous retinopathy, AMD, Retinitis pigmentosa, Glaucoma, Cataract, Age-related cataract, Traumatic cataract, Hyperopia, Myopia, Astigmatism, Presbyopia, Color blindness, Keratoconus, Dry eye syndrome, Strabismus, Floaters, Congenital nystagmus, Meniere's disease, Otosclerosis, Age-related hearing loss, Tinnitus, Congenital deafness, Pterygium, No vis auditory disease

**Cardiovascular and Circulatory Diseases**

Hypertension, Myocardial infarction, Angina, Mitral valve prolapse, Hypertrophic cardiomyopathy, Dilated cardiomyopathy, Restrictive cardiomyopathy, Other cardiomyopathy, WPW syndrome, Heart block, Atrial fibrillation, PVCs, Cardiac arrhythmia, Heart failure, Hyperlipidemia, Stroke, Aortic aneurysm, Other aneurysm, Raynaud's phenomenon, Kawasaki disease, DVT, Varicose veins, Hemorrhoids, Varicocele, No circulatory disease

**Respiratory and ENT (Ear, Nose and Throat) Diseases**

Deviated septum, Nasal polyps, Chronic sinusitis, Acute otitis media, Chronic otitis media, Chronic tonsillitis, Chronic bronchitis, Emphysema, Pneumonia, Pneumothorax, Asthma, ARDS, COPD

**Health Check-Up Data**

Age, Waist, Body Mass Index, Systolic BP, Diastolic BP, Hemoglobin, FBS (Fasting Blood Sugar), HDL, Neutral Fat, Total Cholesterol, Creatinine, ALT, AST

Supplementary Table 6. Selected 557 multiomics features for Psycholome

| **features** | **p-value** | **Adjusted p-value** |
| --- | --- | --- |
| chr8:56113608 | 4.55E-17 | 3.1E-11 |
| chr8:56113627 | 3.82E-15 | 1.3E-09 |
| chr8:56113606 | 2.08E-14 | 4.73E-09 |
| chr8:56113625 | 3.53E-14 | 5.03E-09 |
| XRCC5 | 3.68E-14 | 5.03E-09 |
| chrX:2583990 | 5.1E-14 | 5.8E-09 |
| ATP6V1E1 | 8.15E-14 | 7.94E-09 |
| chr16:30610279 | 1.64E-13 | 0.000000014 |
| chr10:119892349 | 1.84E-13 | 0.000000014 |
| TRIP4 | 2.45E-13 | 1.67E-08 |
| chr7:19708978 | 4.58E-13 | 2.61E-08 |
| chr7:19708990 | 4.45E-13 | 2.61E-08 |
| ARPC2 | 6.98E-13 | 3.66E-08 |
| chrX:2583997 | 1.01E-12 | 0.000000046 |
| SYF2 | 1E-12 | 0.000000046 |
| chr8:56113602 | 1.52E-12 | 0.000000065 |
| chr16:30610288 | 1.66E-12 | 6.65E-08 |
| chr7:19708981 | 1.85E-12 | 7.03E-08 |
| chr2:26345821 | 2.1E-12 | 7.55E-08 |
| chr2:12718444 | 4.11E-12 | 0.00000014 |
| chr11:64304843 | 6.32E-12 | 0.000000205 |
| SNW1 | 7.36E-12 | 0.000000228 |
| chr19:36053922 | 1.06E-11 | 0.000000316 |
| CNBP | 1.12E-11 | 0.000000318 |
| chr14:75064071 | 1.73E-11 | 0.000000445 |
| chr2:26345840 | 1.76E-11 | 0.000000445 |
| ATP6V1D | 1.7E-11 | 0.000000445 |
| chr1:235648940 | 2.11E-11 | 0.000000514 |
| chr13:46211578 | 2.68E-11 | 0.000000631 |
| chr19:36053918 | 4.33E-11 | 0.000000985 |
| chr19:36053913 | 1.58E-10 | 0.00000349 |
| SRP14 | 1.76E-10 | 0.00000376 |
| GTF2B | 2.33E-10 | 0.00000468 |
| chr14:75064065 | 2.27E-10 | 0.00000468 |
| chr17:38297137 | 2.6E-10 | 0.00000507 |
| chr3:143973413 | 3.77E-10 | 0.00000664 |
| chr16:30610276 | 3.8E-10 | 0.00000664 |
| chr18:46104880 | 3.62E-10 | 0.00000664 |
| chr20:13784929 | 3.51E-10 | 0.00000664 |
| DRG1 | 3.92E-10 | 0.00000668 |
| chr2:176002858 | 4.76E-10 | 0.00000792 |
| chr15:76335338 | 5.36E-10 | 0.00000871 |
| chr11:64304866 | 5.95E-10 | 0.00000944 |
| chr3:143973419 | 6.32E-10 | 0.00000981 |
| chr8:22605329 | 6.92E-10 | 0.0000105 |
| chr17:38297134 | 8.13E-10 | 0.0000121 |
| chr20:5950586 | 8.83E-10 | 0.0000126 |
| chr18:32093343 | 8.69E-10 | 0.0000126 |
| chr11:28108247 | 9.65E-10 | 0.0000134 |
| chr7:19708988 | 1.05E-09 | 0.0000144 |
| ZNF622 | 1.22E-09 | 0.0000163 |
| chr13:57634666 | 1.36E-09 | 0.0000179 |
| chr11:28108210 | 1.62E-09 | 0.0000204 |
| EIF2S2 | 1.61E-09 | 0.0000204 |
| CUTC | 1.64E-09 | 0.0000204 |
| HNRNPC | 2.07E-09 | 0.0000253 |
| chr1:7075402 | 2.49E-09 | 0.0000289 |
| chr7:66996879 | 2.44E-09 | 0.0000289 |
| chr14:31457055 | 2.5E-09 | 0.0000289 |
| CLP1 | 2.61E-09 | 0.0000297 |
| chr18:32093329 | 2.67E-09 | 0.0000299 |
| chr19:36916250 | 2.77E-09 | 0.0000304 |
| chr6:6004843 | 3.02E-09 | 0.0000327 |
| chr20:5950575 | 3.19E-09 | 0.000034 |
| chr15:43493482 | 3.29E-09 | 0.0000346 |
| chr8:76681424 | 3.73E-09 | 0.0000385 |
| chr20:13784936 | 4.33E-09 | 0.0000441 |
| GNL2 | 4.6E-09 | 0.0000462 |
| chr19:51904628 | 6.03E-09 | 0.0000596 |
| chr16:75623202 | 6.37E-09 | 0.0000621 |
| chr3:138329259 | 7.22E-09 | 0.0000694 |
| SCCPDH | 8.27E-09 | 0.0000783 |
| PSMC1 | 8.56E-09 | 0.00008 |
| FMNL1-DT | 8.85E-09 | 0.0000816 |
| chr19:47514885 | 9.49E-09 | 0.0000864 |
| chr3:122680934 | 9.67E-09 | 0.0000868 |
| chr7:122305171 | 9.88E-09 | 0.0000875 |
| chr12:45991909 | 1.02E-08 | 0.0000893 |
| chr15:76335325 | 1.07E-08 | 0.0000922 |
| PSMC2 | 1.13E-08 | 0.0000945 |
| EOLA2-DT | 1.15E-08 | 0.0000945 |
| chr3:169666240 | 1.11E-08 | 0.0000945 |
| chr7:27152245 | 1.15E-08 | 0.0000945 |
| chr3:72100598 | 1.18E-08 | 0.0000956 |
| UTP6 | 1.23E-08 | 0.0000988 |
| CCT5 | 1.27E-08 | 0.000101 |
| chr14:39170180 | 1.53E-08 | 0.00012 |
| EAPP | 0.000000018 | 0.00014 |
| chr2:172086863 | 1.88E-08 | 0.000144 |
| chr15:62067266 | 1.93E-08 | 0.000146 |
| chr19:51904625 | 2.12E-08 | 0.000157 |
| PCMT1 | 2.12E-08 | 0.000157 |
| NDUFA1 | 2.24E-08 | 0.000161 |
| chr19:47514890 | 2.23E-08 | 0.000161 |
| ZNF350 | 2.25E-08 | 0.000161 |
| chr19:7874672 | 2.35E-08 | 0.000167 |
| MAGOH | 2.61E-08 | 0.000182 |
| ZCCHC17 | 2.64E-08 | 0.000182 |
| TIMM23 | 2.62E-08 | 0.000182 |
| ZMAT2 | 3.11E-08 | 0.000211 |
| ISY1 | 3.12E-08 | 0.000211 |
| chr6:151241401 | 3.24E-08 | 0.000216 |
| chr2:36967143 | 3.37E-08 | 0.000217 |
| chr15:51623027 | 3.33E-08 | 0.000217 |
| chr1:50968337 | 3.32E-08 | 0.000217 |
| chr15:55196952 | 3.36E-08 | 0.000217 |
| chr7:66996887 | 3.44E-08 | 0.000219 |
| chr13:57634660 | 3.57E-08 | 0.000225 |
| TMED9 | 3.93E-08 | 0.000246 |
| chr3:196942966 | 4.01E-08 | 0.000249 |
| chr19:4342487 | 0.000000041 | 0.000252 |
| chr8:22605340 | 4.42E-08 | 0.000269 |
| chr5:133611631 | 4.47E-08 | 0.00027 |
| chr12:50085118 | 4.65E-08 | 0.000278 |
| chr3:169666246 | 4.71E-08 | 0.000279 |
| DNAJA1 | 4.86E-08 | 0.000286 |
| chr2:172086869 | 0.00000005 | 0.000292 |
| chr13:98576066 | 5.96E-08 | 0.000344 |
| chr19:43935131 | 6.37E-08 | 0.000365 |
| TAF12 | 6.41E-08 | 0.000365 |
| chr14:39170177 | 7.21E-08 | 0.000406 |
| SNX2 | 0.000000075 | 0.00042 |
| RBMX2 | 8.17E-08 | 0.000453 |
| AIF1 | 8.69E-08 | 0.000475 |
| chr2:75200721 | 8.64E-08 | 0.000475 |
| chr6:151241407 | 8.89E-08 | 0.000477 |
| COPS4 | 8.85E-08 | 0.000477 |
| chr17:75633176 | 9.24E-08 | 0.000486 |
| chr6:1605726 | 9.26E-08 | 0.000486 |
| chr8:22367823 | 9.24E-08 | 0.000486 |
| chr4:102826677 | 9.67E-08 | 0.000504 |
| PRSS53 | 0.000000103 | 0.000533 |
| RPA2 | 0.000000106 | 0.000544 |
| chr7:27152238 | 0.000000113 | 0.000573 |
| chr22:37953218 | 0.000000119 | 0.000599 |
| chr2:36967137 | 0.000000126 | 0.000631 |
| chr5:472719 | 0.000000127 | 0.000631 |
| chr3:185937848 | 0.00000013 | 0.000641 |
| chr2:28869971 | 0.000000131 | 0.000644 |
| FTX | 0.000000134 | 0.000655 |
| chr5:88268688 | 0.000000146 | 0.000709 |
| chr7:66996885 | 0.000000148 | 0.000712 |
| chr17:75666956 | 0.000000151 | 0.000719 |
| chr11:119169375 | 0.000000159 | 0.000755 |
| LXN | 0.000000164 | 0.000772 |
| HAT1 | 0.000000169 | 0.000788 |
| PCCB | 0.000000171 | 0.000793 |
| chr15:50687166 | 0.000000176 | 0.000813 |
| chr3:134373499 | 0.000000187 | 0.00085 |
| chr18:32093339 | 0.000000187 | 0.00085 |
| chr6:168370364 | 0.000000188 | 0.00085 |
| ZNF264 | 0.000000198 | 0.000889 |
| chr3:49340281 | 0.000000202 | 0.0009 |
| chr3:196942953 | 0.000000205 | 0.00091 |
| GALK2 | 0.000000211 | 0.000929 |
| chr4:73868977 | 0.000000215 | 0.00094 |
| chr7:66996876 | 0.000000217 | 0.000943 |
| chr17:75633169 | 0.000000219 | 0.000944 |
| ERH | 0.000000226 | 0.000969 |
| ATP5PF | 0.000000228 | 0.000973 |
| SPAG7 | 0.000000269 | 0.00114 |
| CIR1 | 0.000000273 | 0.00115 |
| chr6:30213630 | 0.00000028 | 0.00117 |
| chr19:47256419 | 0.000000308 | 0.00128 |
| chr13:33066604 | 0.000000323 | 0.00133 |
| chr3:40457223 | 0.000000321 | 0.00133 |
| GHITM | 0.000000327 | 0.00134 |
| chr13:98576078 | 0.000000339 | 0.00138 |
| chr10:132767837 | 0.000000349 | 0.00141 |
| chr10:114938337 | 0.000000351 | 0.00141 |
| chr7:85185524 | 0.000000357 | 0.00142 |
| ARPC3 | 0.000000356 | 0.00142 |
| chr11:288662 | 0.000000382 | 0.00151 |
| NME1 | 0.000000388 | 0.00151 |
| IRAG2 | 0.000000388 | 0.00151 |
| chr11:19777890 | 0.000000392 | 0.00152 |
| chr14:76376339 | 0.000000401 | 0.00155 |
| TALAM1 | 0.00000043 | 0.00165 |
| PSMA4 | 0.000000448 | 0.0017 |
| chr11:28108245 | 0.00000045 | 0.0017 |
| chr10:22927766 | 0.00000046 | 0.00173 |
| SLX1B-SULT1A4 | 0.000000493 | 0.00185 |
| RSKR | 0.000000515 | 0.00192 |
| chr5:134648482 | 0.000000524 | 0.00194 |
| chr12:123534446 | 0.000000532 | 0.00196 |
| ISY1-RAB43 | 0.000000537 | 0.00197 |
| METAP2 | 0.00000055 | 0.00201 |
| chr3:31981528 | 0.000000581 | 0.0021 |
| chr22:42872055 | 0.000000582 | 0.0021 |
| STX8 | 0.000000598 | 0.00215 |
| RPF1 | 0.000000606 | 0.00217 |
| SFT2D2 | 0.000000614 | 0.00218 |
| BTBD19 | 0.000000618 | 0.00219 |
| chr19:47514895 | 0.000000636 | 0.00224 |
| chr8:140098995 | 0.000000642 | 0.00225 |
| chr12:113233223 | 0.000000653 | 0.00227 |
| IGBP1 | 0.000000696 | 0.00241 |
| OSER1 | 0.000000716 | 0.00247 |
| PLRG1 | 0.000000722 | 0.00248 |
| chr4:186208345 | 0.000000737 | 0.00251 |
| chr1:231337606 | 0.000000761 | 0.00257 |
| chr14:103905970 | 0.000000758 | 0.00257 |
| PBDC1 | 0.000000769 | 0.00259 |
| ETFA | 0.000000781 | 0.00261 |
| RTCB | 0.000000814 | 0.00271 |
| VPS25 | 0.00000084 | 0.00277 |
| chr15:62067251 | 0.00000084 | 0.00277 |
| chr12:45991877 | 0.000000866 | 0.00284 |
| RABEPK | 0.000000913 | 0.00294 |
| NRBP2 | 0.000000917 | 0.00294 |
| SNRNP40 | 0.000000907 | 0.00294 |
| MMADHC | 0.000000912 | 0.00294 |
| DDX50 | 0.000000918 | 0.00294 |
| CARD16 | 0.000000958 | 0.00305 |
| PABPC1L | 0.000001 | 0.00318 |
| chr1:50968328 | 0.00000101 | 0.00319 |
| ATP5F1C | 0.00000101 | 0.00319 |
| UQCRC2 | 0.00000102 | 0.00319 |
| TMEM199 | 0.00000103 | 0.0032 |
| UVSSA | 0.00000105 | 0.00325 |
| chr1:3903970 | 0.00000108 | 0.00329 |
| chr17:51260096 | 0.00000107 | 0.00329 |
| FAM32A | 0.00000107 | 0.00329 |
| PDCL3 | 0.00000111 | 0.00335 |
| chr7:43729919 | 0.00000111 | 0.00335 |
| chrX:1443644 | 0.00000111 | 0.00335 |
| DNAJA2 | 0.00000111 | 0.00335 |
| chr17:5468965 | 0.00000113 | 0.00337 |
| TAX1BP1 | 0.00000114 | 0.00339 |
| chr11:68904313 | 0.0000012 | 0.00353 |
| NRF1 | 0.00000121 | 0.00353 |
| chr2:674850 | 0.0000012 | 0.00353 |
| chr13:27966304 | 0.00000119 | 0.00353 |
| chr19:38831642 | 0.00000123 | 0.00359 |
| INTS12 | 0.00000125 | 0.00363 |
| VDAC3 | 0.00000126 | 0.00363 |
| ZNF483 | 0.00000127 | 0.00363 |
| ACTR10 | 0.00000126 | 0.00363 |
| chr21:31343797 | 0.00000139 | 0.00396 |
| FABP5 | 0.00000143 | 0.00405 |
| NDUFV2 | 0.00000145 | 0.0041 |
| chr9:131530590 | 0.0000015 | 0.00424 |
| chr2:106160349 | 0.00000152 | 0.00426 |
| TTC1 | 0.00000152 | 0.00426 |
| GMFG | 0.00000158 | 0.0044 |
| HOOK3 | 0.0000016 | 0.00444 |
| chr19:43935121 | 0.00000161 | 0.00445 |
| chr12:130163719 | 0.00000163 | 0.00448 |
| chr14:74241302 | 0.00000165 | 0.00451 |
| CWC22 | 0.00000166 | 0.00454 |
| HSPA4 | 0.00000168 | 0.00457 |
| chr3:134373511 | 0.00000187 | 0.00508 |
| TOR3A | 0.00000191 | 0.00516 |
| chr1:68051743 | 0.00000192 | 0.00516 |
| chr21:31560080 | 0.00000197 | 0.00528 |
| chr4:4248473 | 0.00000211 | 0.00563 |
| chr3:122416428 | 0.00000219 | 0.00581 |
| chr16:1781812 | 0.00000221 | 0.00585 |
| chr8:32548596 | 0.00000229 | 0.006 |
| EIF1AY | 0.00000228 | 0.006 |
| chr18:32093331 | 0.00000237 | 0.00619 |
| chr9:78030553 | 0.00000241 | 0.00627 |
| PAIP2 | 0.00000242 | 0.00627 |
| chr15:51623023 | 0.00000246 | 0.00635 |
| chr12:76084877 | 0.00000248 | 0.00639 |
| chr2:53970485 | 0.00000249 | 0.00639 |
| chr3:129440078 | 0.00000251 | 0.00643 |
| chr12:53295779 | 0.00000256 | 0.00651 |
| chr12:45991873 | 0.00000259 | 0.00658 |
| PARK7 | 0.00000261 | 0.00659 |
| PDHB | 0.00000262 | 0.00659 |
| chr7:96709858 | 0.00000268 | 0.00673 |
| chr9:96875822 | 0.0000027 | 0.00676 |
| chr21:26967654 | 0.00000277 | 0.00691 |
| MYL12A | 0.00000283 | 0.00701 |
| chr8:798506 | 0.00000304 | 0.0075 |
| CIAPIN1 | 0.00000304 | 0.0075 |
| LNPEP | 0.00000307 | 0.00754 |
| chr8:140099021 | 0.00000317 | 0.00774 |
| chr12:130161400 | 0.0000033 | 0.00804 |
| ATP5PB | 0.00000333 | 0.00808 |
| chr19:56093458 | 0.00000344 | 0.00832 |
| EBNA1BP2 | 0.00000351 | 0.00846 |
| CFDP1 | 0.00000352 | 0.00846 |
| POMK | 0.0000036 | 0.0086 |
| HACL1 | 0.0000036 | 0.0086 |
| chr2:172108083 | 0.00000368 | 0.00876 |
| chr2:172108077 | 0.00000372 | 0.00882 |
| chr8:38105400 | 0.00000388 | 0.00916 |
| FRG1 | 0.00000396 | 0.00923 |
| chr17:50508668 | 0.00000396 | 0.00923 |
| ADH5 | 0.00000396 | 0.00923 |
| chr3:49340286 | 0.00000395 | 0.00923 |
| chr7:43729874 | 0.00000401 | 0.00931 |
| chr6:30213620 | 0.00000408 | 0.00943 |
| CCDC12 | 0.00000426 | 0.00974 |
| chr9:89605922 | 0.00000424 | 0.00974 |
| chr13:112134312 | 0.00000424 | 0.00974 |
| EIF2B2 | 0.0000043 | 0.00982 |
| SAP18 | 0.00000437 | 0.00995 |
| chr18:32093322 | 0.00000441 | 0.01 |
| chr19:45092899 | 0.00000456 | 0.0103 |
| chr8:69942829 | 0.00000463 | 0.0104 |
| chr9:91194118 | 0.00000478 | 0.0107 |
| chr5:42951950 | 0.00000487 | 0.0109 |
| chr17:65559579 | 0.00000487 | 0.0109 |
| chr12:53300108 | 0.00000489 | 0.0109 |
| RPL21P120 | 0.00000493 | 0.0109 |
| HNRNPH2 | 0.00000493 | 0.0109 |
| chr8:140098972 | 0.00000494 | 0.0109 |
| chr10:22927759 | 0.00000501 | 0.011 |
| ATG3 | 0.00000504 | 0.011 |
| chr6:39049195 | 0.00000509 | 0.0111 |
| GPN1 | 0.00000514 | 0.0112 |
| chr6:168278130 | 0.00000522 | 0.0113 |
| chr8:25000329 | 0.00000523 | 0.0113 |
| MRPL39 | 0.00000529 | 0.0114 |
| chr15:51623025 | 0.00000549 | 0.0118 |
| UGP2 | 0.00000571 | 0.0121 |
| DDX28 | 0.0000057 | 0.0121 |
| chr16:57091731 | 0.00000569 | 0.0121 |
| chr11:69384674 | 0.00000573 | 0.0122 |
| DNTTIP2 | 0.00000577 | 0.0122 |
| PFDN2 | 0.00000577 | 0.0122 |
| MYL6 | 0.00000581 | 0.0122 |
| chr15:51623035 | 0.00000587 | 0.0123 |
| PSMD7 | 0.00000592 | 0.0123 |
| NDUFAF1 | 0.00000595 | 0.0124 |
| chr9:19102705 | 0.00000598 | 0.0124 |
| chr3:49340288 | 0.00000612 | 0.0126 |
| chr4:151673872 | 0.00000617 | 0.0127 |
| chr5:132815142 | 0.00000621 | 0.0128 |
| chr8:140464362 | 0.00000635 | 0.013 |
| RARS1 | 0.00000641 | 0.0131 |
| SF3B6 | 0.00000666 | 0.0135 |
| chr8:140099045 | 0.00000666 | 0.0135 |
| ANAPC13 | 0.00000676 | 0.0137 |
| chr19:15125278 | 0.00000681 | 0.0137 |
| chr1:15876374 | 0.00000689 | 0.0137 |
| chr20:37521391 | 0.00000691 | 0.0137 |
| chr12:48981757 | 0.0000069 | 0.0137 |
| chr3:72100628 | 0.00000687 | 0.0137 |
| MORF4L1P1 | 0.00000691 | 0.0137 |
| chr15:51623040 | 0.00000696 | 0.0138 |
| ZNF20 | 0.00000698 | 0.0138 |
| chr3:129440084 | 0.00000701 | 0.0138 |
| PPP1R8 | 0.00000712 | 0.014 |
| FEN1 | 0.00000725 | 0.0142 |
| CAPN3 | 0.00000737 | 0.0144 |
| chr15:65611100 | 0.00000743 | 0.0145 |
| chr12:121352361 | 0.0000075 | 0.0146 |
| HDAC2 | 0.00000753 | 0.0146 |
| chr2:142130274 | 0.00000759 | 0.0147 |
| chr1:24642873 | 0.00000775 | 0.0149 |
| chr11:77590231 | 0.00000789 | 0.0152 |
| CWF19L2 | 0.00000807 | 0.0155 |
| chr20:1118371 | 0.00000814 | 0.0155 |
| chr17:82298230 | 0.00000814 | 0.0155 |
| chr2:20294489 | 0.00000838 | 0.0159 |
| chr15:51623032 | 0.00000849 | 0.0161 |
| chr17:50076404 | 0.00000878 | 0.0166 |
| chr19:50051087 | 0.00000886 | 0.0167 |
| RPS4Y1 | 0.00000906 | 0.017 |
| CGAS | 0.00000928 | 0.0174 |
| chr3:196942951 | 0.00000937 | 0.0175 |
| chr7:19708993 | 0.0000094 | 0.0175 |
| DLEU2 | 0.0000095 | 0.0177 |
| EIF3D | 0.00000956 | 0.0177 |
| chr20:329572 | 0.00000953 | 0.0177 |
| chr2:137964756 | 0.00000978 | 0.0178 |
| NDUFB8 | 0.00000973 | 0.0178 |
| DDX49 | 0.00000976 | 0.0178 |
| chr19:50431597 | 0.00000981 | 0.0178 |
| chr14:39170188 | 0.0000098 | 0.0178 |
| chr12:57591361 | 0.0000098 | 0.0178 |
| chr1:12616235 | 0.00000997 | 0.018 |
| TTLL3 | 0.00000996 | 0.018 |
| HMOX2 | 0.00000995 | 0.018 |
| PLCE1 | 0.0000101 | 0.0181 |
| ATF4 | 0.0000101 | 0.0182 |
| H2AZ1 | 0.0000102 | 0.0182 |
| chr4:146636952 | 0.0000104 | 0.0186 |
| SLBP | 0.0000105 | 0.0187 |
| TIMMDC1 | 0.0000106 | 0.0188 |
| chr17:21416455 | 0.0000107 | 0.019 |
| UBL5 | 0.0000109 | 0.0193 |
| chr2:96859041 | 0.0000109 | 0.0193 |
| TXNDC12 | 0.0000111 | 0.0195 |
| PSMB1 | 0.0000111 | 0.0195 |
| chr2:162344227 | 0.0000113 | 0.0196 |
| chr12:57782994 | 0.0000113 | 0.0196 |
| CTNNBL1 | 0.0000113 | 0.0197 |
| DNAJC8 | 0.0000115 | 0.02 |
| chr12:56129212 | 0.0000118 | 0.0204 |
| NSUN5P1 | 0.0000128 | 0.0221 |
| chr2:237486933 | 0.0000129 | 0.0223 |
| SUCLG1 | 0.0000131 | 0.0224 |
| chr21:41802801 | 0.0000131 | 0.0224 |
| chr19:51904643 | 0.0000138 | 0.0236 |
| chr11:77590225 | 0.0000138 | 0.0236 |
| POLR3GL | 0.0000139 | 0.0236 |
| chr5:38427573 | 0.000014 | 0.0237 |
| IK | 0.000014 | 0.0238 |
| MRPL47 | 0.0000141 | 0.0239 |
| CSAD | 0.0000143 | 0.0241 |
| MCOLN3 | 0.0000145 | 0.0244 |
| BTF3 | 0.0000148 | 0.0248 |
| COMMD1 | 0.0000149 | 0.0248 |
| chr8:798487 | 0.0000149 | 0.0248 |
| UNC50 | 0.0000151 | 0.0252 |
| chr13:112587455 | 0.0000152 | 0.0252 |
| chr15:51623043 | 0.0000154 | 0.0255 |
| MRPL51 | 0.0000156 | 0.0257 |
| chr1:32205997 | 0.0000156 | 0.0258 |
| chr1:2917078 | 0.0000158 | 0.026 |
| chr12:45991875 | 0.0000159 | 0.0261 |
| chr2:132669211 | 0.000016 | 0.0261 |
| TTF1 | 0.0000161 | 0.0263 |
| chr17:61462123 | 0.0000162 | 0.0263 |
| DCTN6 | 0.0000162 | 0.0264 |
| DNAJC7 | 0.0000163 | 0.0265 |
| chr17:45262406 | 0.0000165 | 0.0266 |
| chr4:153153181 | 0.0000169 | 0.0272 |
| chr11:22625260 | 0.0000169 | 0.0272 |
| RGMB | 0.0000171 | 0.0275 |
| NUTM2A-AS1 | 0.0000171 | 0.0275 |
| FAM193B | 0.0000172 | 0.0275 |
| chr8:143928873 | 0.0000173 | 0.0276 |
| chr17:29568379 | 0.0000173 | 0.0276 |
| chr2:237486931 | 0.0000175 | 0.0277 |
| chr11:28108240 | 0.0000176 | 0.0278 |
| SLC15A2 | 0.0000179 | 0.0283 |
| chr19:45092893 | 0.000018 | 0.0283 |
| chr12:45991871 | 0.0000182 | 0.0285 |
| THBS4-AS1 | 0.0000181 | 0.0285 |
| CHCHD2 | 0.0000183 | 0.0286 |
| chr21:42879610 | 0.0000184 | 0.0286 |
| chr8:32547797 | 0.0000183 | 0.0286 |
| chr1:161176776 | 0.0000184 | 0.0286 |
| chr12:113233229 | 0.0000186 | 0.0288 |
| HEXD | 0.0000188 | 0.0291 |
| chr1:207321371 | 0.000019 | 0.0294 |
| chr22:41446787 | 0.0000195 | 0.03 |
| chr1:89763307 | 0.0000195 | 0.03 |
| chr17:49223998 | 0.0000198 | 0.0304 |
| HENMT1 | 0.0000201 | 0.0308 |
| chr15:76335332 | 0.0000203 | 0.0311 |
| chr1:112714704 | 0.0000205 | 0.0312 |
| chr15:78872372 | 0.0000207 | 0.0315 |
| AP1G2-AS1 | 0.0000208 | 0.0315 |
| chr2:65396366 | 0.0000208 | 0.0315 |
| chr1:111386766 | 0.0000212 | 0.0319 |
| chr1:22025007 | 0.0000214 | 0.0321 |
| chr17:83108965 | 0.0000214 | 0.0321 |
| BTBD10 | 0.0000216 | 0.0322 |
| CCDC77 | 0.0000216 | 0.0322 |
| HSPE1 | 0.0000215 | 0.0322 |
| TMSB10 | 0.0000217 | 0.0322 |
| chr1:32972861 | 0.0000218 | 0.0322 |
| GIMAP1-GIMAP5 | 0.0000217 | 0.0322 |
| chr8:140099145 | 0.0000218 | 0.0322 |
| chr17:36939917 | 0.0000221 | 0.0327 |
| chr16:2603889 | 0.0000222 | 0.0327 |
| chr22:44729476 | 0.0000225 | 0.0327 |
| SARNP | 0.0000224 | 0.0327 |
| MDM4 | 0.0000225 | 0.0327 |
| chr6:52994844 | 0.0000224 | 0.0327 |
| ANO9 | 0.0000223 | 0.0327 |
| chr17:5468949 | 0.0000227 | 0.0331 |
| chr19:3626886 | 0.0000228 | 0.0331 |
| PAN2 | 0.0000229 | 0.0332 |
| chr17:74204222 | 0.0000231 | 0.0334 |
| EIF2B3 | 0.0000232 | 0.0335 |
| PSMC3 | 0.0000234 | 0.0337 |
| chr5:43020136 | 0.0000236 | 0.0338 |
| chr6:107489639 | 0.0000235 | 0.0338 |
| chr11:45924213 | 0.0000245 | 0.035 |
| ATP5F1A | 0.0000245 | 0.035 |
| chr17:49993520 | 0.0000244 | 0.035 |
| chr2:12718454 | 0.0000248 | 0.0351 |
| DBI | 0.0000248 | 0.0351 |
| chr19:51904640 | 0.0000248 | 0.0351 |
| chr17:4909148 | 0.0000251 | 0.0354 |
| chr22:44729505 | 0.0000251 | 0.0354 |
| PROSER3 | 0.0000256 | 0.036 |
| chr11:123061590 | 0.0000257 | 0.036 |
| R3HCC1L | 0.0000259 | 0.0363 |
| chr1:208243145 | 0.0000264 | 0.0369 |
| ITGA10 | 0.0000267 | 0.0372 |
| KCNAB3 | 0.0000268 | 0.0373 |
| chr3:127676113 | 0.0000268 | 0.0373 |
| RPL6P19 | 0.0000269 | 0.0373 |
| chr5:43020139 | 0.000027 | 0.0374 |
| OIP5-AS1 | 0.0000273 | 0.0377 |
| chr8:140098954 | 0.0000279 | 0.0384 |
| SNRPB2 | 0.000028 | 0.0385 |
| chr9:88076939 | 0.000028 | 0.0385 |
| chr2:101002285 | 0.0000282 | 0.0386 |
| CDC26 | 0.0000284 | 0.0389 |
| chr1:2526560 | 0.0000287 | 0.0391 |
| chr8:140098976 | 0.0000287 | 0.0391 |
| chr21:42652534 | 0.0000288 | 0.0391 |
| PPP1R7 | 0.000029 | 0.0393 |
| RAMAC | 0.0000292 | 0.0394 |
| PSMA5 | 0.0000291 | 0.0394 |
| ROMO1 | 0.0000294 | 0.0396 |
| chr17:65558426 | 0.0000294 | 0.0396 |
| SSBP1 | 0.0000297 | 0.0397 |
| chr4:6523531 | 0.0000296 | 0.0397 |
| IRGQ | 0.0000297 | 0.0397 |
| PGAP1 | 0.0000298 | 0.0398 |
| ACAA2 | 0.00003 | 0.0399 |
| chr5:43020156 | 0.0000303 | 0.0401 |
| ATP5F1B | 0.0000303 | 0.0401 |
| chr17:5485990 | 0.0000304 | 0.0401 |
| chr4:5203652 | 0.0000302 | 0.0401 |
| chr20:45316872 | 0.0000304 | 0.0401 |
| FGFRL1 | 0.0000305 | 0.0402 |
| MDH1 | 0.0000307 | 0.0404 |
| chr10:97150240 | 0.0000308 | 0.0404 |
| chr17:82730032 | 0.0000309 | 0.0405 |
| TLR10 | 0.0000311 | 0.0406 |
| chr2:190437197 | 0.0000311 | 0.0406 |
| chr14:93431064 | 0.0000313 | 0.0408 |
| chr4:51884543 | 0.0000315 | 0.0409 |
| SNRPD1 | 0.0000317 | 0.0411 |
| MFSD14CP | 0.0000317 | 0.0411 |
| POLB | 0.0000319 | 0.0412 |
| ELP6 | 0.000032 | 0.0412 |
| chr5:134648514 | 0.0000323 | 0.0415 |
| chr1:11719366 | 0.0000323 | 0.0415 |
| chr12:50085130 | 0.0000326 | 0.0416 |
| chr2:44938388 | 0.0000326 | 0.0416 |
| PPIB | 0.0000326 | 0.0416 |
| chr17:2049654 | 0.0000327 | 0.0417 |
| DLD | 0.000033 | 0.042 |
| chr7:120054441 | 0.0000335 | 0.0426 |
| chr20:58855879 | 0.0000337 | 0.0427 |
| chr9:129835018 | 0.0000347 | 0.0439 |
| SLU7 | 0.0000355 | 0.0448 |
| chr12:98515446 | 0.0000356 | 0.0449 |
| chr22:42614820 | 0.0000361 | 0.0453 |
| WBP4 | 0.000036 | 0.0453 |
| FBXO48 | 0.0000361 | 0.0453 |
| chr22:17984383 | 0.0000366 | 0.0459 |
| chr17:7562539 | 0.0000373 | 0.0466 |
| COX6A1 | 0.0000374 | 0.0467 |
| BCCIP | 0.0000379 | 0.0471 |
| RUSC1-AS1 | 0.0000381 | 0.0474 |
| chr3:185937854 | 0.0000384 | 0.0477 |
| chr1:2917091 | 0.0000387 | 0.0479 |
| CSTA | 0.0000391 | 0.0483 |
| WDR97 | 0.0000397 | 0.049 |
| ATP5PO | 0.0000398 | 0.0491 |
| chr8:46836788 | 0.0000401 | 0.0493 |
| SEC11A | 0.0000401 | 0.0493 |
| chr5:93603214 | 0.0000408 | 0.05 |

Supplementary Table 7. Selected 1436 transcript features and age feature for Transcriptome model

| **Features** | **p-value** | **Adjusted p-value** |
| --- | --- | --- |
| Age | 9.16E-28 | 1.69E-23 |
| FBLN2 | 5.36E-16 | 4.95E-12 |
| CD248 | 1.15E-15 | 7.05E-12 |
| NRCAM | 1.4E-14 | 6.44E-11 |
| ROBO1 | 6.33E-13 | 2.34E-09 |
| SHANK1 | 7.81E-12 | 0.000000024 |
| CACHD1 | 6.65E-11 | 0.000000175 |
| DNMT3A | 2.01E-10 | 0.000000375 |
| GNAI1 | 1.77E-10 | 0.000000375 |
| NOG | 2.04E-10 | 0.000000375 |
| SLC16A10 | 2.78E-10 | 0.000000428 |
| ARHGEF4 | 2.68E-10 | 0.000000428 |
| ITGA6 | 3.45E-10 | 0.000000455 |
| SATB1 | 3.46E-10 | 0.000000455 |
| OTUD7A | 5.28E-10 | 0.00000065 |
| WNT9A | 6.21E-10 | 0.000000716 |
| MANEAL | 6.66E-10 | 0.000000723 |
| MAML2 | 2.58E-09 | 0.00000264 |
| TAF4B | 3.02E-09 | 0.00000265 |
| LRRN3 | 2.91E-09 | 0.00000265 |
| GIPC3 | 2.75E-09 | 0.00000265 |
| NEFL | 4.77E-09 | 0.000004 |
| NPAS2 | 5.47E-09 | 0.00000439 |
| FLNB | 6.44E-09 | 0.0000048 |
| SMARCA1 | 6.51E-09 | 0.0000048 |
| ABTB3 | 8.26E-09 | 0.00000586 |
| SLC7A3 | 1.06E-08 | 0.00000725 |
| ZBED3 | 1.12E-08 | 0.00000735 |
| ANXA1 | 1.57E-08 | 0.00000996 |
| ZNF518B | 1.75E-08 | 0.0000108 |
| PDE9A | 0.000000022 | 0.0000131 |
| LEF1-AS1 | 2.58E-08 | 0.0000149 |
| ZDBF2 | 3.53E-08 | 0.0000197 |
| CBX2 | 3.88E-08 | 0.0000211 |
| ARHGAP32 | 4.43E-08 | 0.0000234 |
| SERINC5 | 7.11E-08 | 0.0000364 |
| LEF1 | 0.000000079 | 0.0000394 |
| ASIC1 | 8.72E-08 | 0.0000423 |
| AMIGO1 | 9.24E-08 | 0.0000437 |
| IPCEF1 | 0.000000112 | 0.0000516 |
| PLAG1 | 0.000000115 | 0.0000516 |
| SCML1 | 0.000000151 | 0.0000663 |
| SLC4A10 | 0.00000018 | 0.0000771 |
| RETREG1 | 0.000000215 | 0.0000861 |
| HOOK1 | 0.000000213 | 0.0000861 |
| AKAP12 | 0.000000211 | 0.0000861 |
| FOXJ1 | 0.000000252 | 0.0000988 |
| EDAR | 0.000000287 | 0.000107 |
| NRIP1 | 0.00000028 | 0.000107 |
| RHOC | 0.000000291 | 0.000107 |
| TRBV6-4 | 0.000000298 | 0.000108 |
| SPEG | 0.000000373 | 0.000121 |
| REG4 | 0.00000037 | 0.000121 |
| NT5E | 0.000000357 | 0.000121 |
| WNT10A | 0.000000345 | 0.000121 |
| PCDHGB2 | 0.000000359 | 0.000121 |
| PCBP4 | 0.000000364 | 0.000121 |
| OBSCN | 0.000000395 | 0.000126 |
| KLHL6 | 0.000000457 | 0.000143 |
| OXNAD1 | 0.00000047 | 0.000145 |
| RCAN2 | 0.000000496 | 0.000149 |
| CD70 | 0.000000502 | 0.000149 |
| OLMALINC | 0.000000523 | 0.000153 |
| AGMAT | 0.000000579 | 0.000167 |
| IATPR | 0.00000068 | 0.000193 |
| CEP41 | 0.000000781 | 0.000218 |
| SMOC1 | 0.000000882 | 0.000243 |
| CHN1 | 0.000000904 | 0.000245 |
| CTSG | 0.000000921 | 0.000246 |
| GPA33 | 0.000000963 | 0.000248 |
| STXBP6 | 0.000000958 | 0.000248 |
| TTC24 | 0.000000968 | 0.000248 |
| IKZF2 | 0.00000101 | 0.000255 |
| TOGARAM2 | 0.00000109 | 0.000265 |
| SATB2 | 0.00000109 | 0.000265 |
| ABLIM1 | 0.00000107 | 0.000265 |
| PER2 | 0.00000115 | 0.000276 |
| RBMS3 | 0.00000117 | 0.000278 |
| TPPP3 | 0.00000122 | 0.000285 |
| MYC | 0.00000124 | 0.000286 |
| RASGRF2 | 0.00000127 | 0.000288 |
| FGFBP2 | 0.00000135 | 0.000299 |
| PDE7A | 0.00000136 | 0.000299 |
| ENPP2 | 0.00000134 | 0.000299 |
| CHML | 0.00000145 | 0.000312 |
| CASC15 | 0.00000145 | 0.000312 |
| PTK7 | 0.00000159 | 0.000337 |
| TBCB | 0.00000161 | 0.000338 |
| SATB1-AS1 | 0.00000172 | 0.000356 |
| CNST | 0.00000178 | 0.000365 |
| TARBP1 | 0.00000184 | 0.000373 |
| GZMH | 0.0000019 | 0.00038 |
| CCR8 | 0.00000197 | 0.000391 |
| SFRP5 | 0.000002 | 0.000391 |
| PAPSS2 | 0.00000202 | 0.000391 |
| FSBP | 0.00000224 | 0.000431 |
| LINC00824 | 0.00000227 | 0.000432 |
| CAMK4 | 0.00000234 | 0.00044 |
| MEST | 0.00000239 | 0.000446 |
| NDFIP1 | 0.00000246 | 0.000449 |
| KLHL34 | 0.00000246 | 0.000449 |
| RAB34 | 0.00000258 | 0.000462 |
| FAM117B | 0.00000258 | 0.000462 |
| LINC01259 | 0.00000265 | 0.000469 |
| FCGBP | 0.0000028 | 0.000488 |
| DCTN2 | 0.00000279 | 0.000488 |
| SLC23A2 | 0.00000313 | 0.00054 |
| TPPP | 0.00000327 | 0.000559 |
| NELL2 | 0.0000035 | 0.000592 |
| IRS1 | 0.00000357 | 0.000594 |
| GLB1L2 | 0.00000356 | 0.000594 |
| AFAP1L1 | 0.00000391 | 0.000643 |
| RGS9 | 0.00000401 | 0.000649 |
| FOXD2-AS1 | 0.00000401 | 0.000649 |
| ALDH5A1 | 0.00000416 | 0.000656 |
| SLC7A6 | 0.00000413 | 0.000656 |
| ATF7IP2 | 0.00000411 | 0.000656 |
| FLT4 | 0.00000421 | 0.000659 |
| PDE3B | 0.0000043 | 0.000662 |
| DCHS1 | 0.0000043 | 0.000662 |
| SYT11 | 0.00000443 | 0.000675 |
| DDX21 | 0.00000489 | 0.000722 |
| PLPP2 | 0.00000488 | 0.000722 |
| IL6ST | 0.00000478 | 0.000722 |
| APOBEC3H | 0.00000489 | 0.000722 |
| PLCG1-AS1 | 0.00000507 | 0.000742 |
| ATP8A2 | 0.00000514 | 0.000746 |
| LDLRAP1 | 0.00000545 | 0.000774 |
| GCSAML | 0.00000541 | 0.000774 |
| VMO1 | 0.00000537 | 0.000774 |
| KIR3DX1 | 0.00000551 | 0.000776 |
| MTA3 | 0.00000585 | 0.000818 |
| GZMB | 0.00000593 | 0.000823 |
| PUM1 | 0.00000609 | 0.000839 |
| PCDHGA10 | 0.00000622 | 0.000849 |
| GPC2 | 0.0000065 | 0.000882 |
| SPTBN1 | 0.00000661 | 0.000882 |
| CERS6 | 0.0000067 | 0.000882 |
| PATL2 | 0.00000662 | 0.000882 |
| TMEM53 | 0.00000668 | 0.000882 |
| TXNRD3 | 0.00000679 | 0.000888 |
| GPRASP1 | 0.00000719 | 0.000934 |
| PRKAR1B | 0.00000747 | 0.000963 |
| HOXC5 | 0.00000772 | 0.000989 |
| PARP3 | 0.00000793 | 0.000996 |
| AJAP1 | 0.00000788 | 0.000996 |
| WDR13 | 0.00000791 | 0.000996 |
| GATA2 | 0.00000825 | 0.00102 |
| NBEA | 0.00000817 | 0.00102 |
| ZEB1 | 0.00000849 | 0.00103 |
| RAB11FIP5 | 0.00000855 | 0.00103 |
| SCD | 0.00000856 | 0.00103 |
| LEXM | 0.00000838 | 0.00103 |
| ZMYND8 | 0.00000892 | 0.00107 |
| PDK1 | 0.00000895 | 0.00107 |
| TCF7 | 0.0000091 | 0.00108 |
| PTGER2 | 0.00000955 | 0.00111 |
| IFNG | 0.00000959 | 0.00111 |
| FCRL6 | 0.00000951 | 0.00111 |
| STK16 | 0.00000975 | 0.00112 |
| MCOLN3 | 0.00000989 | 0.00113 |
| COMTD1 | 0.00001 | 0.00114 |
| LINC01550 | 0.0000109 | 0.00123 |
| TTC28 | 0.000011 | 0.00123 |
| SPAG6 | 0.000011 | 0.00123 |
| ASL | 0.0000111 | 0.00124 |
| TBXA2R | 0.0000119 | 0.00131 |
| BFSP1 | 0.0000121 | 0.00133 |
| ZNF609 | 0.0000123 | 0.00135 |
| GPRASP2 | 0.0000126 | 0.00136 |
| METAP1D | 0.0000127 | 0.00137 |
| BTBD6 | 0.0000128 | 0.00138 |
| LGALS1 | 0.0000134 | 0.00142 |
| TRDV2 | 0.0000134 | 0.00142 |
| SEC14L2 | 0.0000136 | 0.00143 |
| RNF144A | 0.0000136 | 0.00143 |
| FZD4 | 0.0000138 | 0.00144 |
| LDOC1 | 0.0000144 | 0.00147 |
| PHLDA3 | 0.0000145 | 0.00147 |
| BLOC1S1 | 0.0000146 | 0.00147 |
| PHEX | 0.0000147 | 0.00147 |
| ZNF677 | 0.0000143 | 0.00147 |
| SLC1A7 | 0.0000147 | 0.00147 |
| WDR3 | 0.0000146 | 0.00147 |
| ITGB8 | 0.0000147 | 0.00147 |
| ZNF594 | 0.0000151 | 0.00148 |
| PRAG1 | 0.0000151 | 0.00148 |
| SCCPDH | 0.0000149 | 0.00148 |
| PDZD11 | 0.0000162 | 0.00158 |
| CPA3 | 0.0000169 | 0.00163 |
| LRRC8D | 0.0000169 | 0.00163 |
| EDARADD | 0.0000177 | 0.00169 |
| FOXRED2 | 0.0000176 | 0.00169 |
| CRIP1 | 0.000018 | 0.00171 |
| PAR_Y_IL3RA | 0.0000184 | 0.00171 |
| IL3RA | 0.0000184 | 0.00171 |
| GZMA | 0.0000182 | 0.00171 |
| RAD9A | 0.0000183 | 0.00171 |
| PLEKHA7 | 0.0000186 | 0.00172 |
| SCN7A | 0.0000189 | 0.00174 |
| NAP1L2 | 0.0000189 | 0.00174 |
| CCNDBP1 | 0.0000191 | 0.00175 |
| TRABD2A | 0.0000192 | 0.00175 |
| ADGRA3 | 0.0000193 | 0.00175 |
| SYNGR3 | 0.0000199 | 0.00179 |
| SLC50A1 | 0.0000205 | 0.00181 |
| CCM2 | 0.0000204 | 0.00181 |
| PRSS23 | 0.0000203 | 0.00181 |
| HSPG2 | 0.0000211 | 0.00185 |
| NAA16 | 0.000021 | 0.00185 |
| IGFBP3 | 0.0000216 | 0.00188 |
| PPP2R2B | 0.0000215 | 0.00188 |
| GPR153 | 0.0000218 | 0.00189 |
| ANXA4 | 0.0000221 | 0.00191 |
| IGFBP7 | 0.0000227 | 0.00194 |
| S100A10 | 0.0000226 | 0.00194 |
| DYRK1B | 0.0000234 | 0.00199 |
| LINC00426 | 0.0000241 | 0.00201 |
| SERBP1 | 0.0000241 | 0.00201 |
| CNIH2 | 0.0000242 | 0.00201 |
| FAM83F | 0.0000238 | 0.00201 |
| LMO7 | 0.0000238 | 0.00201 |
| MYO1G | 0.0000245 | 0.00202 |
| TRIM46 | 0.0000246 | 0.00203 |
| SH3RF3 | 0.0000261 | 0.00214 |
| ATM | 0.0000264 | 0.00215 |
| PAICS | 0.0000267 | 0.00217 |
| MIR34AHG | 0.0000275 | 0.00222 |
| B3GAT1 | 0.0000285 | 0.00229 |
| LRP5 | 0.0000292 | 0.00234 |
| ENPP3 | 0.0000302 | 0.00241 |
| RCAN3 | 0.0000303 | 0.00241 |
| BACH2 | 0.0000307 | 0.00243 |
| CCR7 | 0.0000316 | 0.00249 |
| EFR3B | 0.0000331 | 0.00255 |
| PFAS | 0.000033 | 0.00255 |
| LTBP4 | 0.0000331 | 0.00255 |
| ACTR1A | 0.0000324 | 0.00255 |
| SSBP4 | 0.0000331 | 0.00255 |
| RPS6KB2 | 0.0000341 | 0.00262 |
| DEFA1B | 0.0000344 | 0.00263 |
| ADGRG1 | 0.0000352 | 0.00269 |
| SESN2 | 0.0000368 | 0.00277 |
| LINC00892 | 0.0000365 | 0.00277 |
| XPNPEP2 | 0.0000368 | 0.00277 |
| PLK3 | 0.0000373 | 0.00279 |
| SLC45A3 | 0.0000376 | 0.0028 |
| BATF | 0.0000383 | 0.00285 |
| PTGDS | 0.0000384 | 0.00285 |
| FTH1P22 | 0.0000393 | 0.0029 |
| ZFYVE28 | 0.0000398 | 0.00293 |
| CRTC3 | 0.0000408 | 0.00298 |
| STARD4-AS1 | 0.0000426 | 0.00311 |
| CNKSR2 | 0.0000429 | 0.00311 |
| KIAA1217 | 0.0000432 | 0.00313 |
| RTL10 | 0.0000436 | 0.00314 |
| MPPED2 | 0.0000442 | 0.00317 |
| OLFM2 | 0.0000457 | 0.00326 |
| UCK1 | 0.0000464 | 0.0033 |
| RAPGEF6 | 0.0000472 | 0.00335 |
| DMPK | 0.0000477 | 0.00337 |
| TIFAB | 0.0000479 | 0.00338 |
| ZMYND10 | 0.0000492 | 0.00345 |
| RREB1 | 0.00005 | 0.00348 |
| ZFAND2B | 0.0000498 | 0.00348 |
| AEBP1 | 0.0000508 | 0.00352 |
| MTR | 0.000051 | 0.00352 |
| CARMIL3 | 0.0000515 | 0.00355 |
| TIGIT | 0.0000519 | 0.00356 |
| CRIP2 | 0.0000522 | 0.00357 |
| ANKRD18A | 0.0000532 | 0.0036 |
| HNRNPA1P21 | 0.0000528 | 0.0036 |
| CERCAM | 0.0000533 | 0.0036 |
| CLCF1 | 0.0000537 | 0.00361 |
| WDR12 | 0.0000538 | 0.00361 |
| LMNA | 0.0000546 | 0.00363 |
| OBSCN-AS1 | 0.0000544 | 0.00363 |
| TMEM220 | 0.0000547 | 0.00363 |
| CA8 | 0.0000555 | 0.00366 |
| LIN37 | 0.0000555 | 0.00366 |
| C1orf198 | 0.0000558 | 0.00366 |
| PRXL2A | 0.0000564 | 0.00368 |
| SERPINF1 | 0.0000564 | 0.00368 |
| PDGFRB | 0.0000572 | 0.00372 |
| ZNF507 | 0.0000576 | 0.00373 |
| VIM | 0.0000592 | 0.00382 |
| ATOSB | 0.0000594 | 0.00382 |
| FAM229A | 0.0000603 | 0.00386 |
| GPR34 | 0.0000614 | 0.00392 |
| ZNF821 | 0.0000621 | 0.00394 |
| TBX21 | 0.0000622 | 0.00394 |
| P3H2 | 0.0000625 | 0.00395 |
| RASGRF1 | 0.0000639 | 0.00398 |
| MTSS2 | 0.0000636 | 0.00398 |
| CLDND2 | 0.0000637 | 0.00398 |
| ZCCHC12 | 0.0000637 | 0.00398 |
| CDKN2A | 0.0000647 | 0.004 |
| NMUR1 | 0.0000647 | 0.004 |
| PHGDH | 0.0000654 | 0.00403 |
| KATNA1 | 0.0000659 | 0.00405 |
| PLCD1 | 0.000067 | 0.00406 |
| TTLL7 | 0.0000668 | 0.00406 |
| RASL11A | 0.0000663 | 0.00406 |
| LGMN | 0.0000668 | 0.00406 |
| CHD7 | 0.0000675 | 0.00407 |
| STK26 | 0.0000675 | 0.00407 |
| LINC02295 | 0.0000681 | 0.00409 |
| HOXC4 | 0.0000684 | 0.0041 |
| CLINT1 | 0.0000698 | 0.00413 |
| DDB2 | 0.0000694 | 0.00413 |
| FAM102A | 0.0000697 | 0.00413 |
| NOLC1 | 0.0000694 | 0.00413 |
| ATP6V0E2-AS1 | 0.0000706 | 0.00416 |
| GABARAPL1 | 0.0000708 | 0.00416 |
| PLXNB1 | 0.0000728 | 0.00426 |
| LINC02481 | 0.0000734 | 0.00427 |
| RBM20 | 0.0000732 | 0.00427 |
| URB1 | 0.0000742 | 0.00431 |
| PARK7P1 | 0.0000752 | 0.00432 |
| GPM6B | 0.0000749 | 0.00432 |
| FNBP1L | 0.0000757 | 0.00432 |
| LRPPRC | 0.0000749 | 0.00432 |
| ANKS6 | 0.0000754 | 0.00432 |
| STYK1 | 0.000077 | 0.00438 |
| RRAS | 0.0000783 | 0.00444 |
| CD99 | 0.0000785 | 0.00444 |
| CD2AP | 0.00008 | 0.00445 |
| RYR3 | 0.0000799 | 0.00445 |
| RAB6C-AS1 | 0.0000793 | 0.00445 |
| UFD1 | 0.0000795 | 0.00445 |
| LLGL2 | 0.0000798 | 0.00445 |
| BZW2 | 0.00008 | 0.00445 |
| LYPD2 | 0.0000803 | 0.00445 |
| ERBB2 | 0.0000808 | 0.00446 |
| VSIG4 | 0.0000811 | 0.00447 |
| SREBF1 | 0.0000822 | 0.00451 |
| HOPX | 0.0000838 | 0.00459 |
| ZFP69B | 0.0000843 | 0.0046 |
| AMN | 0.0000877 | 0.00476 |
| GLIS3 | 0.0000877 | 0.00476 |
| USP13 | 0.0000886 | 0.00479 |
| PCGF2 | 0.0000904 | 0.00488 |
| GFI1 | 0.000092 | 0.00495 |
| MXRA8 | 0.0000939 | 0.00496 |
| MRTFB | 0.0000941 | 0.00496 |
| MAN1C1 | 0.000094 | 0.00496 |
| LEPROTL1 | 0.0000934 | 0.00496 |
| CYP4F22 | 0.0000938 | 0.00496 |
| SPSB3 | 0.0000926 | 0.00496 |
| PRR5L | 0.0000931 | 0.00496 |
| MTUS1 | 0.0000952 | 0.005 |
| EFHD2-AS1 | 0.0000967 | 0.00505 |
| MIAT | 0.0000969 | 0.00505 |
| FOXJ3 | 0.0000972 | 0.00505 |
| IFI27L2 | 0.0000973 | 0.00505 |
| TBC1D17 | 0.0000974 | 0.00505 |
| PTPRS | 0.0000982 | 0.00507 |
| COLGALT2 | 0.0000994 | 0.00512 |
| TRAV1-2 | 0.0001 | 0.00514 |
| ZNF487 | 0.0001 | 0.00514 |
| TENT4B | 0.000101 | 0.00515 |
| EPHA4 | 0.000102 | 0.0052 |
| SLC18A2 | 0.000103 | 0.00522 |
| ZNF688 | 0.000103 | 0.00523 |
| DALRD3 | 0.000106 | 0.00532 |
| ZNF629 | 0.000105 | 0.00532 |
| FOXO1 | 0.000107 | 0.00536 |
| LINC01226 | 0.000108 | 0.00536 |
| S100A4 | 0.000107 | 0.00536 |
| IER5L-AS1 | 0.000108 | 0.00536 |
| CST7 | 0.000109 | 0.00542 |
| SNHG14 | 0.000111 | 0.0055 |
| SHISA2 | 0.000111 | 0.00551 |
| RNF138 | 0.000112 | 0.00551 |
| ITGB1 | 0.000112 | 0.00551 |
| EML5 | 0.000113 | 0.00553 |
| VPS16 | 0.000114 | 0.00554 |
| SERPINE2 | 0.000114 | 0.00554 |
| MTMR1 | 0.000114 | 0.00554 |
| PELI3 | 0.000114 | 0.00554 |
| GARRE1 | 0.000114 | 0.00554 |
| PAR_Y_WASH6P | 0.000117 | 0.00563 |
| CPXM1 | 0.000118 | 0.00563 |
| TAF1 | 0.000117 | 0.00563 |
| KLHL3 | 0.000118 | 0.00563 |
| WASH6P | 0.000117 | 0.00563 |
| LDHD | 0.000117 | 0.00563 |
| EPAS1 | 0.000121 | 0.00574 |
| CIB1 | 0.000122 | 0.00577 |
| ALPK2 | 0.000122 | 0.00579 |
| GDPD5 | 0.000125 | 0.00585 |
| SFTPD | 0.000125 | 0.00585 |
| ZBTB10 | 0.000125 | 0.00585 |
| IGHV1OR15-3 | 0.000124 | 0.00585 |
| URB2 | 0.000127 | 0.00588 |
| DCT | 0.000127 | 0.00588 |
| ZNF827 | 0.000126 | 0.00588 |
| HIVEP2 | 0.000127 | 0.00588 |
| FASLG | 0.00013 | 0.006 |
| HSF5 | 0.00013 | 0.00601 |
| CYP2J2 | 0.000132 | 0.00609 |
| TPRG1 | 0.000133 | 0.00612 |
| DNAJB5 | 0.000135 | 0.00614 |
| LBX2-AS1 | 0.000135 | 0.00614 |
| KRT73-AS1 | 0.000136 | 0.00618 |
| SPON1 | 0.000137 | 0.00622 |
| ZNF770 | 0.000137 | 0.00622 |
| ARPC4 | 0.000139 | 0.00628 |
| MFGE8 | 0.000141 | 0.00634 |
| SINHCAF | 0.000142 | 0.00638 |
| COPZ2 | 0.000143 | 0.00643 |
| DAB2IP | 0.000144 | 0.00643 |
| S1PR5 | 0.000147 | 0.00655 |
| ZNF550 | 0.000149 | 0.00662 |
| APOOL | 0.000151 | 0.00662 |
| FUZ | 0.000151 | 0.00662 |
| KLHL29 | 0.00015 | 0.00662 |
| ARRDC5 | 0.00015 | 0.00662 |
| CUX2 | 0.000149 | 0.00662 |
| PCOLCE | 0.00015 | 0.00662 |
| SEH1L | 0.000154 | 0.00676 |
| MCOLN2 | 0.000156 | 0.00683 |
| ZNF496 | 0.000157 | 0.00686 |
| SIRPG | 0.000159 | 0.00691 |
| GOLM1 | 0.000161 | 0.00696 |
| RNF167 | 0.00016 | 0.00696 |
| DNTT | 0.000161 | 0.00696 |
| FLJ40194 | 0.000162 | 0.00697 |
| CYP7B1 | 0.000162 | 0.00697 |
| AR | 0.000162 | 0.00697 |
| ZNF101 | 0.000164 | 0.00704 |
| TET1 | 0.000165 | 0.00707 |
| ZNF662 | 0.000168 | 0.00715 |
| GOLGA3 | 0.000169 | 0.00715 |
| NEFH | 0.000168 | 0.00715 |
| PXYLP1 | 0.00017 | 0.00718 |
| C1orf21 | 0.00017 | 0.00718 |
| MMP23B | 0.000176 | 0.00738 |
| VPS37B | 0.000176 | 0.00738 |
| FCGR3A | 0.000175 | 0.00738 |
| TAS2R6P | 0.000177 | 0.0074 |
| ZC3HAV1L | 0.00018 | 0.00752 |
| SNAPC2 | 0.000183 | 0.00758 |
| MLXIP | 0.000183 | 0.00758 |
| PRRC1 | 0.000183 | 0.00758 |
| N4BP3 | 0.000183 | 0.00758 |
| LINC00641 | 0.000187 | 0.00772 |
| ICAM4 | 0.000188 | 0.00772 |
| ASCL5 | 0.000188 | 0.00772 |
| TERT | 0.000187 | 0.00772 |
| TRIM3 | 0.00019 | 0.00776 |
| KLHL13 | 0.000196 | 0.00795 |
| PLCE1 | 0.000195 | 0.00795 |
| LGR4 | 0.000196 | 0.00795 |
| UBASH3B | 0.000199 | 0.008 |
| TRNP1 | 0.000198 | 0.008 |
| KLRF1 | 0.000198 | 0.008 |
| FCHSD1 | 0.000199 | 0.008 |
| PPAT | 0.000205 | 0.00824 |
| JAKMIP2 | 0.000207 | 0.00829 |
| TTC38 | 0.000208 | 0.00831 |
| AJM1 | 0.00021 | 0.00839 |
| KCNQ5-DT | 0.000213 | 0.00846 |
| MTX1 | 0.000213 | 0.00846 |
| TADA3 | 0.000217 | 0.00857 |
| LINC02728 | 0.000216 | 0.00857 |
| GAL3ST4 | 0.000217 | 0.00857 |
| BCKDHB | 0.000219 | 0.00864 |
| POMK | 0.000221 | 0.00868 |
| PSMD9 | 0.000222 | 0.00868 |
| SALL2 | 0.000221 | 0.00868 |
| HCG11 | 0.000223 | 0.00869 |
| BRK1 | 0.000224 | 0.00869 |
| WDR45 | 0.000224 | 0.00869 |
| PPP1R12C | 0.000223 | 0.00869 |
| AK5 | 0.000224 | 0.00869 |
| NAA25 | 0.000226 | 0.00873 |
| COL17A1 | 0.000227 | 0.00874 |
| MED18 | 0.000227 | 0.00874 |
| TFR2 | 0.000228 | 0.00876 |
| GPR173 | 0.000232 | 0.00889 |
| TMEM131L | 0.000236 | 0.00902 |
| PPP1CA | 0.000239 | 0.00912 |
| FAM89B | 0.00024 | 0.00916 |
| GP5 | 0.000245 | 0.0093 |
| ELOVL1 | 0.000246 | 0.00934 |
| DCBLD2 | 0.000248 | 0.00938 |
| MAD2L2 | 0.000248 | 0.00939 |
| NPFF | 0.000253 | 0.00953 |
| UBE2F | 0.000258 | 0.0097 |
| ZFTA | 0.000258 | 0.00971 |
| CRMP1 | 0.000259 | 0.00971 |
| TRGC1 | 0.00026 | 0.00973 |
| GNA14 | 0.000261 | 0.00973 |
| PLEKHG4 | 0.000261 | 0.00973 |
| SIRPG-AS1 | 0.000266 | 0.00987 |
| NINL | 0.000266 | 0.00987 |
| CERS5 | 0.000266 | 0.00987 |
| SYNJ2 | 0.000267 | 0.00988 |
| GCC2 | 0.000271 | 0.00999 |
| ZNF320 | 0.000272 | 0.00999 |
| SRGAP3 | 0.000272 | 0.00999 |
| RASGEF1A | 0.000274 | 0.01 |
| DEFA4 | 0.000279 | 0.0102 |
| MLF1 | 0.000284 | 0.0104 |
| LMLN | 0.000285 | 0.0104 |
| TIMM17B | 0.000287 | 0.0104 |
| RBL2 | 0.000295 | 0.0106 |
| ITPKB | 0.000292 | 0.0106 |
| RCOR3 | 0.000292 | 0.0106 |
| BPI | 0.000295 | 0.0106 |
| MOB2 | 0.000293 | 0.0106 |
| TMEM107 | 0.0003 | 0.0107 |
| LCN2 | 0.0003 | 0.0107 |
| KIFC3 | 0.000298 | 0.0107 |
| DCP1A | 0.000296 | 0.0107 |
| MYL12B | 0.000305 | 0.0109 |
| PI16 | 0.000304 | 0.0109 |
| NAT8L | 0.000309 | 0.011 |
| EXD2 | 0.000316 | 0.0111 |
| SIGLEC10-AS1 | 0.000314 | 0.0111 |
| PLEKHF1 | 0.000316 | 0.0111 |
| ELK4 | 0.000312 | 0.0111 |
| TAGLN2 | 0.000316 | 0.0111 |
| ARMCX2 | 0.000318 | 0.0112 |
| WDR11-DT | 0.000321 | 0.0112 |
| SFXN3 | 0.00032 | 0.0112 |
| FITM2 | 0.000319 | 0.0112 |
| FUBP1 | 0.000323 | 0.0113 |
| TP73-AS1 | 0.000332 | 0.0114 |
| FAM135A | 0.000328 | 0.0114 |
| SYNCRIP | 0.000326 | 0.0114 |
| PODXL2 | 0.000328 | 0.0114 |
| FADS3 | 0.00033 | 0.0114 |
| RANBP2 | 0.000331 | 0.0114 |
| IDH3A | 0.000329 | 0.0114 |
| KCNMB4 | 0.000332 | 0.0114 |
| LAMP5 | 0.000328 | 0.0114 |
| FOXO6 | 0.000332 | 0.0114 |
| LGALS3 | 0.000337 | 0.0115 |
| RAB11FIP2 | 0.000339 | 0.0115 |
| SH2D2A | 0.000337 | 0.0115 |
| TASOR2 | 0.000343 | 0.0116 |
| INPP5K | 0.000341 | 0.0116 |
| ZNF594-DT | 0.000345 | 0.0116 |
| KCNQ5 | 0.000344 | 0.0116 |
| PRKXP1 | 0.000348 | 0.0117 |
| LRRC58 | 0.00035 | 0.0117 |
| DCAF16 | 0.000347 | 0.0117 |
| TBC1D25 | 0.000347 | 0.0117 |
| GEMIN4 | 0.000353 | 0.0118 |
| LAMTOR5 | 0.000355 | 0.0118 |
| GATA2-AS1 | 0.000354 | 0.0118 |
| SEC23IP | 0.000353 | 0.0118 |
| HOXB7 | 0.00036 | 0.0119 |
| SCUBE1 | 0.000358 | 0.0119 |
| ARVCF | 0.000359 | 0.0119 |
| LEMD3 | 0.000362 | 0.012 |
| CEACAM8 | 0.000369 | 0.012 |
| ETS1 | 0.000366 | 0.012 |
| NREP | 0.000362 | 0.012 |
| HDDC2 | 0.000364 | 0.012 |
| N4BP2 | 0.000367 | 0.012 |
| PSENEN | 0.000363 | 0.012 |
| RIF1 | 0.000367 | 0.012 |
| LINC01341 | 0.000374 | 0.0122 |
| UTP20 | 0.00038 | 0.0122 |
| ISM1 | 0.000378 | 0.0122 |
| TNIP2 | 0.000373 | 0.0122 |
| LINC02446 | 0.000375 | 0.0122 |
| FBXL8 | 0.000377 | 0.0122 |
| DDX43 | 0.000379 | 0.0122 |
| FOXD1 | 0.000376 | 0.0122 |
| MTDH | 0.000389 | 0.0124 |
| IP6K2 | 0.000386 | 0.0124 |
| SCART1 | 0.000387 | 0.0124 |
| LRRC45 | 0.000386 | 0.0124 |
| ZNF546 | 0.000385 | 0.0124 |
| IAH1 | 0.000395 | 0.0125 |
| GOLGA8M | 0.000397 | 0.0125 |
| TSG101 | 0.000396 | 0.0125 |
| TMCO4 | 0.000391 | 0.0125 |
| PAG1 | 0.000392 | 0.0125 |
| H2AZ1 | 0.000392 | 0.0125 |
| ARFGEF2 | 0.000399 | 0.0126 |
| SH3GLB2 | 0.000405 | 0.0127 |
| OR10AA1P | 0.000404 | 0.0127 |
| CD8B | 0.000405 | 0.0127 |
| TPH1 | 0.000407 | 0.0127 |
| MSC | 0.000406 | 0.0127 |
| ZSCAN25 | 0.000411 | 0.0128 |
| PRF1 | 0.000412 | 0.0128 |
| TANC1 | 0.000417 | 0.0129 |
| CYTH2 | 0.000416 | 0.0129 |
| ARL6IP5 | 0.000413 | 0.0129 |
| ZCCHC14 | 0.000417 | 0.0129 |
| EFHD2 | 0.000421 | 0.013 |
| LINC02185 | 0.000424 | 0.013 |
| FAM222A | 0.00042 | 0.013 |
| DDI2 | 0.000423 | 0.013 |
| COL5A1 | 0.00042 | 0.013 |
| LATS1 | 0.000426 | 0.0131 |
| PLCL1 | 0.000429 | 0.0131 |
| RBM27 | 0.000433 | 0.0132 |
| PIK3IP1 | 0.000434 | 0.0132 |
| RABEP1 | 0.000438 | 0.0132 |
| DTWD2 | 0.000434 | 0.0132 |
| CRNDE | 0.000436 | 0.0132 |
| ITPRIPL1 | 0.000434 | 0.0132 |
| IGLV1-36 | 0.000436 | 0.0132 |
| ITGB1-DT | 0.000441 | 0.0133 |
| ZBTB20-AS1 | 0.000441 | 0.0133 |
| EPHA2 | 0.000443 | 0.0133 |
| CCNJ | 0.000439 | 0.0133 |
| PM20D2 | 0.000446 | 0.0133 |
| SUGCT | 0.000448 | 0.0133 |
| MTHFD1L | 0.000448 | 0.0133 |
| NKG7 | 0.000446 | 0.0133 |
| WDR36 | 0.000446 | 0.0133 |
| STX8 | 0.000445 | 0.0133 |
| DEPDC7 | 0.000452 | 0.0134 |
| GAB3 | 0.000452 | 0.0134 |
| UBB | 0.000453 | 0.0134 |
| SMIM35 | 0.000457 | 0.0135 |
| CPSF6 | 0.00046 | 0.0135 |
| RBM11 | 0.000457 | 0.0135 |
| C22orf15 | 0.000457 | 0.0135 |
| ERBIN | 0.000461 | 0.0135 |
| CDK2AP2 | 0.000469 | 0.0137 |
| CEACAM6 | 0.000471 | 0.0138 |
| GPR82 | 0.000473 | 0.0138 |
| NLK | 0.000474 | 0.0138 |
| DLG3 | 0.00047 | 0.0138 |
| TSPEAR | 0.000479 | 0.0139 |
| ADAMTSL5 | 0.000488 | 0.0142 |
| LYRM7 | 0.000493 | 0.0142 |
| KCNA5 | 0.00049 | 0.0142 |
| SLC28A3 | 0.000489 | 0.0142 |
| THOC3 | 0.00049 | 0.0142 |
| TSPAN2 | 0.000495 | 0.0143 |
| HK2-DT | 0.000497 | 0.0143 |
| KLF3-AS1 | 0.0005 | 0.0143 |
| SNX9 | 0.000501 | 0.0143 |
| ZNF782 | 0.000499 | 0.0143 |
| MIR23AHG | 0.000495 | 0.0143 |
| TCHP | 0.000511 | 0.0144 |
| IBTK | 0.000504 | 0.0144 |
| PHPT1 | 0.000504 | 0.0144 |
| WDR43 | 0.00051 | 0.0144 |
| ADAMTS14 | 0.000511 | 0.0144 |
| MYO1C | 0.00051 | 0.0144 |
| SLC44A3 | 0.000513 | 0.0144 |
| PET100 | 0.000512 | 0.0144 |
| AKR1C3 | 0.000512 | 0.0144 |
| ZNF280D | 0.00051 | 0.0144 |
| EBP | 0.000515 | 0.0145 |
| NPAT | 0.000516 | 0.0145 |
| PHC3 | 0.000518 | 0.0145 |
| F2R | 0.000517 | 0.0145 |
| KIT | 0.000519 | 0.0145 |
| PRADC1 | 0.000524 | 0.0146 |
| CD34 | 0.000527 | 0.0146 |
| TRIM44 | 0.000524 | 0.0146 |
| TMEM183A | 0.000529 | 0.0146 |
| NUCB2 | 0.000526 | 0.0146 |
| MED30 | 0.000528 | 0.0146 |
| FBXL6 | 0.00053 | 0.0146 |
| MYO6 | 0.000523 | 0.0146 |
| KSR2 | 0.000529 | 0.0146 |
| TSPEAR-AS1 | 0.000535 | 0.0147 |
| KBTBD11 | 0.000543 | 0.0149 |
| COPG2IT1 | 0.000545 | 0.0149 |
| SENP6 | 0.000544 | 0.0149 |
| NR4A1 | 0.000549 | 0.015 |
| CALHM2 | 0.000546 | 0.015 |
| CARMIL1 | 0.000556 | 0.0151 |
| GAPDH | 0.000554 | 0.0151 |
| HDAC9 | 0.00056 | 0.0152 |
| IL7R | 0.000565 | 0.0153 |
| ACTA2 | 0.000565 | 0.0153 |
| TSPAN5 | 0.000575 | 0.0155 |
| MS4A2 | 0.000572 | 0.0155 |
| OLA1P2 | 0.000579 | 0.0156 |
| LYNX1 | 0.000578 | 0.0156 |
| CNN3 | 0.000583 | 0.0156 |
| LDLRAD4 | 0.000578 | 0.0156 |
| SPTSSB | 0.000584 | 0.0156 |
| POF1B | 0.000581 | 0.0156 |
| ZNF22-AS1 | 0.000577 | 0.0156 |
| CNPPD1 | 0.000594 | 0.0158 |
| BCL11B | 0.000593 | 0.0158 |
| RABIF | 0.000592 | 0.0158 |
| MPO | 0.000598 | 0.0159 |
| MAPKAPK2 | 0.000602 | 0.016 |
| VRK3 | 0.000607 | 0.0161 |
| LRP6 | 0.000609 | 0.0161 |
| SH2D1B | 0.000608 | 0.0161 |
| HEATR9 | 0.000607 | 0.0161 |
| GPR19 | 0.00061 | 0.0161 |
| LINC00899 | 0.000614 | 0.0162 |
| SEC24A | 0.000624 | 0.0164 |
| PLXDC1 | 0.000629 | 0.0165 |
| EPHA1 | 0.000638 | 0.0166 |
| MAT2A | 0.000634 | 0.0166 |
| GYPB | 0.000635 | 0.0166 |
| LINC00891 | 0.000635 | 0.0166 |
| ATG4D | 0.000638 | 0.0166 |
| CRLF3 | 0.000635 | 0.0166 |
| GPRIN1 | 0.000645 | 0.0168 |
| TRIM33 | 0.000648 | 0.0168 |
| MOB1B | 0.000666 | 0.0173 |
| UBE3A | 0.000667 | 0.0173 |
| OTUB1 | 0.000671 | 0.0174 |
| BABAM1 | 0.000672 | 0.0174 |
| ZMYM2 | 0.00068 | 0.0175 |
| MYL6 | 0.000681 | 0.0175 |
| TCF4 | 0.000686 | 0.0176 |
| PSMG4 | 0.00069 | 0.0177 |
| CHKB | 0.000688 | 0.0177 |
| SHPRH | 0.0007 | 0.0178 |
| ZMAT1 | 0.000695 | 0.0178 |
| LRRC43 | 0.000698 | 0.0178 |
| DCTN3 | 0.000696 | 0.0178 |
| NCR3LG1 | 0.000698 | 0.0178 |
| HDC | 0.000703 | 0.0179 |
| SLC25A14 | 0.00071 | 0.018 |
| PHLDA1 | 0.000715 | 0.0181 |
| ADAMTS6 | 0.000717 | 0.0182 |
| MPST | 0.000718 | 0.0182 |
| PUS7 | 0.000722 | 0.0182 |
| GPC3 | 0.000718 | 0.0182 |
| RRS1 | 0.000732 | 0.0183 |
| ADNP2 | 0.000729 | 0.0183 |
| MORN4 | 0.000729 | 0.0183 |
| SSTR3 | 0.000728 | 0.0183 |
| ZNF587B | 0.000728 | 0.0183 |
| COMMD4 | 0.000742 | 0.0185 |
| RALGDS | 0.000741 | 0.0185 |
| FOXK1 | 0.000745 | 0.0185 |
| DNAH10 | 0.00074 | 0.0185 |
| DUSP4 | 0.000744 | 0.0185 |
| TMEM168 | 0.000737 | 0.0185 |
| UBIAD1 | 0.000752 | 0.0186 |
| IL12RB1 | 0.000749 | 0.0186 |
| MAP3K6 | 0.000748 | 0.0186 |
| SNRK | 0.00076 | 0.0188 |
| NR3C2 | 0.00076 | 0.0188 |
| TMEM41B | 0.000765 | 0.0189 |
| MTSS1 | 0.000777 | 0.0191 |
| TUBB4B | 0.000779 | 0.0191 |
| DOCK10 | 0.00078 | 0.0191 |
| GDI1 | 0.000775 | 0.0191 |
| TAS2R5 | 0.00079 | 0.0192 |
| RFX7 | 0.000786 | 0.0192 |
| GOLGA8O | 0.000793 | 0.0192 |
| ATP13A3 | 0.000793 | 0.0192 |
| IL23R | 0.000788 | 0.0192 |
| TMIGD2 | 0.000791 | 0.0192 |
| SLC14A1 | 0.000789 | 0.0192 |
| MED25 | 0.000785 | 0.0192 |
| RIC1 | 0.000789 | 0.0192 |
| ATP6V1D | 0.000795 | 0.0193 |
| RAB3GAP1 | 0.000809 | 0.0194 |
| APOBEC3G | 0.000807 | 0.0194 |
| NLGN2 | 0.000804 | 0.0194 |
| TINF2 | 0.000806 | 0.0194 |
| HTRA2 | 0.000804 | 0.0194 |
| TMEM59 | 0.000804 | 0.0194 |
| SDF2 | 0.000808 | 0.0194 |
| PRKD3 | 0.000813 | 0.0195 |
| AFF4 | 0.000817 | 0.0195 |
| CCL4 | 0.000817 | 0.0195 |
| ZNF618 | 0.000825 | 0.0197 |
| STX17 | 0.000834 | 0.0199 |
| SCRN1 | 0.000839 | 0.02 |
| IFT20 | 0.000844 | 0.02 |
| FAM117A | 0.000841 | 0.02 |
| RBM26 | 0.00085 | 0.0202 |
| FRMPD3 | 0.000853 | 0.0202 |
| EGLN3 | 0.000858 | 0.0203 |
| LPCAT4 | 0.000857 | 0.0203 |
| CCDC159 | 0.00086 | 0.0203 |
| PRKCA | 0.000867 | 0.0204 |
| USP51 | 0.000871 | 0.0205 |
| ZNF470 | 0.00087 | 0.0205 |
| KANK2 | 0.000878 | 0.0205 |
| DUXAP1 | 0.000883 | 0.0205 |
| DBNDD2 | 0.000872 | 0.0205 |
| ZNF18 | 0.000875 | 0.0205 |
| PTGER3 | 0.000877 | 0.0205 |
| SERPINB6 | 0.000875 | 0.0205 |
| FAM50A | 0.000883 | 0.0205 |
| NARS2 | 0.000883 | 0.0205 |
| PSMC3IP | 0.00088 | 0.0205 |
| UTP23 | 0.000894 | 0.0207 |
| EPPK1 | 0.000892 | 0.0207 |
| CLK3 | 0.000891 | 0.0207 |
| LINC00205 | 0.0009 | 0.0208 |
| RIPK4 | 0.000906 | 0.0209 |
| ZZZ3 | 0.000911 | 0.021 |
| OR6N1 | 0.000926 | 0.0212 |
| WDR54 | 0.000919 | 0.0212 |
| FCRLB | 0.000926 | 0.0212 |
| LRRC8C | 0.00092 | 0.0212 |
| WDR89 | 0.000932 | 0.0213 |
| ANGEL1 | 0.000931 | 0.0213 |
| PDE4A | 0.000946 | 0.0214 |
| TMEM102 | 0.000942 | 0.0214 |
| UBAC1 | 0.000945 | 0.0214 |
| GLTP | 0.000935 | 0.0214 |
| ST7 | 0.000944 | 0.0214 |
| OAZ1 | 0.000941 | 0.0214 |
| CEP85L | 0.000939 | 0.0214 |
| SYNGAP1 | 0.000937 | 0.0214 |
| TSPAN17 | 0.000951 | 0.0215 |
| ZNF275 | 0.000961 | 0.0216 |
| GNAO1 | 0.000954 | 0.0216 |
| PXDN | 0.000959 | 0.0216 |
| TWNK | 0.000958 | 0.0216 |
| MFSD10 | 0.000965 | 0.0217 |
| THAP3 | 0.000978 | 0.0219 |
| LARP4 | 0.000974 | 0.0219 |
| ATP5IF1 | 0.000978 | 0.0219 |
| NUDT12 | 0.000982 | 0.022 |
| LGR6 | 0.000991 | 0.0221 |
| BUD31 | 0.000989 | 0.0221 |
| NAGS | 0.000991 | 0.0221 |
| COG4 | 0.000993 | 0.0221 |
| OPTN | 0.000991 | 0.0221 |
| PCDHGB5 | 0.000993 | 0.0221 |
| GREM2 | 0.001 | 0.0222 |
| PFDN5 | 0.000999 | 0.0222 |
| DNAI3 | 0.001 | 0.0222 |
| SLC7A1 | 0.00101 | 0.0223 |
| MIEN1 | 0.00101 | 0.0223 |
| ZMYM4 | 0.00102 | 0.0224 |
| ERMAP | 0.00103 | 0.0225 |
| STK38 | 0.00102 | 0.0225 |
| MMP14 | 0.00103 | 0.0225 |
| IGLV3-21 | 0.00102 | 0.0225 |
| ADAMTS4 | 0.00102 | 0.0225 |
| LINC02086 | 0.00103 | 0.0226 |
| GSTK1 | 0.00103 | 0.0226 |
| DYRK2 | 0.00104 | 0.0227 |
| TAF8 | 0.00104 | 0.0227 |
| BSG | 0.00104 | 0.0227 |
| CHMP7 | 0.00105 | 0.0228 |
| ZNF354C | 0.00105 | 0.0228 |
| PELATON | 0.00105 | 0.0228 |
| BAX | 0.00105 | 0.0228 |
| ABI3 | 0.00105 | 0.0228 |
| CAMP | 0.00106 | 0.023 |
| CABLES1 | 0.00107 | 0.0231 |
| MFSD4A | 0.00107 | 0.0231 |
| ZDHHC9 | 0.00107 | 0.0231 |
| HRH4 | 0.00108 | 0.0232 |
| ACVR2B | 0.00108 | 0.0232 |
| NQO1 | 0.00108 | 0.0232 |
| CD2 | 0.00108 | 0.0232 |
| MAMLD1 | 0.00109 | 0.0234 |
| DLGAP4 | 0.0011 | 0.0234 |
| RAB4B | 0.0011 | 0.0234 |
| IMPACT | 0.0011 | 0.0234 |
| TXNDC17 | 0.00109 | 0.0234 |
| REEP4 | 0.0011 | 0.0234 |
| HPGD | 0.0011 | 0.0235 |
| PSMD13 | 0.00111 | 0.0236 |
| ZNF90 | 0.00111 | 0.0236 |
| SOX12 | 0.00111 | 0.0236 |
| C1orf43 | 0.00111 | 0.0236 |
| TNFRSF4 | 0.00112 | 0.0237 |
| TCF25 | 0.00112 | 0.0237 |
| SERTAD3 | 0.00113 | 0.0238 |
| COX7B | 0.00113 | 0.0238 |
| LINC01644 | 0.00114 | 0.0239 |
| TRIM58 | 0.00114 | 0.0239 |
| ANXA2 | 0.00114 | 0.0239 |
| NTRK1 | 0.00114 | 0.0239 |
| ATP1B3 | 0.00113 | 0.0239 |
| SLC39A1 | 0.00113 | 0.0239 |
| ERBB3 | 0.00116 | 0.024 |
| RUNX1 | 0.00117 | 0.024 |
| PDZK1IP1 | 0.00116 | 0.024 |
| SLC25A25-AS1 | 0.00116 | 0.024 |
| CDC14A | 0.00116 | 0.024 |
| PLCG1 | 0.00115 | 0.024 |
| ELOB | 0.00116 | 0.024 |
| RAB5IF | 0.00117 | 0.024 |
| UPP1 | 0.00117 | 0.024 |
| TSEN2 | 0.00116 | 0.024 |
| MT1E | 0.00116 | 0.024 |
| RER1 | 0.00117 | 0.024 |
| MCRIP1 | 0.00116 | 0.024 |
| NUP50-DT | 0.00117 | 0.024 |
| DHX33 | 0.00117 | 0.024 |
| AFF2 | 0.00115 | 0.024 |
| MLC1 | 0.00116 | 0.024 |
| PARD6A | 0.00117 | 0.024 |
| FGD6 | 0.00117 | 0.024 |
| CCDC39 | 0.00117 | 0.024 |
| NSUN6 | 0.00117 | 0.0241 |
| LINC02175 | 0.00118 | 0.0241 |
| HLTF | 0.00118 | 0.0241 |
| EML4 | 0.00118 | 0.0241 |
| SH3RF3-AS1 | 0.00119 | 0.0242 |
| TFAP4 | 0.00119 | 0.0242 |
| GTF2A1 | 0.00119 | 0.0242 |
| SRRM1 | 0.00119 | 0.0242 |
| SORCS3 | 0.00119 | 0.0242 |
| LINC00324 | 0.0012 | 0.0243 |
| TSNAXIP1 | 0.00121 | 0.0244 |
| TNRC6C | 0.0012 | 0.0244 |
| BAD | 0.00121 | 0.0244 |
| UBL5 | 0.00121 | 0.0244 |
| RASSF8 | 0.00121 | 0.0245 |
| SIGLEC6 | 0.00122 | 0.0245 |
| SEMG1 | 0.00122 | 0.0246 |
| EFNA5 | 0.00122 | 0.0246 |
| RMC1 | 0.00123 | 0.0246 |
| LIG3 | 0.00124 | 0.0248 |
| ZNF687-AS1 | 0.00124 | 0.0248 |
| ABCE1 | 0.00124 | 0.0248 |
| RIMKLB | 0.00125 | 0.0249 |
| BMPR2 | 0.00125 | 0.0249 |
| ZNF469 | 0.00126 | 0.025 |
| ARHGEF28 | 0.00126 | 0.025 |
| NLN | 0.00126 | 0.025 |
| CD2BP2 | 0.00126 | 0.0251 |
| ELMOD3 | 0.00127 | 0.0251 |
| CYTOR | 0.00126 | 0.0251 |
| ROR2 | 0.00126 | 0.0251 |
| SOX13 | 0.00128 | 0.0253 |
| PPP1R16A | 0.00129 | 0.0253 |
| LINC01506 | 0.00128 | 0.0253 |
| TPP2 | 0.00128 | 0.0253 |
| PIF1 | 0.00128 | 0.0253 |
| ZNHIT1 | 0.00129 | 0.0254 |
| ZSCAN29 | 0.0013 | 0.0256 |
| SLC25A36 | 0.00131 | 0.0256 |
| CPEB1 | 0.0013 | 0.0256 |
| FIBCD1 | 0.00131 | 0.0257 |
| SOCS6 | 0.00132 | 0.0258 |
| SH3BP4 | 0.00133 | 0.0259 |
| FCMR | 0.00133 | 0.0259 |
| SPATA33 | 0.00133 | 0.0259 |
| QPRT | 0.00133 | 0.0259 |
| USP49 | 0.00134 | 0.026 |
| PCDHGB6 | 0.00134 | 0.026 |
| PORCN | 0.00134 | 0.026 |
| BIRC7 | 0.00135 | 0.0261 |
| TEX101 | 0.00135 | 0.0261 |
| PAXBP1-AS1 | 0.00135 | 0.0261 |
| SLC35G2 | 0.00135 | 0.0261 |
| HAGH | 0.00135 | 0.0261 |
| HBD | 0.00135 | 0.0261 |
| COX6A1 | 0.00135 | 0.0261 |
| ENOSF1 | 0.00136 | 0.0262 |
| TUT4 | 0.00136 | 0.0262 |
| ADAT2 | 0.00136 | 0.0262 |
| COPE | 0.00137 | 0.0263 |
| FUBP3 | 0.00137 | 0.0264 |
| RPS27L | 0.00138 | 0.0265 |
| HAVCR2 | 0.00139 | 0.0265 |
| LINC00954 | 0.00139 | 0.0265 |
| PHF5A | 0.00138 | 0.0265 |
| OIP5-AS1 | 0.00139 | 0.0265 |
| ZNF394 | 0.00139 | 0.0265 |
| N6AMT1 | 0.00138 | 0.0265 |
| SMIM12 | 0.00138 | 0.0265 |
| CMKLR1 | 0.00139 | 0.0265 |
| RABEP2 | 0.0014 | 0.0266 |
| IL24 | 0.00141 | 0.0267 |
| RRP15 | 0.00141 | 0.0267 |
| TOGARAM1 | 0.00141 | 0.0267 |
| ABHD4 | 0.00142 | 0.0268 |
| HCFC1R1 | 0.00142 | 0.0268 |
| RBPMS | 0.00142 | 0.0269 |
| KCMF1 | 0.00143 | 0.0269 |
| ANKRD35 | 0.00143 | 0.0269 |
| TMEM208 | 0.00143 | 0.0269 |
| CD3G | 0.00144 | 0.027 |
| LINC00923 | 0.00144 | 0.027 |
| PNISR | 0.00144 | 0.0271 |
| TCAF2P1 | 0.00145 | 0.0271 |
| SLC38A1 | 0.00145 | 0.0271 |
| PTOV1 | 0.00145 | 0.0271 |
| LINC01398 | 0.00145 | 0.0271 |
| PNPO | 0.00146 | 0.0272 |
| CD63 | 0.0015 | 0.0279 |
| BNIP3 | 0.0015 | 0.0279 |
| BCL2L1 | 0.0015 | 0.0279 |
| TSR1 | 0.00151 | 0.028 |
| BMERB1 | 0.00151 | 0.028 |
| EEA1 | 0.00151 | 0.028 |
| ATP8A1 | 0.00151 | 0.028 |
| MDP1 | 0.00153 | 0.0281 |
| YTHDC2 | 0.00153 | 0.0281 |
| ADAM12 | 0.00152 | 0.0281 |
| GSC | 0.00152 | 0.0281 |
| RNF181 | 0.00153 | 0.0281 |
| PDK2 | 0.00152 | 0.0281 |
| LINC00865 | 0.00152 | 0.0281 |
| YTHDF1 | 0.00154 | 0.0283 |
| ILK | 0.00154 | 0.0284 |
| ABL2 | 0.00155 | 0.0284 |
| SPATA2L | 0.00155 | 0.0285 |
| RPL17P50 | 0.00157 | 0.0287 |
| SLFN5 | 0.00157 | 0.0287 |
| CDKN2B-AS1 | 0.00158 | 0.0288 |
| ARHGAP27P2 | 0.00158 | 0.0288 |
| KICS2 | 0.00158 | 0.0288 |
| DLGAP1-AS1 | 0.00157 | 0.0288 |
| OR10G2 | 0.00158 | 0.0288 |
| AKAP11 | 0.00159 | 0.0289 |
| COBL | 0.00161 | 0.0291 |
| ERCC6L2 | 0.00161 | 0.0291 |
| CSRP1 | 0.0016 | 0.0291 |
| ERCC4 | 0.0016 | 0.0291 |
| RPS6KA3 | 0.00161 | 0.0291 |
| ASXL2 | 0.00161 | 0.0291 |
| HNRNPH1 | 0.00162 | 0.0292 |
| MYADM | 0.00162 | 0.0292 |
| PIK3C2A | 0.00162 | 0.0292 |
| POLL | 0.00162 | 0.0292 |
| ATP11C | 0.00162 | 0.0292 |
| PACSIN1 | 0.00163 | 0.0293 |
| PDIA5 | 0.00163 | 0.0293 |
| SYNGR2 | 0.00163 | 0.0293 |
| MED15 | 0.00166 | 0.0295 |
| ANKRD17 | 0.00165 | 0.0295 |
| PLS3 | 0.00165 | 0.0295 |
| ZNF529 | 0.00166 | 0.0295 |
| ARF5 | 0.00165 | 0.0295 |
| PTPN18 | 0.00165 | 0.0295 |
| MMD | 0.00165 | 0.0295 |
| MBTD1 | 0.00166 | 0.0296 |
| IGHV3-30 | 0.00167 | 0.0297 |
| CKB | 0.00168 | 0.0299 |
| DEGS2 | 0.00169 | 0.0299 |
| DCXR | 0.00169 | 0.0299 |
| ZNHIT6 | 0.00168 | 0.0299 |
| ZNF486 | 0.0017 | 0.03 |
| TET3 | 0.0017 | 0.03 |
| NEGR1 | 0.00171 | 0.0302 |
| KCNK10 | 0.00171 | 0.0302 |
| PIKFYVE | 0.00174 | 0.0305 |
| KRT1 | 0.00173 | 0.0305 |
| IL16 | 0.00173 | 0.0305 |
| ACADSB | 0.00173 | 0.0305 |
| TMEM263 | 0.00175 | 0.0306 |
| PSMA7 | 0.00174 | 0.0306 |
| PRSS22 | 0.00175 | 0.0306 |
| POPDC2 | 0.00174 | 0.0306 |
| LTF | 0.00176 | 0.0307 |
| TOMM40L | 0.00176 | 0.0308 |
| TMA7 | 0.00178 | 0.031 |
| ZNF573 | 0.00177 | 0.031 |
| OR2L2 | 0.00179 | 0.0311 |
| PALS1 | 0.00179 | 0.0311 |
| APP | 0.00179 | 0.0311 |
| CAV2 | 0.00178 | 0.0311 |
| YLPM1 | 0.00179 | 0.0311 |
| RANP4 | 0.00181 | 0.0314 |
| SCARA5 | 0.00181 | 0.0314 |
| DNAH6 | 0.00182 | 0.0315 |
| LINC00624 | 0.00182 | 0.0315 |
| LINC00237 | 0.00183 | 0.0316 |
| LMAN2 | 0.00183 | 0.0316 |
| ATP6V0E1 | 0.00184 | 0.0317 |
| SLC13A5 | 0.00185 | 0.0318 |
| TRIR | 0.00186 | 0.0319 |
| EXPH5 | 0.00185 | 0.0319 |
| NPR3 | 0.00185 | 0.0319 |
| LMBR1 | 0.00185 | 0.0319 |
| H3C10 | 0.00186 | 0.0319 |
| NIN | 0.00186 | 0.032 |
| PMF1 | 0.00187 | 0.0321 |
| TBC1D10B | 0.00188 | 0.0321 |
| OTUD4 | 0.00188 | 0.0321 |
| ZBED4 | 0.00188 | 0.0321 |
| MARS2 | 0.00188 | 0.0321 |
| FOXP1 | 0.00187 | 0.0321 |
| FRS2 | 0.00189 | 0.0322 |
| PSMC6 | 0.00189 | 0.0322 |
| COL6A3 | 0.00189 | 0.0322 |
| AIRE | 0.00191 | 0.0324 |
| PER3 | 0.00191 | 0.0324 |
| GATA3-AS1 | 0.00192 | 0.0325 |
| PHF23 | 0.00192 | 0.0325 |
| ZC3H12D | 0.00192 | 0.0325 |
| UBQLN2 | 0.00194 | 0.0328 |
| SYTL3 | 0.00194 | 0.0328 |
| DMXL1 | 0.00194 | 0.0328 |
| MXD4 | 0.00197 | 0.0331 |
| PHKG2 | 0.00197 | 0.0331 |
| CDC34 | 0.00197 | 0.0331 |
| SARNP | 0.00197 | 0.0331 |
| PPP2R5B | 0.00197 | 0.0331 |
| ZC3H12C | 0.00196 | 0.0331 |
| AP4M1 | 0.00198 | 0.0331 |
| GNLY | 0.00198 | 0.0331 |
| TNS1 | 0.00197 | 0.0331 |
| CXCL2 | 0.00199 | 0.0332 |
| G3BP2 | 0.00199 | 0.0332 |
| BATF3 | 0.00199 | 0.0332 |
| ZNF124 | 0.00201 | 0.0336 |
| SH3RF2 | 0.00202 | 0.0337 |
| NAALADL1 | 0.00203 | 0.0338 |
| GFOD2 | 0.00203 | 0.0338 |
| TMEM35B | 0.00203 | 0.0338 |
| LDHA | 0.00204 | 0.0339 |
| STX17-DT | 0.00205 | 0.034 |
| CHD6 | 0.00205 | 0.034 |
| RASSF4 | 0.00206 | 0.0341 |
| LTK | 0.00207 | 0.0342 |
| KIAA1549 | 0.00207 | 0.0342 |
| USP44 | 0.00208 | 0.0342 |
| UBR3 | 0.00207 | 0.0342 |
| PRKAG1 | 0.00208 | 0.0342 |
| C12orf75 | 0.00209 | 0.0344 |
| COG5 | 0.0021 | 0.0345 |
| GABPA | 0.0021 | 0.0345 |
| ADGRG6 | 0.0021 | 0.0345 |
| C1orf109 | 0.0021 | 0.0345 |
| MBTPS2 | 0.0021 | 0.0345 |
| PAGE2B | 0.00211 | 0.0346 |
| L3MBTL4 | 0.00211 | 0.0346 |
| TNFRSF21 | 0.00211 | 0.0346 |
| FBXO9 | 0.00214 | 0.0347 |
| GRASLND | 0.00214 | 0.0347 |
| SLC26A6 | 0.00213 | 0.0347 |
| BTAF1 | 0.00212 | 0.0347 |
| ENSA | 0.00214 | 0.0347 |
| PTPN11 | 0.00212 | 0.0347 |
| TMEM132C | 0.00214 | 0.0347 |
| BCL2A1 | 0.00214 | 0.0347 |
| PRELID1 | 0.00213 | 0.0347 |
| TANGO2 | 0.00215 | 0.0347 |
| MAP3K7CL | 0.00215 | 0.0347 |
| KCNK17 | 0.00215 | 0.0347 |
| PUM2 | 0.00215 | 0.0348 |
| UBC | 0.00216 | 0.0348 |
| TTC16 | 0.00215 | 0.0348 |
| MCC | 0.00218 | 0.0351 |
| NOM1 | 0.00219 | 0.0353 |
| TMEM8B | 0.00219 | 0.0353 |
| C19orf84 | 0.0022 | 0.0353 |
| RUNX2 | 0.0022 | 0.0353 |
| SSU72 | 0.0022 | 0.0353 |
| TMBIM6 | 0.00219 | 0.0353 |
| FLNB-AS1 | 0.00221 | 0.0354 |
| KCNJ14 | 0.00221 | 0.0354 |
| ARL5B | 0.00221 | 0.0354 |
| COX10-DT | 0.00221 | 0.0354 |
| RFC2 | 0.00224 | 0.0356 |
| NUDT3 | 0.00224 | 0.0356 |
| GPR137 | 0.00224 | 0.0356 |
| MSS51 | 0.00223 | 0.0356 |
| PAQR4 | 0.00224 | 0.0356 |
| ZBTB16 | 0.00224 | 0.0357 |
| UQCR11 | 0.00225 | 0.0357 |
| AGPAT5 | 0.00228 | 0.0359 |
| ZBED3-AS1 | 0.00228 | 0.0359 |
| STT3B | 0.00227 | 0.0359 |
| MUC12 | 0.00228 | 0.0359 |
| TRAPPC4 | 0.00228 | 0.0359 |
| ANKRD44 | 0.00226 | 0.0359 |
| MCOLN1 | 0.00227 | 0.0359 |
| RNF34 | 0.00226 | 0.0359 |
| NORAD | 0.00228 | 0.0359 |
| FANCG | 0.00227 | 0.0359 |
| FAM106A | 0.00227 | 0.0359 |
| TIMM8A | 0.00229 | 0.036 |
| MATCAP2 | 0.0023 | 0.0361 |
| RHBDD2 | 0.00232 | 0.0363 |
| LINC02019 | 0.00231 | 0.0363 |
| MIIP | 0.00231 | 0.0363 |
| SON | 0.00233 | 0.0364 |
| HSP90AB1 | 0.00232 | 0.0364 |
| IQCA1 | 0.00234 | 0.0365 |
| RBM12B | 0.00234 | 0.0365 |
| IRF2BP2 | 0.00234 | 0.0365 |
| HSPA13 | 0.00236 | 0.0366 |
| SCAF11 | 0.00236 | 0.0366 |
| ELOVL6 | 0.00236 | 0.0366 |
| CEP78 | 0.00235 | 0.0366 |
| MKRN1 | 0.00235 | 0.0366 |
| LARP1 | 0.00236 | 0.0366 |
| AGPAT4 | 0.00236 | 0.0366 |
| DCHS1-AS1 | 0.00235 | 0.0366 |
| HIGD1A | 0.00237 | 0.0367 |
| NRBP1 | 0.00237 | 0.0367 |
| C2CD2 | 0.00238 | 0.0368 |
| SAXO2 | 0.00238 | 0.0368 |
| CYLD | 0.0024 | 0.0369 |
| CENPV | 0.0024 | 0.0369 |
| CDO1 | 0.0024 | 0.0369 |
| PHF2P2 | 0.00239 | 0.0369 |
| TSPO | 0.00239 | 0.0369 |
| REEP5 | 0.00241 | 0.0369 |
| AAK1 | 0.00241 | 0.0369 |
| ATF4 | 0.0024 | 0.0369 |
| STUB1 | 0.00241 | 0.037 |
| ZCCHC17 | 0.00242 | 0.037 |
| GPRC5B | 0.00242 | 0.0371 |
| CLIC3 | 0.00243 | 0.0371 |
| RORC | 0.00243 | 0.0372 |
| CACNB3 | 0.00243 | 0.0372 |
| AUP1 | 0.00244 | 0.0372 |
| SASS6 | 0.00244 | 0.0373 |
| SKOR1 | 0.00245 | 0.0373 |
| RALGAPA1 | 0.00246 | 0.0374 |
| YWHAH | 0.00245 | 0.0374 |
| TTLL3 | 0.00248 | 0.0376 |
| SENP7 | 0.00247 | 0.0376 |
| UBE4A | 0.00249 | 0.0378 |
| ARRB2 | 0.00251 | 0.0379 |
| PREPL | 0.0025 | 0.0379 |
| ZSCAN30 | 0.00251 | 0.0379 |
| RTN2 | 0.0025 | 0.0379 |
| ZC3H13 | 0.00252 | 0.038 |
| CDR2 | 0.00252 | 0.038 |
| SMARCAD1 | 0.00252 | 0.038 |
| TMEM123 | 0.00252 | 0.038 |
| PRDX5 | 0.00255 | 0.0382 |
| CKS1B | 0.00254 | 0.0382 |
| DSG2 | 0.00255 | 0.0382 |
| LSM10 | 0.00254 | 0.0382 |
| SPON2 | 0.00254 | 0.0382 |
| SH3BGRL3 | 0.00256 | 0.0382 |
| GPATCH2L | 0.00256 | 0.0382 |
| LEP | 0.00253 | 0.0382 |
| MAMDC4 | 0.00256 | 0.0382 |
| UBL7 | 0.00256 | 0.0382 |
| ZFHX3-AS1 | 0.00255 | 0.0382 |
| MARCHF6 | 0.00257 | 0.0383 |
| STK25 | 0.00256 | 0.0383 |
| NEMP2 | 0.00257 | 0.0383 |
| MVD | 0.00258 | 0.0384 |
| ATXN7 | 0.00259 | 0.0385 |
| FOXN2 | 0.0026 | 0.0386 |
| DPCD | 0.0026 | 0.0386 |
| SELENOW | 0.0026 | 0.0386 |
| RNF166 | 0.00262 | 0.0388 |
| STX5 | 0.00262 | 0.0388 |
| SHC2 | 0.00261 | 0.0388 |
| SP6 | 0.00263 | 0.0389 |
| FAS | 0.00263 | 0.0389 |
| DTD2 | 0.00263 | 0.0389 |
| RBBP5 | 0.00264 | 0.039 |
| UBN2 | 0.00266 | 0.0392 |
| LINGO2 | 0.00267 | 0.0393 |
| SAP30BP | 0.00267 | 0.0393 |
| CHRNE | 0.00267 | 0.0393 |
| ZKSCAN1 | 0.00269 | 0.0396 |
| TMCC2 | 0.00271 | 0.0399 |
| CNN2P1 | 0.00273 | 0.0401 |
| COL4A4 | 0.00273 | 0.0401 |
| SUCO | 0.00275 | 0.0403 |
| MRPL13 | 0.00275 | 0.0403 |
| KYNU | 0.00278 | 0.0406 |
| HECTD3 | 0.00278 | 0.0406 |
| AP4E1 | 0.00278 | 0.0406 |
| TEX264 | 0.00278 | 0.0406 |
| U2SURP | 0.00279 | 0.0407 |
| MIEF1 | 0.00282 | 0.0408 |
| LCNL1 | 0.00281 | 0.0408 |
| RBX1 | 0.00281 | 0.0408 |
| NARF | 0.00281 | 0.0408 |
| NID1 | 0.00281 | 0.0408 |
| NUDT18 | 0.00282 | 0.0408 |
| GNGT2 | 0.0028 | 0.0408 |
| SIRT2 | 0.00282 | 0.0409 |
| PRSS40A | 0.00283 | 0.0409 |
| CREB3 | 0.00285 | 0.0412 |
| EIF4E2 | 0.00286 | 0.0413 |
| TMCC1-DT | 0.00286 | 0.0413 |
| IKZF5 | 0.00289 | 0.0415 |
| PTRHD1 | 0.00288 | 0.0415 |
| SLC2A1 | 0.00288 | 0.0415 |
| HMG20A | 0.00289 | 0.0415 |
| DISP2 | 0.00288 | 0.0415 |
| TCERG1 | 0.00289 | 0.0415 |
| FUS | 0.00289 | 0.0415 |
| FARSB | 0.00289 | 0.0415 |
| CX3CR1 | 0.00291 | 0.0417 |
| SUPT4H1 | 0.00293 | 0.0419 |
| SLC35F5 | 0.00294 | 0.0421 |
| SEC11A | 0.00295 | 0.0421 |
| ZNF653 | 0.00296 | 0.0423 |
| NAA38 | 0.00296 | 0.0423 |
| LARS1 | 0.00296 | 0.0423 |
| NCOA3 | 0.00298 | 0.0424 |
| BPGM | 0.00298 | 0.0424 |
| CAMSAP2 | 0.00297 | 0.0424 |
| UBE2A | 0.00298 | 0.0424 |
| ALOX12B | 0.00299 | 0.0425 |
| FEM1B | 0.00299 | 0.0425 |
| RAB1B | 0.00299 | 0.0425 |
| H2AC25 | 0.003 | 0.0426 |
| CDC25C | 0.003 | 0.0426 |
| LINC02772 | 0.00303 | 0.0428 |
| CYB561D2 | 0.00303 | 0.0428 |
| MTERF4 | 0.00303 | 0.0428 |
| PATZ1 | 0.00304 | 0.0429 |
| CCDC65 | 0.00304 | 0.0429 |
| OGT | 0.00304 | 0.0429 |
| ARMC9 | 0.00306 | 0.0431 |
| NET1 | 0.00306 | 0.0431 |
| CCR4 | 0.00307 | 0.0433 |
| DPPA4 | 0.00309 | 0.0434 |
| ZFP62 | 0.00309 | 0.0434 |
| NUDT7 | 0.00308 | 0.0434 |
| DEDD | 0.0031 | 0.0435 |
| TAF3 | 0.00311 | 0.0436 |
| C1orf216 | 0.00311 | 0.0436 |
| LINC00504 | 0.00313 | 0.0438 |
| ATAD2B | 0.00315 | 0.0439 |
| P2RY1 | 0.00315 | 0.0439 |
| TRGV9 | 0.00315 | 0.0439 |
| HMG20B | 0.00315 | 0.0439 |
| ZNF606 | 0.00314 | 0.0439 |
| DCK | 0.00315 | 0.0439 |
| GTF2I | 0.00316 | 0.044 |
| CDKN2D | 0.00317 | 0.044 |
| ATOX1 | 0.00317 | 0.044 |
| LMAN1 | 0.00318 | 0.044 |
| SLC18B1 | 0.00317 | 0.044 |
| PSMB1 | 0.00316 | 0.044 |
| NAA15 | 0.00317 | 0.044 |
| NR4A2 | 0.00319 | 0.0441 |
| OCEL1 | 0.00319 | 0.0441 |
| ZFYVE19 | 0.00319 | 0.0441 |
| PPME1 | 0.00321 | 0.0444 |
| NKD1 | 0.00322 | 0.0445 |
| ZNF407 | 0.00324 | 0.0447 |
| SNX3 | 0.00323 | 0.0447 |
| TMEM52B | 0.00324 | 0.0447 |
| SIGIRR | 0.00327 | 0.045 |
| RNPEPL1 | 0.00326 | 0.045 |
| TMEM161B-DT | 0.00328 | 0.0451 |
| DOK2 | 0.00329 | 0.0452 |
| LINC01521 | 0.0033 | 0.0453 |
| SRA1 | 0.0033 | 0.0453 |
| UCP3 | 0.00331 | 0.0454 |
| CBX5 | 0.00331 | 0.0454 |
| FGR | 0.00331 | 0.0454 |
| ODAD3 | 0.00332 | 0.0454 |
| DEFA1 | 0.00335 | 0.0457 |
| PAQR5 | 0.00334 | 0.0457 |
| IL10RA | 0.00335 | 0.0457 |
| NDUFB1 | 0.00335 | 0.0457 |
| ARGLU1 | 0.00337 | 0.0459 |
| ROMO1 | 0.00339 | 0.0461 |
| ZNF891 | 0.00339 | 0.0461 |
| ADNP | 0.00339 | 0.0461 |
| APBA2 | 0.0034 | 0.0462 |
| POLR2C | 0.0034 | 0.0462 |
| LINC01871 | 0.0034 | 0.0462 |
| NDUFA4 | 0.0034 | 0.0462 |
| CRTAM | 0.00342 | 0.0463 |
| BNC2 | 0.00342 | 0.0463 |
| FOXD2 | 0.00343 | 0.0465 |
| TRBV6-2 | 0.00344 | 0.0466 |
| RAP1GAP2 | 0.00346 | 0.0467 |
| PEF1 | 0.00346 | 0.0467 |
| ICAM5 | 0.00345 | 0.0467 |
| ARAF | 0.00347 | 0.0468 |
| PSMA6 | 0.00348 | 0.0469 |
| ZNF496-DT | 0.00348 | 0.0469 |
| SREK1 | 0.00351 | 0.047 |
| CLIC1 | 0.0035 | 0.047 |
| ZNF541 | 0.00351 | 0.047 |
| TMEM86B | 0.00351 | 0.047 |
| C1QTNF6 | 0.0035 | 0.047 |
| AP1S1 | 0.0035 | 0.047 |
| THSD7A | 0.00354 | 0.0473 |
| CHRM3-AS2 | 0.00353 | 0.0473 |
| MVP | 0.00354 | 0.0473 |
| LPCAT1 | 0.00355 | 0.0474 |
| JADE3 | 0.00356 | 0.0475 |
| MSI2 | 0.00355 | 0.0475 |
| DDIT4L | 0.00357 | 0.0477 |
| SELENOT | 0.00359 | 0.0478 |
| UCP2 | 0.00359 | 0.0478 |
| KLHL15 | 0.00358 | 0.0478 |
| AP1AR | 0.0036 | 0.0479 |
| COTL1 | 0.00362 | 0.048 |
| STKLD1 | 0.00363 | 0.048 |
| ZNF556 | 0.00363 | 0.048 |
| MYB | 0.00362 | 0.048 |
| ZNF683 | 0.00363 | 0.048 |
| RRP1B | 0.00361 | 0.048 |
| ALG10B | 0.00361 | 0.048 |
| ZNF711 | 0.00363 | 0.048 |
| BHLHE40-AS1 | 0.00366 | 0.0481 |
| GOPC | 0.00366 | 0.0481 |
| ATP9B | 0.00366 | 0.0481 |
| ZNF776 | 0.00364 | 0.0481 |
| CCDC137 | 0.00365 | 0.0481 |
| NUAK1 | 0.00364 | 0.0481 |
| COPS3 | 0.00365 | 0.0481 |
| JMY | 0.00365 | 0.0481 |
| ITGB1P1 | 0.00367 | 0.0482 |
| OLR1 | 0.00367 | 0.0482 |
| TAFA1 | 0.00368 | 0.0483 |
| CHD9 | 0.00373 | 0.0488 |
| AK3P3 | 0.00373 | 0.0488 |
| TRIM51BP | 0.00372 | 0.0488 |
| TMEM30B | 0.00373 | 0.0489 |
| GTF2IRD1 | 0.00374 | 0.0489 |
| CUL5 | 0.00374 | 0.0489 |
| ZFYVE9 | 0.00375 | 0.0489 |
| VN1R83P | 0.00375 | 0.0489 |
| GXYLT1 | 0.00378 | 0.0492 |
| KIF2A | 0.00378 | 0.0492 |
| SEMA7A | 0.00378 | 0.0492 |
| ZSCAN12 | 0.00378 | 0.0492 |
| AGFG2 | 0.00379 | 0.0492 |
| TMBIM1 | 0.0038 | 0.0493 |
| CBLL1 | 0.0038 | 0.0493 |
| MEOX1 | 0.0038 | 0.0493 |
| PCBP2-OT1 | 0.00381 | 0.0494 |
| PIGZ | 0.00382 | 0.0494 |
| ZBTB21 | 0.00382 | 0.0494 |
| THOC2 | 0.00383 | 0.0495 |
| SNAPIN | 0.00383 | 0.0496 |
| STOML2 | 0.00385 | 0.0497 |
| SLC4A3 | 0.00385 | 0.0497 |
| HAUS1 | 0.00386 | 0.0498 |
| RNF25 | 0.00387 | 0.0498 |
| HERPUD2-AS1 | 0.00387 | 0.0498 |
| PBXIP1 | 0.00386 | 0.0498 |
| RAB4B-EGLN2 | 0.00386 | 0.0498 |
| ZNF844 | 0.00388 | 0.0499 |
| MGA | 0.00389 | 0.0499 |
| RPRD1B | 0.0039 | 0.05 |
| LRRC75A | 0.0039 | 0.05 |

Supplementary Table 8. Selected 241 CpG features and age feature for Epigenome model

| **position2** | **pvalue** | **adjusted_pvalue** |
| --- | --- | --- |
| Age | 5.52E-15 | 3.66E-09 |
| chr12:3468532 | 1.56E-09 | 0.000517 |
| chr10:13527445 | 5.32E-09 | 0.00118 |
| chr2:163733757 | 0.000000025 | 0.00207 |
| chr3:51707355 | 1.47E-08 | 0.00207 |
| chr3:51707359 | 1.99E-08 | 0.00207 |
| chr10:13527425 | 2.48E-08 | 0.00207 |
| chr7:120989655 | 1.85E-08 | 0.00207 |
| chr15:39172406 | 2.93E-08 | 0.00209 |
| chr6:138500217 | 3.77E-08 | 0.00209 |
| chr10:13527476 | 0.000000035 | 0.00209 |
| chr7:8442746 | 0.000000035 | 0.00209 |
| chr3:51707396 | 6.69E-08 | 0.00342 |
| chr3:50350420 | 0.000000104 | 0.00494 |
| chr10:33273318 | 0.000000189 | 0.00562 |
| chr2:163475092 | 0.00000018 | 0.00562 |
| chr10:22322823 | 0.000000184 | 0.00562 |
| chr11:64661527 | 0.000000195 | 0.00562 |
| chr6:147207385 | 0.000000154 | 0.00562 |
| chr12:72271117 | 0.000000176 | 0.00562 |
| chr5:146340339 | 0.000000157 | 0.00562 |
| chr3:51707379 | 0.000000171 | 0.00562 |
| chr19:37334407 | 0.000000155 | 0.00562 |
| chr17:48631657 | 0.000000224 | 0.00576 |
| chr2:163733746 | 0.000000225 | 0.00576 |
| chr2:66427512 | 0.000000218 | 0.00576 |
| chr3:160450348 | 0.000000308 | 0.00684 |
| chr5:140041317 | 0.00000033 | 0.00684 |
| chr11:111456246 | 0.000000295 | 0.00684 |
| chr22:46057958 | 0.000000327 | 0.00684 |
| chr19:37334633 | 0.00000032 | 0.00684 |
| chr2:163733755 | 0.000000302 | 0.00684 |
| chr2:189584383 | 0.000000361 | 0.00704 |
| chr1:2213391 | 0.000000359 | 0.00704 |
| chr19:57934974 | 0.000000448 | 0.00816 |
| chr10:22319696 | 0.00000044 | 0.00816 |
| chr2:163475164 | 0.000000455 | 0.00816 |
| chr2:163733762 | 0.000000528 | 0.00923 |
| chr19:37334594 | 0.000000549 | 0.00935 |
| chr10:22317973 | 0.00000062 | 0.01 |
| chr1:24392093 | 0.000000681 | 0.01 |
| chr19:37334670 | 0.00000068 | 0.01 |
| chr19:37334654 | 0.000000649 | 0.01 |
| chr3:156816319 | 0.000000678 | 0.01 |
| chr3:48550120 | 0.000000663 | 0.01 |
| chr4:127784306 | 0.000000721 | 0.0104 |
| chr22:46044342 | 0.000000819 | 0.0116 |
| chr4:123786416 | 0.00000101 | 0.0122 |
| chr13:57633179 | 0.00000101 | 0.0122 |
| chr4:110641443 | 0.000000884 | 0.0122 |
| chr19:37334672 | 0.000000956 | 0.0122 |
| chr2:4111316 | 0.000000958 | 0.0122 |
| chr19:19514206 | 0.000000977 | 0.0122 |
| chr5:146340348 | 0.000000982 | 0.0122 |
| chr11:72813280 | 0.000000958 | 0.0122 |
| chr7:41696248 | 0.00000104 | 0.0123 |
| chr19:37334416 | 0.00000119 | 0.0138 |
| chr19:10836557 | 0.00000122 | 0.0139 |
| chr7:41696175 | 0.00000125 | 0.0141 |
| chr10:86970039 | 0.0000013 | 0.0143 |
| chr4:41215253 | 0.0000014 | 0.0152 |
| chr6:90295082 | 0.00000145 | 0.0156 |
| chr7:27103799 | 0.0000015 | 0.0158 |
| chr3:160450307 | 0.00000165 | 0.0169 |
| chr7:32299483 | 0.00000164 | 0.0169 |
| chr2:72144012 | 0.0000017 | 0.0171 |
| chr3:149377682 | 0.00000176 | 0.0175 |
| chr17:21452697 | 0.00000183 | 0.0176 |
| chr9:95046624 | 0.0000018 | 0.0176 |
| chr11:132865919 | 0.00000198 | 0.0181 |
| chr10:122954592 | 0.00000198 | 0.0181 |
| chr20:60472409 | 0.00000194 | 0.0181 |
| chr2:66427592 | 0.00000193 | 0.0181 |
| chr10:111078647 | 0.00000209 | 0.0188 |
| chr11:380393 | 0.0000022 | 0.019 |
| chr11:18455995 | 0.00000215 | 0.019 |
| chr4:121950125 | 0.0000022 | 0.019 |
| chr14:36654573 | 0.00000242 | 0.0198 |
| chr9:12814418 | 0.00000244 | 0.0198 |
| chr2:235937918 | 0.00000239 | 0.0198 |
| chr5:151924796 | 0.00000239 | 0.0198 |
| chr1:90533818 | 0.00000233 | 0.0198 |
| chr20:45304675 | 0.00000255 | 0.0202 |
| chr5:146340464 | 0.00000253 | 0.0202 |
| chr3:127036552 | 0.00000259 | 0.0202 |
| chr7:122885624 | 0.00000264 | 0.0204 |
| chr6:168454465 | 0.0000027 | 0.0206 |
| chr19:19514190 | 0.00000279 | 0.0208 |
| chr6:28227250 | 0.0000028 | 0.0208 |
| chr7:27103789 | 0.00000285 | 0.0208 |
| chr1:209742562 | 0.00000284 | 0.0208 |
| chr17:21452758 | 0.00000294 | 0.0212 |
| chr10:75772580 | 0.00000311 | 0.0217 |
| chr6:25652372 | 0.00000308 | 0.0217 |
| chr2:72144121 | 0.00000311 | 0.0217 |
| chr16:29996858 | 0.0000032 | 0.0219 |
| chr3:160450354 | 0.0000032 | 0.0219 |
| chr18:32469220 | 0.00000334 | 0.0224 |
| chr19:37334544 | 0.00000337 | 0.0224 |
| chr2:163733784 | 0.00000337 | 0.0224 |
| chr17:79777290 | 0.00000349 | 0.0228 |
| chr1:8449345 | 0.0000035 | 0.0228 |
| chr4:145618542 | 0.00000365 | 0.0235 |
| chr3:127714880 | 0.00000376 | 0.0235 |
| chr17:49790118 | 0.00000375 | 0.0235 |
| chr2:66427596 | 0.00000378 | 0.0235 |
| chr12:21775727 | 0.00000383 | 0.0235 |
| chr19:37334604 | 0.00000381 | 0.0235 |
| chr3:51707330 | 0.00000391 | 0.0238 |
| chr17:74430222 | 0.00000396 | 0.0239 |
| chr2:144359066 | 0.00000405 | 0.0242 |
| chr8:26183506 | 0.00000435 | 0.0246 |
| chr3:51707369 | 0.00000455 | 0.0246 |
| chr11:45721649 | 0.00000436 | 0.0246 |
| chr7:93904686 | 0.00000442 | 0.0246 |
| chr7:63054203 | 0.00000456 | 0.0246 |
| chr6:87151582 | 0.00000451 | 0.0246 |
| chr15:36580394 | 0.00000447 | 0.0246 |
| chr16:9764492 | 0.00000422 | 0.0246 |
| chr14:73760728 | 0.00000426 | 0.0246 |
| chr10:111078659 | 0.00000447 | 0.0246 |
| chr19:37334639 | 0.00000442 | 0.0246 |
| chr4:3505478 | 0.00000439 | 0.0246 |
| chr8:143042805 | 0.00000462 | 0.0247 |
| chr11:44305622 | 0.00000474 | 0.025 |
| chr7:74374041 | 0.00000475 | 0.025 |
| chr12:120266410 | 0.00000484 | 0.0253 |
| chr17:49226744 | 0.00000498 | 0.0258 |
| chr10:111360774 | 0.00000503 | 0.0259 |
| chr2:127701146 | 0.00000533 | 0.0272 |
| chr12:32534118 | 0.0000057 | 0.0288 |
| chr2:70949293 | 0.00000589 | 0.0288 |
| chr22:50146706 | 0.00000592 | 0.0288 |
| chr18:6413752 | 0.00000578 | 0.0288 |
| chr1:114669149 | 0.00000572 | 0.0288 |
| chr3:169667577 | 0.00000593 | 0.0288 |
| chr11:70103113 | 0.00000587 | 0.0288 |
| chr7:1625278 | 0.000006 | 0.0289 |
| chr6:35343119 | 0.0000063 | 0.0299 |
| chr5:168343227 | 0.00000629 | 0.0299 |
| chr6:87151594 | 0.00000638 | 0.03 |
| chr1:178739551 | 0.00000653 | 0.0305 |
| chr3:51707335 | 0.00000665 | 0.0306 |
| chr20:63126481 | 0.00000668 | 0.0306 |
| chr11:111456180 | 0.00000667 | 0.0306 |
| chr1:111309730 | 0.00000691 | 0.0314 |
| chr13:112171447 | 0.00000701 | 0.0317 |
| chr7:154882763 | 0.00000714 | 0.0318 |
| chr17:42548069 | 0.00000712 | 0.0318 |
| chr1:112705618 | 0.00000721 | 0.0319 |
| chr4:183876310 | 0.00000731 | 0.0322 |
| chr19:37334553 | 0.00000737 | 0.0322 |
| chr14:73712323 | 0.00000756 | 0.0328 |
| chr7:158074137 | 0.00000767 | 0.0331 |
| chr18:37241763 | 0.00000784 | 0.0336 |
| chr7:157904541 | 0.00000791 | 0.0336 |
| chr19:35963712 | 0.00000797 | 0.0337 |
| chr2:235765530 | 0.00000818 | 0.0344 |
| chr12:63150081 | 0.00000828 | 0.0346 |
| chr8:94638995 | 0.0000084 | 0.0348 |
| chr3:149376833 | 0.00000855 | 0.0353 |
| chr1:55216295 | 0.0000088 | 0.0354 |
| chr10:130748110 | 0.00000879 | 0.0354 |
| chr5:141485357 | 0.00000867 | 0.0354 |
| chr12:12952502 | 0.00000871 | 0.0354 |
| chr11:31983918 | 0.0000091 | 0.0359 |
| chr2:46374654 | 0.00000911 | 0.0359 |
| chr2:66427319 | 0.00000908 | 0.0359 |
| chr5:107809357 | 0.00000913 | 0.0359 |
| chr19:8589857 | 0.00000925 | 0.0361 |
| chr10:48465491 | 0.0000096 | 0.0366 |
| chr7:57404039 | 0.00000958 | 0.0366 |
| chr5:141251395 | 0.00000953 | 0.0366 |
| chr2:176083101 | 0.00000951 | 0.0366 |
| chr3:160450364 | 0.0000098 | 0.0367 |
| chr6:147207431 | 0.00000983 | 0.0367 |
| chr3:4868111 | 0.00000982 | 0.0367 |
| chr22:46044338 | 0.00000978 | 0.0367 |
| chr4:785333 | 0.0000101 | 0.0367 |
| chr1:30823875 | 0.00000994 | 0.0367 |
| chr20:50692293 | 0.00001 | 0.0367 |
| chr6:71419887 | 0.0000101 | 0.0367 |
| chr2:86789070 | 0.00000998 | 0.0367 |
| chr18:50559279 | 0.0000103 | 0.0373 |
| chr1:236394441 | 0.0000104 | 0.0373 |
| chr11:66590545 | 0.0000105 | 0.0374 |
| chr8:127795071 | 0.0000107 | 0.0379 |
| chr7:106010026 | 0.0000108 | 0.038 |
| chr9:37030301 | 0.0000109 | 0.038 |
| chr20:2749954 | 0.000011 | 0.038 |
| chr4:183905031 | 0.0000108 | 0.038 |
| chr4:7183437 | 0.000011 | 0.038 |
| chr2:172783217 | 0.0000113 | 0.0388 |
| chr3:71574558 | 0.0000113 | 0.0388 |
| chr11:108691806 | 0.0000118 | 0.0399 |
| chr19:19514170 | 0.0000117 | 0.0399 |
| chr5:64165430 | 0.0000121 | 0.0404 |
| chr18:23134112 | 0.0000121 | 0.0404 |
| chr16:89097372 | 0.0000123 | 0.041 |
| chr12:109545261 | 0.0000123 | 0.041 |
| chr4:1626554 | 0.0000125 | 0.0412 |
| chr10:80409724 | 0.0000126 | 0.0415 |
| chr10:22322960 | 0.0000128 | 0.0416 |
| chr17:20784480 | 0.0000128 | 0.0416 |
| chr15:60945559 | 0.000013 | 0.0419 |
| chr19:37334418 | 0.0000129 | 0.0419 |
| chr2:45644350 | 0.0000131 | 0.0419 |
| chr9:132486717 | 0.0000131 | 0.0419 |
| chr20:63733803 | 0.0000133 | 0.0422 |
| chr12:102950600 | 0.0000135 | 0.0426 |
| chr4:1626492 | 0.0000136 | 0.0428 |
| chr12:7189070 | 0.0000137 | 0.0429 |
| chr1:232435512 | 0.0000138 | 0.0429 |
| chr18:58400163 | 0.0000138 | 0.0429 |
| chr1:245301568 | 0.0000139 | 0.0429 |
| chr9:13927291 | 0.0000144 | 0.0439 |
| chr7:120989649 | 0.0000145 | 0.0439 |
| chr7:149415286 | 0.0000143 | 0.0439 |
| chr12:9168145 | 0.0000144 | 0.0439 |
| chr3:139044931 | 0.0000147 | 0.0442 |
| chr7:106009852 | 0.0000147 | 0.0442 |
| chr15:41621911 | 0.0000149 | 0.0443 |
| chr3:62369472 | 0.0000149 | 0.0443 |
| chr13:111167086 | 0.0000153 | 0.0451 |
| chr3:27724830 | 0.0000153 | 0.0451 |
| chr7:2607369 | 0.0000154 | 0.0451 |
| chr18:68722183 | 0.0000154 | 0.0451 |
| chr3:49861571 | 0.0000156 | 0.0453 |
| chr6:116100431 | 0.0000157 | 0.0453 |
| chr2:189584606 | 0.0000156 | 0.0453 |
| chr1:15305377 | 0.000016 | 0.0457 |
| chr14:88155080 | 0.0000159 | 0.0457 |
| chr8:115661828 | 0.0000162 | 0.046 |
| chr17:6435010 | 0.0000163 | 0.0461 |
| chr3:160450315 | 0.0000165 | 0.0467 |
| chr7:143287899 | 0.0000167 | 0.0469 |
| chr10:123006818 | 0.0000171 | 0.048 |
| chr1:159077244 | 0.0000172 | 0.0481 |
| chr12:15982097 | 0.0000174 | 0.0482 |
| chr4:47030661 | 0.0000174 | 0.0482 |
| chr16:62711599 | 0.0000175 | 0.0483 |
| chr11:133953230 | 0.0000181 | 0.0495 |
| chr16:88228169 | 0.0000182 | 0.0497 |

Supplementary Table 9. Pearson’s correlation of Euclidean distance between pairs of Health check-up derived ome-components

| Names of pairs | Num. of samples | ED corr. coef. | *p*-values |
| --- | --- | --- | --- |
| (Inflammatome, Psycholome) | 4 | -0.111352 | 8.89E-01 |
| (Vasculome, Psycholome) | 3 | 0.003333 | 9.98E-01 |
| (Inflammatome, Physiome) | 66 | 0.004105 | 9.74E-01 |
| (Immunome, Psycholome) | 4 | 0.006714 | 9.93E-01 |
| (Immunome, Physiome) | 443 | 0.018525 | 6.97E-01 |
| (Vasculome, Metabolome) | 809 | 0.018889 | 5.92E-01 |
| (Psycholome, Physiome) | 19 | 0.026713 | 9.14E-01 |
| (Vasculome, Immunome) | 793 | 0.06209 | 8.06E-02 |
| (Psycholome, Metabolome) | 3 | 0.073135 | 9.53E-01 |
| (Inflammatome, Immunome) | 304 | 0.093516 | 1.04E-01 |
| (Immunome, Metabolome) | 1038 | 0.096597 | 1.84E-03 |
| (Inflammatome, Vasculome) | 103 | 0.178525 | 7.12E-02 |
| (Physiome, Metabolome) | 395 | 0.193111 | 1.12E-04 |
| (Inflammatome, Metabolome) | 201 | 0.208648 | 2.95E-03 |
| (Vasculome, Physiome) | 311 | 0.508076 | 8.15E-22 |

Num, Number; ED, Euclidean distance; corr coef, Pearson’s correlation coefficient

Supplementary Table 10. The slope values of the mean Euclidean distance per age group. The group average age was calculated by averaging the mean ages of two groups. For example, G0 and G1 have age intervals of 15 ≤ G0 < 20 and 20 ≤ G1 < 25. The average age for G0 is 17.5, calculated as (15 + 20) / 2. Similarly, the average age for G1 is 22.5, calculated as (20 + 25) / 2. By averaging these values, we obtain 20 = (17.5 + 22.5) / 2.

| Intervals | Group average age | Slope values |
| --- | --- | --- |
| G0 to G1 | 20 | -0.083 |
| G1 to G2 | 25 | -0.224 |
| G2 to G3 | 30 | 0.086 |
| G3 to G4 | 35 | -0.068 |
| G4 to G5 | 40 | 0.155 |
| G5 to G6 | 45 | -0.121 |
| G6 to G7 | 50 | -0.032 |
| G7 to G8 | 55 | 0.246 |
| G8 to G9 | 60 | 0.135 |
| G9 to G10 | 65 | 0.135 |
| G10 to G11 | 70 | 0.154 |
| G11 to G12 | 75 | 0.088 |
| G12 to G13 | 78.75 | -0.473 |

15 ≤ G0 < 20, 20 ≤ G1 < 25, 25 ≤ G2 < 30, 30 ≤ G3 < 35, 35 ≤ G4 < 40, 40 ≤ G5 < 45, 45 ≤ G6 < 50, 50 ≤ G7 < 55, 55 ≤ G8 < 60, 60 ≤ G9 < 65, 65 ≤ G10 < 70, 70 ≤ G11 < 75, 75 ≤ G12 < 80, 80 ≤ G13 years

Supplementary Table 11. Health check-up information of two subjects included in age group G0.

| ID | KGP-01380 | KGP-00742 | PSH | Units |
| --- | --- | --- | --- | --- |
| ED Ph | 5.037846 | 5.255526 | M: 0  F: 0 |  |
| Age | late teens | late teens | - | years |
| Sex | M | M | - |  |
| WC | 88 | 80.5 | M: 75.34  F: 70.80 | cm |
| BMI | 26.8 | 22.7 | M: 22.26  F: 21.79 | kg/m^2^ |
| SBP | 110 | 126 | M: 112.29  F: 106.44 | mmHg |
| DBP | 60 | 83 | M: 69.11  F: 66.46 | mmHg |
| Hb | 15.9 | 16.2 | M: 14.87  F: 13.44 | g/dL |
| FBS | 89 | 87 | M: 95.58  F: 92.20 | mg/dl |
| HDL | 37 | 80 | M: 48.06  F: 54.10 | mg/dl |
| LDL | 81 | 106 | M: 115  F: 115 | mg/dl |
| TG | 209 | 123 | M: 139.32  F: 105.67 | mg/dl |
| TC | 139 | 194 | M: 178.08  F: 181.93 | mg/dl |
| Cr | 0.81 | 0.86 | M: 0.85  F: 0.64 | mg/dl |
| ALT | 9 | 16 | M: 26.77  F: 18.02 | IU/L |
| AST | 13 | 15 | M: 25.69  F: 21.11 | IU/L |
| GGT | 10 | 50 | M: 21  F: 21 | IU/L |

ED Ph, Euclidean distance in Physiome; KGP, Korean Genome Project; PSH, Pseudo Super Healthy; M, Male; F, Female; WC, Waist Circumference; BMI, Body Mass Index; SBP, Systolic Blood Pressure; DBP, Diastolic Blood Pressure; Hb, Hemoglobin; FBS, Fasting Blood Sugar; HDL, High Density Lipoprotein; LDL, Low Density Lipoprotein; TG, Triglycerides; TC, Total cholesterol; Cr, Creatinine; ALT, Alanine Transaminase; AST, Aspartate Transaminase; GGT, Gamma-Glutamyl Transferase

Supplementary Table 12. Health check-up information of six subjects included in age group G13.

| ID | KGP-01078 | KGP-01209 | KGP-03821 | KGP-03859 | KGP-01110 | KGP-03856 | PSH | Units |
| --- | --- | --- | --- | --- | --- | --- | --- | --- |
| ED Ph | 4.799859 | 3.535948 | 5.68566 | 3.736669 | 3.671703 | 4.471392 | M: 0  F: 0 |  |
| Age | early 80s | early 80s | early 80s | early 80s | early 80s | early 80s | - | years |
| Sex | M | M | M | M | F | F | - |  |
| WC | 93 | 85 | 91.5 | 79 | 82 | 74 | M: 75.34  F: 70.80 | cm |
| BMI | 25.9 | 22.7 | 22.6 | 21.2 | 20.1 | 19.2 | M: 22.26  F: 21.79 | kg/m^2^ |
| SBP | 132 | 134 | 135 | 129 | 121 | 145 | M: 112.29  F: 106.44 | mmHg |
| DBP | 69 | 67 | 70 | 72 | 67 | 63 | M: 69.11  F: 66.46 | mmHg |
| Hb | 14.7 | 14 | 13.1 | 13.7 | 13.2 | 11.2 | M: 14.87  F: 13.44 | g/dL |
| FBS | 89 | 81 | 97 | 93 | 76 | 104 | M: 95.58  F: 92.20 | mg/dl |
| HDL | 37 | 43 | 37 | 77 | 79 | 61 | M: 48.06  F: 54.10 | mg/dl |
| LDL | 86 | 110 | 135 | 134 | 108 | 118 | M: 115  F: 115 | mg/dl |
| TG | 82 | 169 | 208 | 69 | 80 | 69 | M: 139.32  F: 105.67 | mg/dl |
| TC | 130 | 162 | 203 | 206 | 188 | 172 | M: 178.08  F: 181.93 | mg/dl |
| Cr | 0.75 | 0.83 | 1.32 | 0.87 | 0.69 | 0.77 | M: 0.85  F: 0.64 | mg/dl |
| ALT | 18 | 17 | 13 | 28 | 20 | 14 | M: 26.77  F: 18.02 | IU/L |
| AST | 15 | 23 | 19 | 28 | 28 | 20 | M: 25.69  F: 21.11 | IU/L |
| GGT | 14 | 32 | 22 | 16 | 13 | 19 | M: 21  F: 21 | IU/L |

### **Supplementary Methods**

Supplementary Method 1. Details of developing the Five Ome-Components of the Healthome Polygon Using Korea10K Health Check-up Data

##### *Korea10K data preprocessing for Physiome component*

Within the Korea10K cohort, each subject may possess multiple health check-up records spanning various years, potentially containing missing values. To compute the Euclidean distance between a pseudo super healthy subject and the Korea10K subjects, the Korea10K data underwent preprocessing to generate Physiome, Metabolome, Vasculome, Immunome, and Inflammatome datasets.

For the Physiome component, if a subject lacked multiomics data, a health check-up record with minimal missing values was selected from among the multiple records available for that subject. In cases where a subject possessed multiomics data and only one health check-up record, that single record was utilized. However, if a subject had multiomics data and multiple health check-up records, the health check-up record with a year closest to the Epigenome sequencing year was chosen. The years of health check-ups for the remaining subjects were retained and employed for selecting corresponding health check-up records in the Metabolome, Vasculome, Immunome, and Inflammatome datasets to ensure data consistency to the greatest extent possible (Supplementary Figure 3). Subsequently, the Interquartile Range method, augmented by a 1.5 multiplication factor, was applied to exclude outliers.

##### *Korea10K data preprocessing for Metabolome, Vasculome, Immunome, and Inflammatome*

To preprocess the data from the four types of ome-components, missing values were initially imputed column-wise using the mean value of each column from multiple rows corresponding to each subject. If missing values persisted after this imputation, those subjects were excluded, indicating a lack of available data for calculation. From the multiple rows pertaining to one subject, a single row with a date matching the year information generated in the Physiome dataset was selected. In cases where the year information for the Physiome dataset did not encompass a specific subject, the row with the latest date was chosen.
